# RAPID: Evaluation of Cas12a Protospacer Nicking and Chimeric Reporters for PAM-independent RNA and DNA diagnostics

**DOI:** 10.1101/2025.07.12.25331452

**Authors:** Idorenyin A. Iwe, Frank X. Liu, Ariel Corsano, Severino Jefferson Ribeiro da Silva, Jennifer Doucet, Serena Singh, Gabriel Lamothe, Riham Zayani, Jessica Nguyen, Quinn Matthews, Justin RJ. Vigar, Pouriya Bayat, Mohammad Simchi, Kristof Bozovicar, Moiz Charania, Sabina Panfilov, Paul Kelly, Rita Cai, Basil P. Hubbard, XiuJun Li, Tony Mazzulli, Jacques P. Tremblay, Yufeng Zhao, Alexander A. Green, Zhigang Li, Shuhuai Yao, Keith Pardee

## Abstract

CRISPR-Cas nucleases have revolutionized diagnostics and biotechnology by providing programmable specificity. Here, we extend the understanding of Cas12a biology with a screen that, unexpectedly, finds that Cas12a *trans* cleavage activity can be modulated by nicks in the protospacer in a position-dependent manner. Wanting to explore the impact of non-conventional *trans* cleavage substrates, we subsequently find that non-specific Cas12a cleavage can be significantly reduced with RNA and chimeric (mixed RNA/DNA) reporter sequences. Exploiting these features and building on emerging PAM-independent Cas12a diagnostics that use engineered DNA activators and split-guide architectures, we introduce RAPID (**R**NA/DNA **A**dvanced chimeric, **P**AM-independent, **I**ntegrated Nicking, **D**iagnostics), a nick-tuned, PAM-duplex-mediated platform for PAM-independent RNA and DNA detection. By strategically introducing a nick within the spacer region, RAPID expands Cas12a detection to include target RNAs, which can be ligated *in situ* to create a hybrid protospacer-target with *trans* cleavage activity matching conventional Cas12a. We then apply RAPID to detect single point mutations in ssDNA and RNA substrates, a challenge for traditional Cas12 and Cas13 systems. In combination with RT-LAMP, RAPID is used for PAM-independent RNA detection in clinical samples, achieving sensitivity down to ∼1 aM and 100% concordance with RT-qPCR for samples with Ct ≤33.

## INTRODUCTION

CRISPR-Cas12a (Cpf1), a class 2, type V endonuclease, is an RNA-guided nuclease that performs *cis* cleavage of dsDNA (or ssDNA) targets and indiscriminate *trans* cleavage of ssDNA [1, 2]. The programmable and specific targeting capabilities of Cas12a have made it a widely applied tool in both nucleic acid detection and gene editing [3–5]. Cas12a-based diagnostics often also incorporate an isothermal amplification method, such as loop-mediated isothermal amplification (LAMP) or recombinase polymerase amplification (RPA), to enhance detection sensitivity [5–7]. Amplification is necessary due to the inherent sensitivity limitations of Cas12a, which typically operates down to the picomolar level [8]. Combined with isothermal pre-amplification, the genetic material of viral pathogens (e.g. human papillomavirus (HPV), Ebola, Zika, and SARS-CoV-2) has been previously detected with sensitivity in the attomolar range, which is comparable to PCR [7, 9–13]. CRISPR methods thus provide next-generation diagnostic approaches for low-burden clinical-grade point-of-need testing without complex instrumentation [14].

Recent advances in CRISPR-based diagnostics have focused on improved enzyme variants and gRNA (guide RNA or CRISPR RNA) design to enable both DNA and RNA nucleic acid diagnostics with enhanced sensitivity, rapidity, specificity, and the capacity for multiplexed testing. More specifically, the newly characterized Cas12a2 and Cas12g variants exclusively target RNA substrates, which broadens the potential applications of the Cas12 family beyond DNA targeting alone [15, 16]. Moreover, gRNA alterations, including 5’ extensions and sequence splits have improved the kinetics of the CRISPR system as well as introduced multiplexing capabilities [17, 18]. As tandem gRNA can increase sensitivity in both Cas12 and Cas13 systems [19, 20], an engineered enzyme containing two gRNA-integrated regions has also enhanced the limit of detection [21, 22]. Protospacer engineering is another strategy to broaden Cas12a’s diagnostic capabilities, as demonstrated by split activators for DNA and RNA detection [23–27].

Here, we focus on engineering the structure and composition of the nucleic acids in complex with the Cas12a enzyme, building on our previous work involving split activators and substrate nicking [25]. To date, there has been limited exploration of protospacer nicks that compromise activator integrity, or the composition of *trans* cleavage substrates. Moreover, the recovery of Cas activation signal in the presence of pre-existing nicks in the DNA substrate remains unexplored. To unlock Cas12a’s full potential for nucleic acid targeting, the requirement for a protospacer adjacent motif (PAM) in target sequences must be alleviated. Engineering Cas9 has yielded variants that possess relaxed PAM requirements; meanwhile, introduction of PAM-presenting oligonucleotides (PAMmers) that are complementary to ssRNA targets has enabled PAM-free activation and cleavage by Cas9 [28]. Thus far, and to the best of our knowledge, PAM-free detection by Cas12a has only been achieved using ssDNA substrates, or methods based on toehold activation and temperature modulation [29, 30].

Advances in CRISPR-based diagnostics have also improved specificity towards detecting single point mutations [30, 31], which can enable detection of pathogen variants (e.g. influenza subtypes) or drug resistance mutations, for example [32]. However, to date, this requires additional sample preparation steps or high temperatures to convert dsDNA to ssDNA [20, 29], features that are incompatible with high-throughput and field-based biosensing applications. An improved ability of Cas proteins to directly detect single nucleotide polymorphisms (SNPs) would significantly expand the range of diagnostic applications.

Here, we begin with a fundamental screen that explores the effect of adding a nick in a stepwise manner across the protospacer sequence. Unexpectedly, we find that nicking modulates the activation of the Cas12a enzyme in a position-dependent manner, resulting in tunable nuclease activity, including muted, semi-muted, and complete activation. This phenomenon reveals that the Cas12a ribonucleoprotein’s ability to cleave nucleic acid targets relies not only on a PAM and base pairing, but also on the integrity of the target sequence at key loci. We also demonstrate, for the first time, that *in situ* ligation of such nicks can fully restore the Cas activation signal. Leveraging these findings, we have developed a PAM-independent nucleic acid targeting platform that primes Cas12a for activation using a separate PAM-duplex strand, independent of the target sequence. The result is unrestricted Cas12a targeting of sequences (e.g. no PAM constraint) and expanded ability to target RNA substrates (**Fig. 1A**).

**Fig. 1:**
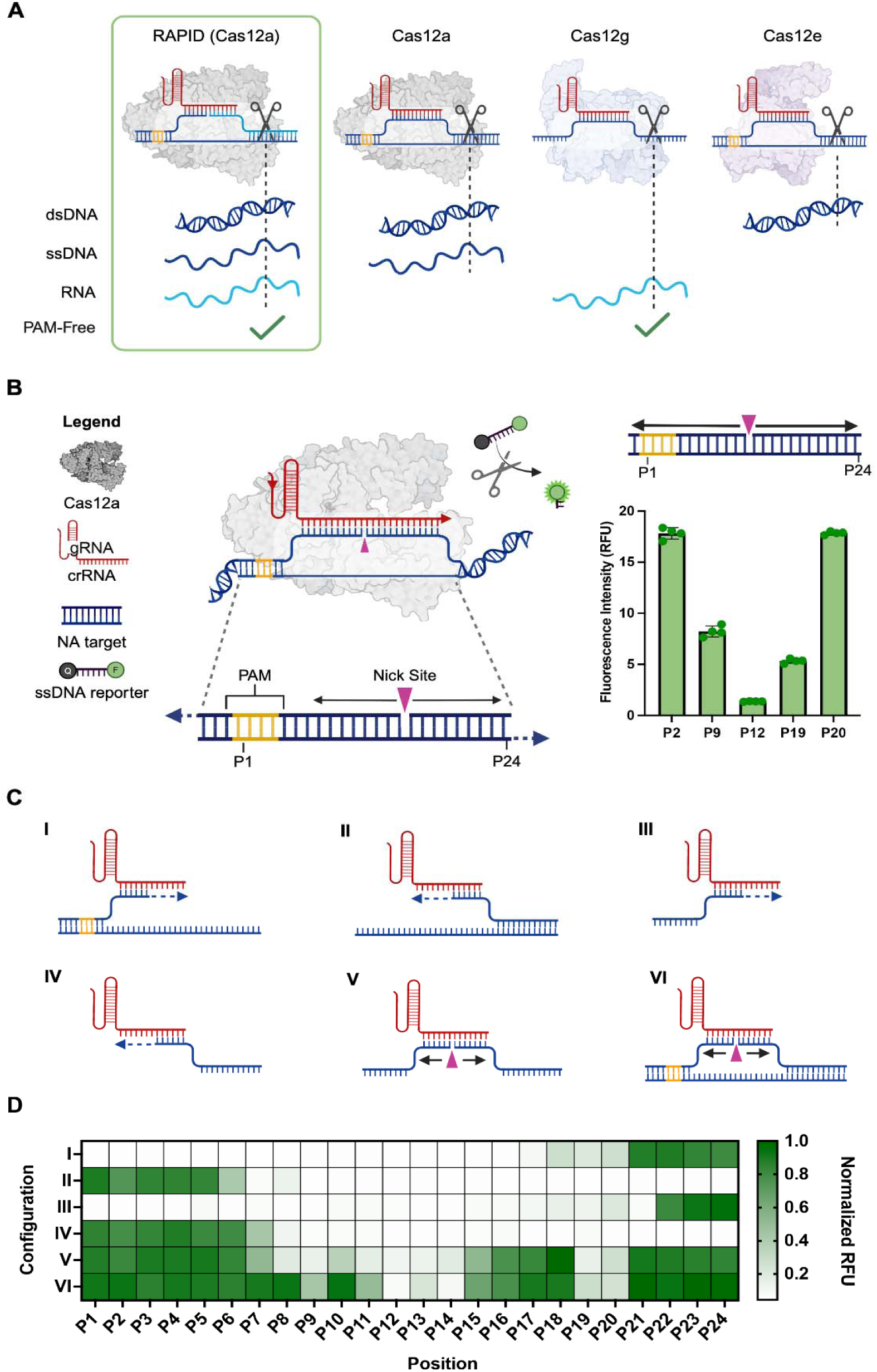
Tunable activation of CRISPR-LbCas12a RAPID system through target sequence breaks. **(A)** Comparison of traditional CRISPR-Cas12a family targeting different nucleic acid substrates with the RAPID system. The RAPID system uniquely detects dsDNA, ssDNA, and RNA without requiring a PAM site on the target nucleic acid. **(B)** Demonstration of tunable activation of the RAPID system using LbCas12a, based on the nick location within the dsDNA. (**Methods 1-3**). Complete activation is observed at positions P2 and P20, partial activation at P9 and P19, and no activation at P12. **(C)** Six foundational configurations of the RAPID system: **(Type I)** Configuration with continuous elongation of the target strand towards the PAM-distal region, while the non-target strand remains fixed. **(Type II)** Similar to (I), but with elongation extending into the PAM motif. **(Types III & IV)** Configurations analogous to (I) and (II), respectively, but without a non-target strand. **(Type V)** Utilizes ssDNA with strategic nick placements within the protospacer region. **(Type VI)** Features dsDNA with single-stranded breaks on the target strand. **(D)** Heat map showing normalized fluorescence intensities resulting from Cas12a activation and *trans* cleavage of the reporter for the six configurations described in (C), across 24 nick locations (P1 to P24) within the protospacer region, including the PAM. n=4 technical replicates; bars represent the arithmetic mean ± SD.

To enhance the performance of the PAM-duplex, we next screened the effect of *trans* cleavage reporter composition on activity. Here, we introduce RNA homopolymers and chimeric (mixed RNA-DNA) reporter sequences. Through this approach, we discovered that Cas12a collateral cleavage reporting using modified RNA, rather than conventional ssDNA, significantly improves the signal-to-noise ratio. By combining the PAM-duplex with these new reporters, we developed a novel platform termed **R**NA/DNA **A**dvanced Chimeric, **P**AM-independent, **I**ntegrated Nicking, **D**iagnostics (RAPID). This platform facilitates direct targeting of miRNA, RNA, ssDNA, and dsDNA downstream of the nick site without compromising Cas12a *trans* cleavage activity. RAPID also demonstrates enhanced performance when used with chimeric reporters.

As a proof of concept, we then apply RAPID to detect point mutations in both ssDNA and miRNA, demonstrating improved specificity over conventional Cas12a detection [2, 33]. When combined with *in situ* ligation for the direct detection of RNAs, the result is a DNA-RNA hybrid target strand formed within the Cas12a clamp which can fully restore activation of nicked targets. Finally, with implementation of an upstream RT-LAMP (reverse transcription-loop-mediated isothermal amplification) reaction, we show that RAPID can also identify mRNA in the attomolar range, a concentration suitable for clinical sample detection. Validation of Cas12a-based RAPID on 21 SARS-CoV-2 patient samples, targeting an RNA sequence that does not contain a PAM and, for samples with Ct ≤ 33, shows 100% concordance to the diagnostic performance of gold standard reverse-transcription quantitative PCR (RT-qPCR) assays. Taken together, we report new insights into the molecular regulation underpinning Cas12a *trans* cleavage through substrate nicking and chimeric reporters, and integrate these features into RAPID to provide high sensitivity, PAM-independent detection, significantly expanding the CRISPR toolbox.

## Materials and Methods

### Reagents and materials

All oligonucleotides including the duplex probes used in this work were synthesized by Integrated DNA Technologies (IDT). All modified oligos were purified by HPLC, while unmodified oligos were only subjected to standard desalting. Acidaminococcus sp (AsCas12a) was purchased from IDT (#10001272). Lachnospiraceae bacterium, LbCas12a (#M0653T), Cas12 diluent (#B0653A), Cas12 reaction buffer (NEBuffer™ r2.1, #B6002S), and RNase inhibitor (#M0314L) were purchased from New England Biolabs (NEB). Magnesium chloride solution (#7786303) was obtained from Sigma Aldrich. Polyethylene glycol/dimethyl sulfoxide solution 50% (w/v) was purchased from Sigma Aldrich (#P7306). DNase/RNase free deionized water (#10977015) from Thermo Fisher Scientific was used in all experiments.

### Assembly of nicked DNA activators (Method 1)

To investigate the effect of nicks on the target strand of dsDNA (target strand: top strand of dsDNA that binds to the gRNA), we assembled three DNA oligonucleotides, designated as NTS, TS Fragment A and TS Fragment B. NTS is the non-target strand while TS is the target strand. DNA oligos were assembled in a 1X phosphate-buffered saline solution and heated to 95 °C for 5 min before cooling down to room temperature to facilitate formation of nicked dsDNA. Nicked DNA activators with nicks at 24 different target points were systematically designed by keeping the NTS constant while varying TS Fragments A and B. All sequences are presented in **Table S1**. Additionally, nicks on the NTS were also tested following the same annealing process. Sequences in **Table S2**. We also tested another nick containing dsDNA to establish generality of concept (see **Table S3** for sequences). All sequences are contained in **Table S1**.

### CRISPR-Cas12a assay (Method 2)

Following the annealing process, CRISPR-Cas12 assay components were prepared in final concentration of 1X NEB 2.1 buffer to a final reaction volume of 40 µL, adjusted with nuclease-free water. Final concentrations of assay components were as follows: 50 nM LbCas12a, 50 nM gRNA, 40 U RNase inhibitor, 125 nM DNA reporter and 10 nM annealed dsDNA. This mixture was aliquoted in 10 µL volumes into a 96-well plate to enable four technical replicates per condition.

Reactions were monitored for 2 hours at 37 °C using the Roche LightCycler 480 II. Fluorescence data at 100 min were normalized and analyzed to assess nicking effects (**Fig. 1** and **Supplementary Fig. S1**). For clarity, the fluorescence signal in each configuration was normalized to the maximum signal observed within that configuration, such that the highest value was scaled to 1. This enabled comparison across all configurations within a single heatmap representation. Sequences of all nucleic acids used are listed in **Tables S1-2**.

To confirm the generality of our findings, additional experiments employing a different gRNA (gRNA_02) within some regions exhibiting tunable Cas12 activation were conducted using the same protocol. Results from these experiments are presented in **Supplementary Fig. S1G**, and the corresponding nucleic acid sequences are provided in **Table S3**.

### Preparation of nucleic acid targets including RNAs (Method 3)

All dsDNA were synthesize using gBlock of ssDNA obtained from IDT. T7-containing ssDNA templates for RNA including miRNA and crRNA transcription along with the primers were also purchased from IDT. 40 nM of the T7-containing ssDNA template was amplified using Q5 PCR kit (NEB #M0491) to form the T7-containing dsDNA. RNAs were synthesized using the HiScribe T7 High Yield RNA Synthesis Kit (NEB # E2050) by spiking in the amplified DNA templates into the reaction, and the reaction was performed at 37 °C from 4 to 16 h. The resulting reaction was treated with 4 units of DNase I (NEB #E2050 -M0303AVIAL) for 15 mins at 37 °C before purification with the Monarch RNA Cleanup Kit (NEB #T2040) and quantification with the NanoDrop One spectrophotometer (Thermo Fisher Scientific).

### Melting temperature analysis (Method 4)

Equimolar amounts of target ssDNA or nicked target ssDNA were mixed with the complementary ssDNA oligonucleotide in NEBuffer 2.1 (NEB) to a final concentration of 2 µM. SYBR Green I was added to a final concentration of 1×. The solution was moved to a CFX96 Real-Time System (BioRad) and incubated for 5 min at 95 °C, then cooled to 25 °C at 0.1 °C s−1 to anneal the DNA duplex. The duplex was then heated at a rate of 0.1 °C s−1 to 95 °C and the corresponding fluorescent signal was used to generate a melt curve. Data analysis was performed using CFX Software (Biorad).

### Gel electrophoresis (Method 5)

Cas12a *trans* cleavage of X174 virion DNA in the presence of nicked dsDNA fragments was evaluated by gel electrophoresis. dsDNA fragments with nicks along the target strand, TS_02 -where 02 represent another dsDNA sequence, at positions P8, P12, P13, and P21 were prepared by the annealing process detailed previously. CRISPR-Cas12 assays included 90 nM LbCas12a, 90 nM gRNA, 20 U RNase inhibitor, 31.25 mM MgCl_2_, X174 virion DNA (NEB N3023S), and 10 nM of annealed nicked dsDNA all in 20 µL 1X NEB r2.1 buffer. The reaction was then incubated for 30 minutes at 37 °C.

Reaction mixtures (20 µL) were resolved by 0.8% agarose gels stained with SYBR™ Safe (Thermo Fisher Scientific S33102). Electrophoretic separation was conducted at 120 V for 1 hour and imaging was performed using a Bio-Rad ChemiDoc imaging system. The results of these experiments are presented in **supplementary Fig. S1D**. All nucleic acid sequences used in these experiments can be found in **Table S1**.

Similar experiments were conducted using dsDNA activators with unperturbed target strands (TS_02) and non-target strands nicked at positions S9, S11 and S16 (NTS_02). Results from these experiments are shown in **Supplementary Fig. S1E**. All sequences for these experiments are listed in **Table S2**.

### RAPID assay (Method 6)

#### ssDNA and RNA Detection

A 50 µL reaction mixture was prepared containing the following components: 1X r2.1 NEB buffer, 90 nM LbCas12a (or AsCas12a), 50 U RNase inhibitor, 20 nM duplex probe, 90 nM gRNA, 31.25 mM MgCl, 500 nM ssDNA reporter (R1), and 50 nM ssDNA or RNA trigger. From this mixture, 15 µL was aliquoted into a 384-well clear reaction plate (Fisher Scientific 4483285) for technical triplicates. The plate was centrifuged for 2 minutes at 2000 × g and 4 °C using. Following centrifugation, the plate was transferred to a QuantStudio 5 real-time PCR system (ThermoFisher Scientific), where fluorescence was measured over a period of 2 hours at 37 °C.

#### dsDNA Detection

A 50 µL reaction mixture was prepared as follows: 1X r2.1 NEB buffer, 90 nM LbCas12a (or AsCas12a), 50 U RNase inhibitor, 200 nM universal PAM, 90 nM gRNA, 31.25 mM MgCl, 500 nM ssDNA reporter (R1), 5.81% PEG8000, and 50 nM PAM-independent dsDNA trigger. Similar to the ssDNA/RNA detection setup, 15 µL of this reaction mixture was aliquoted into a 384-well clear reaction plate. The remaining steps, including centrifugation and fluorescence measurement, were carried out as described above. All nucleic acid sequences are detailed in **Table S6**.

### Two-pot RPA–RAPID (Method 7)

To demonstrate the practicality of RAPID, we adapted an established RPA–CRISPR diagnostic for monkey pox virus [34] to an RPA–RAPID diagnostic. We used published optimized RPA primers and gRNA targeting the monkeypox virus (MPXV) *b6r* gene [34]. For the MPXV *b6r* target, we ordered the *b6r* gene (GenBank: ON563414.3, 165115..166068) as a FLASH gene-to-plasmid, with the pUC57 backbone, from GenScript.

To amplify the *b6r* target, we used the Invitrogen^TM^ Lyo-ready RPA kit (ThermoFisher #A72127). To improve sensitivity, we modified the kits protocol slightly, following their optimization guidelines. Simply, each 20 µL RPA reaction contained 1x reaction buffer, 0.2 mM dNTP mix, 0.3 µM primers (b6r-RPA-fwd and b6r-RPA-rev), 0.06 mg/mL T4 UvsX, 0.06 mg/mL T4 UvsY, 0.4 mg/mL T4 gene protein 32, 0.075 U/µL Bst DNA polymerase, 14 mM MgCl_2_. Each reaction contained 2 µL of *b6r* DNA template with varying final concentrations: 50 pM, 50 fM, and 50 aM pUC57-*b6r* (GenScript), and nuclease-free water for the NTC. We incubated the RPA reactions at 45°C for 30 min, flicking-to-mix after 5 min, in a ProFlex PCR System (ThermoFisher) thermocycler.

To detect the amplified *b6r* target, we used RAPID mode IV system, targeting internal dsDNA, described above (see **Method 6**). To compare the efficiencies of RAPID and DETECTR, we spiked in 1 µL of each RPA reaction into two subsequent RAPID reaction with different gRNA: a regular CRISPR gRNA, targeting a PAM-containing sequence, and a RAPID gRNA, targeting a sequence without a PAM. Reactions were split into three 15 µL technical replicates in a Corning black clear-bottom 384-well plate (#3544). We incubated the plate at 42°C and read the fluorescence (λ_em_= 484 nm, 20 nm bandwidth; λ_em_ = 530 nm, 25 nm bandwidth; gain = 75) 1x/min for 5 h in a BioTek Synergy H1 plate reader (Agilent).

### Fluorescence reporter screening and RPA experiments (Method 8)

**Reporter screening**: to screen the 16 different reporters (including DNA/RNA/chimeric sequences, DNA homopolymers, RNA homopolymers, and chimeric homopolymers) used in this study, a 50 µL reaction mixture was prepared as follows: 1X r2.1 NEB buffer, 90 nM LbCas12a (or AsCas12a), 50 U RNase inhibitor, 20 nM duplex probe, 90 nM gRNA, 31.25 mM MgCl, 10 nM 89-nt ssDNA trigger, and 125 nM of the various reporter types. From this mixture, 15 µL was aliquoted into a 384-well clear reaction plate, following the same procedure as described for ssDNA and RNA detection. The remaining steps, including centrifugation and fluorescence measurement, were performed in the same manner. All sequences are contained in **Table S7**.

**Cas12a RPA experiments**: the RPA-Cas12a reaction is performed in two steps: (**i**) RPA amplification. The RPA pre-amplification was set up starting with TwistAmp® Basic at 37 °C for 20 min. 280mM MgOAc was added to trigger reaction before incubation. Each reaction was 20 μL containing 14 mM MgOAc final concentration. (**ii**) 2 μL of RPA product was added to 18 μL of Cas12a reaction (LbCas12a and AsCas12a) for both conventional ssDNA and chimeric reporters. The CRISPR Cas12a master mix consisted of 1X NEB 2.1 buffer, 50 nM Cas12a, 1U/μL RNase inhibitor murine, 50 nM crRNA and 125 nM ssDNA or chimeric reporters. The fluorescence was then measured every 30 s using Roche Lightcycler 480 II at 37 °C for 30 min.

### miRNA-21 detection (Method 9)

Cas12a reactions were assembled by mixing AsCas12a and gRNA at final concentrations of 90 nM in 1X NEB r2.1 buffer supplemented with 31.25 mM MgCl_2_, 5.81% PEG 8000, and 1U/µL murine RNase inhibitor (NEB). The fluorophore-quencher reporter (R3) and duplex probe (dsDNA co-activator) were added to the Cas12 reaction at final concentrations of 500 nM and 20 nM, respectively, along with the various concentrations (at 20 µL) of the miRNA target in a 50 µL reaction volume on ice. Reactions were transferred into optical 384-well clear plates (Fisher Scientific #4483285) and incubated at 37 °C for 2 hours while the FAM fluorescent intensity was measured every minute using the QuantStudio 5 RT-PCR System. All oligos are listed in **Table S13**. Limit of detection of miRNA-21 calculated from 3σ/S, where σ is the standard deviation of the background signal and S is the slope of the fluorescence of target concentration line.

### Lyophilization (Method 10)

Master mixes for all reaction components including Cas12 proteins, guide RNAs, reporter probes, buffers and salts were prepared as described above, excluding the sample (either miRNA or RT-LAMP amplicons as applicable) and PEG for miRNA reactions. 25 mg/mL each of sucrose and dextran were added to all freeze-dried reactions as a lyoprotectant as described previously [35]. Reactions were flash frozen in liquid nitrogen and freeze-dried overnight in a Labconco FreeZone 6 Liter −84 °C Console Freeze Dryer (catalog #710611200) and vacuum packed with desiccant for storage.

### Mismatch detection in ssDNA and miRNA (Method 11)

All templates containing single or double mismatches in ssDNA were obtained from IDT. A 50 µL reaction mixture was prepared containing the following components: 1X r2.1 NEB buffer, 90 nM LbCas12a, 50 U RNase inhibitor, 20 nM duplex probe, 90 nM gRNA, 31.25 mM MgCl, 500 nM ssDNA reporter (R1), and 10 nM or 2 µM ssDNA with mismatches or WT. From this mixture, 15 µL was aliquoted into a 384-well clear reaction plate (Fisher Scientific #4483285) for technical triplicates. The plate was centrifuged for 2 minutes at 2000 × g and 4 °C. Following centrifugation, the plate was transferred to a QuantStudio 5 real-time PCR system (ThermoFisher Scientific), where fluorescence was measured over a period of 2 hours at 37 °C. We also challenged RAPID’s selectivity using relatively higher and lower oligonucleotide concentrations and found similar performance for both point mutations and paired two-base mismatches (**supplementary Fig. S15B,C**). RNA mismatches were tested the same way, except 5.81% PEG 8000 and chimeric reporter (R3 in place of R1) were added into the reaction and AsCas12a was used instead of LbCas12a. (Sequences in **Tables S14-15**).

#### *Trans* cleavage rate calculation

Background subtracted fluorescent kinetic curves were fit to an exponential association model, following the equation y=A*(1-exp(-k*x)). The rate constant k, representing the *trans* cleavage rate, is plotted for each condition in triplicate. It is important to note that the rate constants are derived from independent fits and are most appropriately compared within, rather than across panels. See **Supplementary Fig. S15**, which highlights the consistency in endpoint fluorescence across WT conditions.

### RT-LAMP assay (Method 12)

RT-LAMP reactions using the WarmStart LAMP 2X Master Mix (E1700S, NEB) were assembled in 30 μL containing 0.2 μM for F3 and B3 primers, 1.6 μM for FIP and BIP primers, 0.4 μM for LF and LB primers, and 1.0 μL of template (water, RNA extracted, or *in vitro* transcribed RNA) was used per reaction. These primers targeted the nucleocapsid protein (**Table S16**). All reactions were prepared on ice, incubated at 61 °C for 30 minutes, then inactivated at 80 °C for 5 minutes.

### RT-LAMP/RAPID reaction for SARS-CoV-2 detection (Method 13)

After the RT-LAMP reaction, 25 µL of the reaction mixture was added to the RAPID reaction mix to achieve a final volume of 50 µL. The final concentrations of the components in the RAPID-LAMP reaction were as follows: 1X r2.1 NEB buffer, 90 nM LbCas12a, 50 U RNase inhibitor, 20 nM PAM duplex 1, 20 nM PAM duplex 2, 90 nM gRNA1, 90 nM gRNA2, 31.25 mM MgCl, 500 nM ssDNA reporter (R1), and 25 µL of the RT-LAMP assay, making up the final volume of 50 µL.

From this mixture, 15 µL was aliquoted into a 384-well clear reaction plate (Fisher Scientific #4483285) in technical triplicates. The plate was centrifuged for 2 minutes at 2000 × g and 4 °C. Following centrifugation, the plate was transferred to a QuantStudio 5 real-time PCR system (ThermoFisher Scientific), where fluorescence was measured over a period of 2 hours at 37 °C. SARS-CoV-2 primers used in this work were obtained from prior work [36].

### Viral RNA extraction and RT-qPCR for SARS-CoV-2 detection (Method 14)

Nasopharyngeal swab specimens were tested for positivity and analyzed by RT-qPCR, according to a protocol established by the U.S. Centers for Disease Control and Prevention (CDC) (Ct value ≤ 40 was determined to indicate a positive sample) [37]. Viral RNA was isolated from patient samples using QIAamp Viral RNA Mini Kit (Qiagen, 52906) and RT-qPCR reactions were conducted using the QuantiNova Probe RT-PCR Kit (Qiagen, 208354) according to the manufacturer’s instructions. Briefly, 10 μL reactions were prepared in a 384-well plate format. Primers and probes for SARS-CoV-2 N and RNase P genes are listed in **Table S15** and were synthesized by IDT. All reactions were performed using the Applied Biosystems QuantStudio 5 Real-Time PCR Systems (Applied Biosystems, USA).

### Patient sample collection and ethical statement (Method 15)

Nasopharyngeal swabs were obtained from 25 suspected individuals with respiratory disease from the clinical diagnostics lab of Mount Sinai Hospital (MSH), Toronto, Canada. This study was approved by the University of Toronto research ethics board (under human ethics protocol number 39531, which permitted the screening of SARS-CoV-2 virus samples. All experiments were conducted in accordance with relevant regulations and guidelines.

### Nicked DNA Repair (Method 16)

DNA fragments corresponding to various nicked positions were prepared by annealing an equimolar mixture of three complementary fragments. The annealing was conducted at 95°C for 4 minutes in an annealing buffer [20 mM Tris-HCl (pH 7.5), 150 mM KCl, 1 mM EDTA, and 50 mM MgCl2], followed by a gradient cooling step to 4°C at a rate of 0.1°C/s. A 50 µL reaction mixture was prepared containing 1× T4 DNA ligase buffer, 1.5 U of T4 DNA ligase enzyme, 90 nM LbCas12a, 90 nM Lb gRNA, 50 U RNase inhibitor, DNA reporter 3, and 50 nM of the annealed DNA fragments. From this mixture, a 15 µL aliquot was dispensed into a clear-bottom black microplate for triplicate fluorescence readings. *Trans* cleavage activity was monitored at 37°C for 60 minutes using a BioTek plate reader, as previously described. The T4 DNA ligase enzyme was procured from Promega (Catalog #: M1801).

### miRNA-DNA Ligation (Method 17)

For miRNA-DNA ligation, two DNA fragments and the miRNA target were annealed under the same conditions described above for nicked DNA repair. The reaction components and conditions were identical to the nicked DNA repair protocol. For both reactions, the T4 DNA ligase enzyme was excluded in conditions where ligation was not required. This method allowed for precise monitoring of *trans* cleavage activity under ligation and non-ligation conditions, enabling the evaluation of nick repair and miRNA detection. For limit of detection assay, 20 μL of various miRNA-21concenration is added to the reaction mix, while every other thing remains the same.

### Mass spectrometry analysis of Cas12a-mediated reporter cleavage (Method 18)

Synthetic reporters R1 (TTATT), R3 (rUArUArUA), and R15 (GrGGrGGrG), without any modification of labelling, were obtained from Integrated DNA Technologies (IDT) with standard desalting purification. Cleavage reactions (200 μL total volume) were assembled in 1× NEBuffer r2.1 (New England Biolabs) and contained 90 nM AsCas12a, 90 nM gRNA targeting miR-21, 50 nM miR-21 RNA target, and 20 nM miR-21 DNA duplex activator. Reactions further included 6 μM reporter (R1, R3, or R15), 1 U μL ¹ murine RNase inhibitor, and MgCl adjusted to a final concentration of 31.25 mM. Samples were incubated at 37 °C for 2 h to allow Cas12a activation and *trans cleavage* of reporter substrates.

Following incubation, reactions were subjected to molecular weight filtration using 10 kDa Amicon Ultra centrifugal filters (Millipore) at 16,000g for 2 h to remove Cas12a protein, ribonucleoprotein complexes, and other high-molecular-weight components. The filtrate, containing low-molecular-weight nucleic acid fragments, was collected and directly subjected to LC–ESI-MS analysis. Control samples lacking AsCas12a were processed in parallel under identical conditions.

#### LC–ESI-MS conditions

Separation of cleavage products was performed using hydrophilic interaction liquid chromatography (HILIC) on a Thermo Scientific Ultimate 3000 UHPLC system equipped with a Waters Atlantis Premier BEH Z-HILIC column (1.7 μm, 2.1 × 100 mm) and guard column. The column was maintained at 40 °C, and the autosampler was held at 5 °C. A 10 μL sample volume was injected for each analysis. Mobile phase A consisted of 15 mM ammonium bicarbonate (pH 9.0) in water, and mobile phase B consisted of 15 mM ammonium bicarbonate (pH 9.0) in 90% acetonitrile/10% water. The flow rate was set to 0.5 mL min ¹. The gradient program was as follows: 0 min at 90% B; linear gradient to 65% B from 0–5 min; hold at 65% B from 5–6 min; return to 90% B from 6–7 min; and re-equilibration at 90% B from 7–13 min.

Mass spectrometric analysis was carried out on a Thermo Scientific Q Exactive Orbitrap mass spectrometer equipped with a heated electrospray ionization (HESI II) source, operated in negative ion mode. Source parameters were set as follows: spray voltage, 3.5 kV; capillary temperature, 325 °C; sheath gas flow rate, 40 arbitrary units; auxiliary gas, 20; sweep gas, 5; and S-lens RF level, 55.

#### Mass spectrometry acquisition and data analysis

Full MS scans were acquired over an m/z range of 200–1200 at a resolution of 70,000 (at m/z 200), with an automatic gain control (AGC) target of 3 × 10 and a maximum injection time of 200 ms. Data-dependent MS/MS (Top10) was performed at a resolution of 17,500, with an AGC target of 1 × 10, maximum injection time of 50 ms, isolation window of 0.4 m/z, and normalized collision energy of 35. Raw data were processed using Thermo Scientific Qual Browser (v4.5.445.18). Peak assignment was performed based on expected nucleotide fragment masses with a tolerance of ±5 ppm. Retention times (RT), mass-to-charge ratios (m/z), and normalized signal intensities were extracted and compiled for each reporter system. Fragment identities were confirmed by comparison with theoretical masses of expected mono- and oligonucleotide products.

## RESULTS

### Platform development

RAPID has been developed as a PAM-independent CRISPR-Cas12 technology that extends detection to both DNA and RNA, something that has not been possible with conventional Cas12 technologies (**Fig. 1A**). We started our study with a systematic investigation of the relationship between single-stranded breaks across the CRISPR-Cas12a target sequence and enzymatic *trans* cleavage activation. Conventionally, in the presence of the target sequence, Cas12a introduces a staggered dsDNA break on the dsDNA target (**supplementary Fig. S1A**), which subsequently triggers *trans* cleavage. Here, wanting to evaluate the importance of this dsDNA integrity on Cas12a function, we rationally introduced single-stranded breaks throughout the protospacer region of the target strand (**Fig. 1B**, pink triangle), (**supplementary Fig. S1B**) [38, 39]. Focusing on the sequence that interacts with the gRNA, we introduce single-stranded breaks at 24 positions starting from site between the first two PAM bases, denoted as P1, to the gRNA terminus adjacent nick, P24. (**Method 1,2**). We then tested the effect of breaks at each site on Cas12a activation by monitoring *trans* cleavage of a standard quencher-/fluorophore-labelled ssDNA. The results were striking, with single stranded breaks having a clear position-dependent effect on activation (**Fig. 1B** and **supplementary Fig. S1C**). The nuclease activity at P12 was entirely muted, with flanking regions exhibiting partial activation (semi-muted; P9, P19) and distal single stranded breaks (P2, P20) showing no inhibition.

Having identified this unexpected impact on Cas12a activation, we then performed a comprehensive experiment with six different system configurations, designated as Types I through VI, each designed to interrogate specific aspects of Cas12a’s interaction with structurally diverse ssDNA and dsDNA substrates (**Fig. 1C**). The Type I configuration consists of dsDNA, wherein the non-target strand is unperturbed, and the target strand is incrementally extended from P1 to P24 to fulfill pairing interactions between the target strand and gRNA. Our objective was to identify the minimal target strand length necessary to trigger Cas12a activation, revealing that activation commenced at P18 on the target strand for Type I (**Fig. 1D**). Conversely, in Type II, the target strand was elongated from the distal end of the PAM region (P24) towards the PAM site (P1), with activation detected only when the target strand is extended to P6 or further. Types III and IV mimic the configurations of Types I and II, respectively, but with the non-target strand omitted to assess the influence of ssDNA on Cas12a activation. Our findings showed activation when the target strand is extended beyond P21 for Type III and when the target strand is extended from P24 to P7 or further for Type IV, generally reflecting the activation patterns of their Type I and II counterparts.

Types V and VI systems display nicked ssDNA and dsDNA configurations, respectively. Type VI interrogates each nick position on a fully dsDNA target strand (**Figs. 1B,D**), while Type V is comprised solely of a ssDNA target strand, featuring single strand breaks across positions P1 to P24. In Type V, Cas12a activation was either completely or partially impaired across several segments, notably from P7 to P15 and P19 to P20, when compared to Type VI. Taken together, these patterns of deactivation, which we found to correspond to regions that are known to interact with the endonuclease domain of Cas12a [40], lends support to our hypothesis that the presence of single strand breaks perturbs protein-DNA interaction dynamics. The activation patterns across these configurations are reported in the heatmap of **Fig. 1D** and the corresponding nucleic acid design sequences are presented in **Table S1**.

With the goal of fully understanding the Type VI design, we next compared the effect of introducing single stranded breaks into the target (**supplementary Fig. S1D, Method 3**)) and non-target (**supplementary Fig. S1E, Table S2**) strands. As expected, breaks in the target strand impair *trans* cleavage activation; however, interestingly, nicks in the non-target strand had no impact of Cas12 activation, underscoring the role of target strand nicks in modulating Cas12a activation. Across the nicked regions of Type VI, full Cas12 activation was observed at positions P1–P8, P10, P15–P18, and P21–P24 (Fig. 1D). Notably, positions P9, P11, P13, P19, and P20 exhibited only partial activation, whereas nicking at positions P12 and P14 completely inhibited Cas12a *trans* cleavage activity (**supplementary Fig. S1F**).

To ensure that these observations are not a sequence-specific artifact of our model system, we expanded our study to include a novel spacer sequence, focusing on positions known to exhibit significant activation variability (P5, P8, P9, P10, P11, P12, P14, P16, P20, and P22; **supplementary Fig. S1G, Table S3**). As can be seen, the outcome was consistent with our initial observations across all six configurations (Types I to VI), supporting that the positional effects we see on Cas12 activation from single stranded breaks are generalizable and independent of the target sequence. We further compared nick-dependent activation across three distinct substrates (**Supplementary Fig. S2**; **Tables S1, S3, S4**) with the classical intact (non-nicked) substrate and found that the classical substrate consistently exhibited activation levels comparable to fully activating nick positions (e.g., P5) across all targets.

To further understand the mechanism of Cas12a inhibition by single stranded breaks, we explored the local protein environments at the nicked positions using published Cas12a crystal structures (PDB: 8Y03) of Cas12a in complex with gRNA and dsDNA [40–42]. With the hypothesis that single stranded breaks at certain positions may alter protein-DNA interactions and therefore affect Cas12a activation, we evaluated the structure at sites where we had seen clear inhibition of activity (**Fig. 1D**). Interestingly, we found the region most affected by single strand breaks (P9 to P14) aligns with the endonuclease domains (RuvC and BH), which are critical for both *cis* and *trans* cleavage [2, 40, 41]. **Table S5** summarizes the rest of the nick positions rendering substantially diminished activity of Cas12a, along with their interacting amino acids (within 3.5 angstroms). Similarly, we found that P19 and P20, which exhibit semi-muted activation upon nick introduction, primarily interact with amino acids in the REC2 domain, which is involved in gRNA binding to the protein [42].

To examine if there might also be an intrinsic nucleic acid stability component underlying nick-dependent Cas12a activation, we performed thermal melting (Tm) analysis on representative substrates, including a non-nicked control and nicked substrates (**Supplementary Fig. S3**) corresponding to complete activation (P5, P8), partial activation (P9, P20), and muted activation (P12, P14), using a CFX96 Real-Time System (BioRad). With the exception of one position (P20), the thermal stability of the annealed duplexes showed a clear correlation with trans-cleavage activity, with higher melting temperatures associated with stronger activation and lower Tm values corresponding to reduced or absent activity (**Supplementary Fig. S3, Table S1, Method 4**). These results indicate that protospacer nicking modulates duplex stability in a position-dependent manner, which in turn influences Cas12a activation. One possible explanation is that reduced duplex stability at certain nick positions leads to increased local strand dynamics or transient fraying, thereby impairing stable target engagement and limiting efficient activation of the enzyme.

While the precise biochemical implications of these alterations remain to be fully explored, the collective data and structural observations provide new insight into the mechanisms regulating Cas12a activation and highlight the role of protospacer integrity. Importantly, these findings also present a novel perspective on how the CRISPR-Cas12 system can be manipulated at the molecular level, introducing an opportunity to exploit these findings to advance the Cas12a diagnostic platform.

### Sensor development

With these new design considerations in-hand, we set out to leverage our findings for the development of an updated Cas12a-based detection platform (**Method 5-6**). To begin sensor development, we assessed Cas12a activation across the 24 nicked positions (P1 to P24, **Fig. 1D**) for configuration Types I through VI to identify an optimal design (nicking point). The Type VI system containing a single stranded break at P8 emerged as a promising candidate as this site produced complete activation of the Type VI system and negligible activation of Types I to V. This unique configuration enables an ‘AND logic’ sensing mechanism within our RAPID system; wherein full system assembly is required for Cas12a activation. Specifically, *trans* cleavage is triggered only when all nucleic acid components are present. Consequently, the Type VI configuration provided the foundation for our RAPID sensors.

The RAPID Type VI system has the potential to provide a highly flexible diagnostics platform as enzyme activation can be triggered by multimodal nucleic acid formats, including ssDNA, mRNA and miRNA, as well its conventional dsDNA target format (**Fig. 2A**, light blue). CRISPR gRNAs (red) were engineered for sequence-specific binding of these diverse nucleic acid types, which, in the presence of a target sequence, leads to Cas12a *trans* cleavage of a fluorescence reporter. Another critical component of RAPID is the PAM duplex (Fig. 2A, dark blue strands), a dsDNA sequence consisting of a PAM (orange) and neighbouring ssDNA toehold domain (dark blue, bottom right). The PAM sequences are introduced in *trans* configuration, meaning PAM sites are separate from the target nucleic acid, relieving the PAM sequence requirement from the diagnostic design process. Crucially, when short target ssDNA or miRNA bind to the toehold domain, a pseudo-nick is created at position P8 in the Type VI system, enabling Cas12a activation (Mode I, **Fig. 2A(I)**). Similarly, longer ssDNA or RNA target sequences can also trigger system activation and form a pseudo-nick at P8 (Mode II, **Fig. 2A(II)**).

**Fig. 2:**
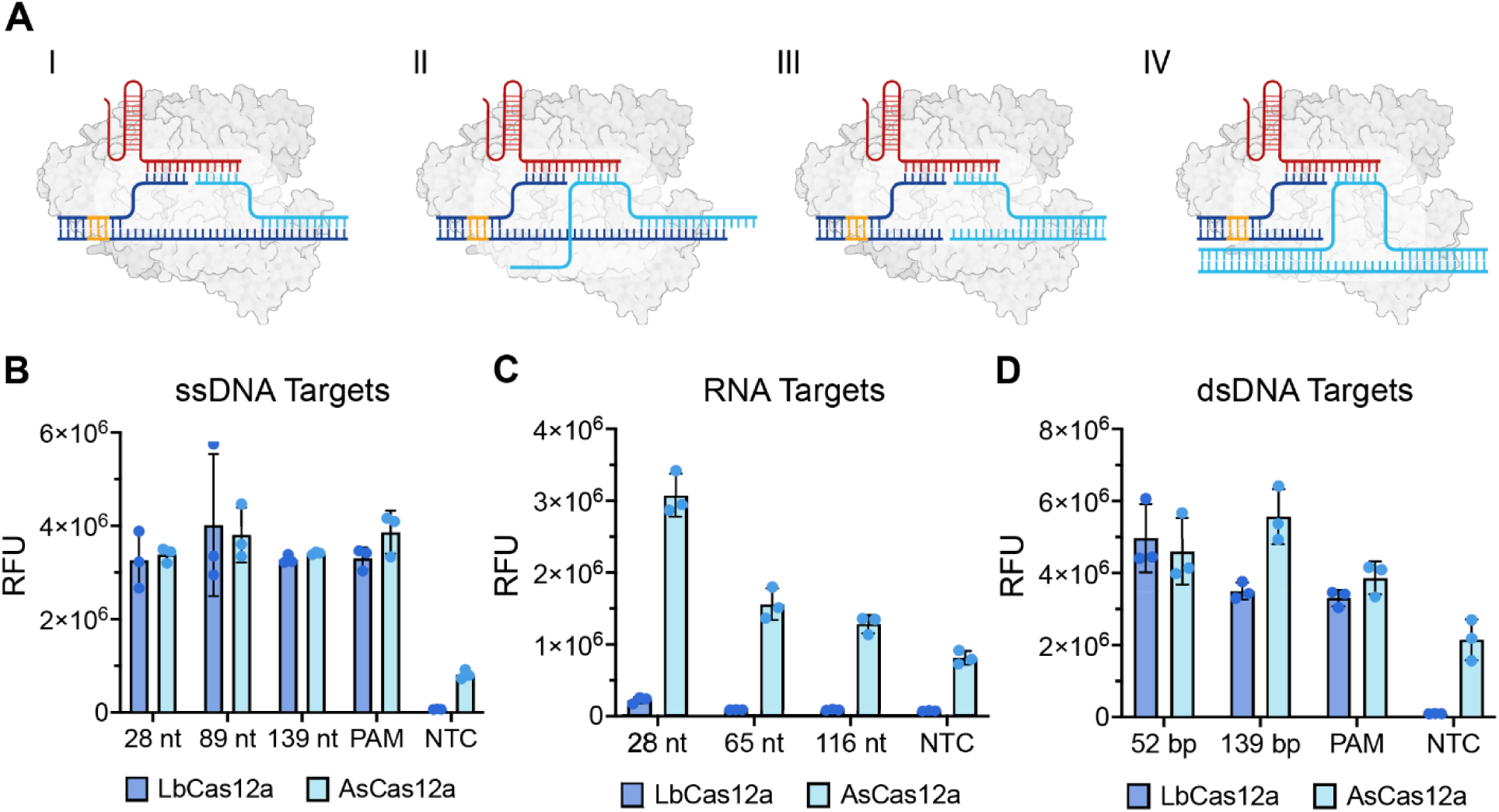
System configurations of RAPID for nucleic acid detection. **(A)** Configurations showing PAM-independent detection across various nucleic acid types: short and long single-stranded nucleic acids, and short (blunt-ended) and long double-stranded DNA (dsDNA). Dark blue represents the PAM duplex, red the gRNA, cyan the target strand, and orange the *trans* PAM. **(B)** Performance of RAPID in detecting various lengths of target ssDNA (28-nt, 89-nt, 139-nt) compared to PAM-containing dsDNA. The non-target controls (NTC) display higher a background signal with AsCas12a and low background with LbCas12a. **(C)** Detection capabilities of RAPID for target RNA lengths of 28-nt, 65-nt, and 116-nt, demonstrating better performance with shorter RNA. AsCas12a exhibits higher background noise with NTC compared to LbCas12a. **(D)** Use of RAPID for PAM-independent detection of target dsDNA, including blunt-ended dsDNA (52-bp) and longer dsDNA (139-bp), compared to PAM-containing dsDNA. RAPID achieves PAM-independent activation via protospacer nick-position tuning and a trans-supplied PAM duplex, contrasting with PAM-independent activator-generation strategies based on toehold strand displacement or bubble substrates. The NTC for AsCas12a shows high background noise, in contrast to LbCas12a, which shows low background noise. (**Method 6**). All nucleic acid sequences are presented in **Table S6**. n=3 technical replicates; bars represent the arithmetic mean ± SD. RFU is relative fluorescence unit.

For the detection of PAM-independent dsDNA, we have designed a universal PAM duplex that lacks a toehold domain (Mode III, **Fig. 2A(III)**). In this configuration, the PAM sequence is again introduced in *trans* and is structurally distinct from the dsDNA trigger. Here, we present two use cases for this dsDNA design. In the first, the gRNA-Cas12a complex binds to both the universal PAM duplex and a blunt-ended PAM-independent dsDNA, leading to activation (**Fig. 2A(III)**), and a second, where a segment of the gRNA binds to the universal PAM while another segment binds to an extended PAM-independent dsDNA target, thereby activating Cas12a (Mode IV, **Fig. 2A(IV)**). Importantly, the RAPID system directly detects a dsDNA target that does not contain a PAM, in contrast to a recent PAM-free dsDNA platform [33], where the dsDNA target must first undergo transcription to generate RNA amplicons for downstream detection [33].

### Nucleic acid sensing with RAPID

We tested RAPID using two commercially available Cas12a orthologs, specifically *Acidaminococcus* sp. (AsCas12a) and *Lachnospiraceae bacterium* (LbCas12a). Our initial experiments focused on enhancing Cas12a detection sensitivity for ssDNA using RAPID by pairing ssDNA with a sticky end dsDNA containing the PAM duplex, as depicted in **Fig. 2A** (I and II), **Method 6**. Notably, the dsDNA configuration enhances the *trans* cleavage signal of CRISPR-Cas12, likely due to its greater stability when complexed with the Cas12-gRNA, in contrast to the weaker signal observed with ssDNA [2, 43]. This property makes our Type VI system particularly interesting, as it mimics the dsDNA configuration while exhibiting an enhanced *trans* cleavage fluorescence signal. We experimented with short ssDNA target strands (light blue) of 28 nucleotides (nt), corresponding to **Fig. 2A(I)**, and longer strands of 89 and 139 nt, corresponding to **Fig. 2A(II)**. Utilizing PAM-containing dsDNA as a benchmark, we observed that the *trans* cleavage signals from all ssDNA triggers were comparable to those from PAM-containing dsDNA for LbCas12a and AsCas12a (**Fig. 2B**). Moreover, we observed that AsCas12a exhibited a higher no template control (NTC) background signal than that of LbCas12a, consistent with findings from previous studies [13, 24].

Extending this analysis to RNA targets, we discovered differential *trans* cleavage activation thresholds between the two Cas12a variants. RNA target sequences activated *trans* cleavage remarkably well when complexed with AsCas12a, while *trans* cleavage was significantly muted with LbCas12a (**Fig. 2C**). The target RNA lengths tested included a 28-nt sequence, akin to the configuration in **Fig. 2A(I)**, and longer sequences of 65 and 116-nt, paralleling the setup in **Fig. 2A(II)**. Notably, the shorter target RNA (28-nt miRNA) was more effective at triggering *trans* cleavage than longer RNA counterparts.

Further exploration of the RAPID system revealed efficient detection of PAM-independent dsDNA. We first tested a titration of the universal PAM duplex concentration in the detection of both blunt-ended and longer dsDNAs, finding the optimal concentration to be 200 nM (**supplementary Fig. S4A,B**). Using this PAM duplex concentration, we next exposed the RAPID system (**Fig. 2A(III)**) to 52 bp PAM-independent target dsDNA sequence, finding PAM-independent sequences could match or exceed activation of the PAM-containing dsDNA (∼10 bp) for both LbCas12a and AsCas12a (**Fig. 2D** for 52 bp, **supplementary Fig. S4C**). We similarly assessed detection of PAM-independent dsDNA at an arbitrary location in a 139 bp target, mirroring the configuration in Fig. 2A(IV), and again found activation of both Cas12a orthologs on par or better than PAM-containing dsDNA (**Fig. 2D** for 139 bp, **supplementary Fig. S4C**).

These results suggest that Cas12a activation is primarily governed by PAM-proximal structural elements, with reduced dependence on distal non-target strand integrity, providing opportunities for further optimization of minimal PAM duplex designs, which is currently ongoing [44]. We propose that gRNA annealing to the PAM-independent dsDNA may proceed via either helicase-assisted unwinding, consistent with canonical Cas12 mechanisms [2], or through a helicase-independent pathway, as previously reported [45, 46]. To further demonstrate practical applicability, Mode IV (**Fig. 2A(IV)**) was further validated using amplified monkey pox virus (MPXV) dsDNA targets in comparison to the traditional CRISPR/Cas system – DETECTR (**Supplementary Fig. S5, Table S10, Method** 7), confirming robust detection of internally located sequences relevant to clinical diagnostics, with comparable signal intensity.

### *Trans* cleavage activation with RAPID

Conventionally, Cas12a systems are considered DNase systems designed to target DNA and enable DNA *trans* cleavage, as demonstrated by platforms like DETECTR and HOLMES [2, 24]. Therefore, Cas12a platforms have historically been developed for ssDNA substrates, with RNA-based *trans* cleavage exhibiting lower efficiency in traditional Cas12a applications [47]. Here, with the aim of expanding Cas12a’s role in diagnostics, we investigate whether other nucleic acid substrates can be harnessed for Cas12a *trans* cleavage (**Fig. 3A**). We began by designing a series of sequences to test *trans* cleavage efficiency with substrates comprising DNA, RNA, and chimeric (e.g. mixed sequences that contain both deoxyribo- and ribo-nucleotides) polymers (Reporters 1 to 16 in **Table S7**).

**Fig. 3:**
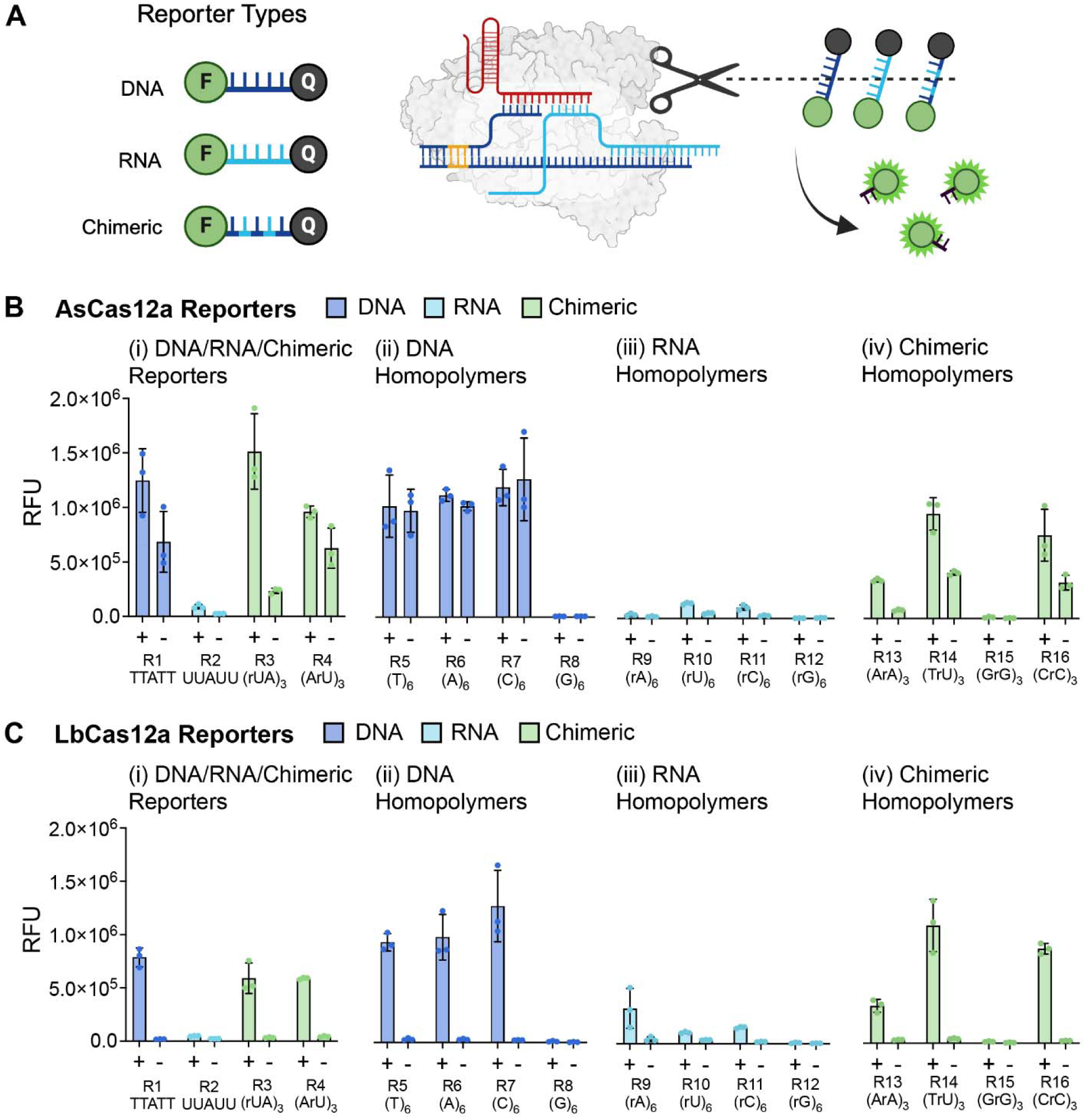
Optimization of reporter sequences for the RAPID system. **(A)** Schematic of the RAPID system configured to cleave DNA, RNA, and chimeric reporters modified with fluorophore-quenching pairs. **(B)** Performance of RAPID with AsCas12a. **(C)** Performance of RAPID with LbCas12a. The system includes 16 reporters, divided into four categories: DNA/RNA/chimeric polymers (R1 to R4), DNA homopolymers (R5 to R8), RNA homopolymers (R9 to R12), and chimeric homopolymers (R13 to R16). Reporters are annotated with repeat numbers indicating base repetition (e.g., (T)_6_ = TTTTTT, (rUA)_3_ = rUArUArUA). (**Method 8**). All reporters are modified with 56-FAM and 31ABkFQ. Results are expressed as mean ± SD (n = 3) in relative fluorescence units (RFU). n=3 technical replicates; bars represent the arithmetic mean ± SD, all sequences can be found in **Table S7**.

We first applied RAPID to *trans* cleave the traditional ssDNA sequence (R1: TTATT) using AsCas12a and LbCas12a (**Fig. 3B, C**). Consistent with conventional Cas12a systems [13], we noted a high non-specific signal in the absence of target sequence (fluorescence background) with AsCas12a compared to LbCas12a (**Fig. 3B(i), C(i)**). We next applied RAPID to *trans* cleave an RNA sequence (R2: rUrUrArUrU, (**Fig. 3B(i)**). The cleavage activity was stronger for AsCas12a than for LbCas12a, but the signal for both Cas12a orthologs was largely muted. Subsequently, we tested modified RNA sequences by introducing DNA bases (R3: (rUA)_3_ and R4: (ArU)_3_).

Surprisingly, the *trans* cleavage signal was stronger for these chimeric sequences than for the RNA-based sequences for both AsCas12a and LbCas12a (**Fig. 3B(i)** and **Fig. 3C(i)**). This provided significant reduction of the background signal for AsCas12a with R3 (**Fig. 3B(i)** for R3), in comparison to conventional DNA substrate R1 (**Fig. 3B(i)** for R1). Importantly, in addition to minimizing background interference, this is the first instance where chimeric reporters have been shown to match the efficiency of DNA-based reporters in Cas12a-based systems.

Building on these preliminary findings, we screened Cas12a RAPID DNase activity with a series of ssDNA homopolymer reporters in the presence and absence of target sequence [R5: (T)_6_, R6: (A)_6_, R7: (C)_6_, and R8: (G)_6_]. As before, ssDNA homopolymers yielded very high background signals for R5, R6, and R7 with AsCas12a, compared to the high signal and low background observed for LbCas12a. Negligible *trans* cleavage was observed for poly G (R8) in either Cas12a ortholog (**Fig. 3B(ii)** and **Fig. 3C(ii)**), which we anticipate is primarily due to photoinduced electron transfer from guanine, the most easily oxidized nucleobase, to the excited fluorophore [48, 49].

Next, we tested RAPID with an expanded series of RNA homopolymer reporters [R9: (rA)_6_, R10: (rU)_6_, R11: (rC)_6_, and R12: (rG)_6_)] to assess *trans* cleavage RNase activity. As expected for Cas12a, we saw a generally weak response, with no RNase activity observed for R9 and R12 in AsCas12a and for R12 in LbCas12a. Limited activity was seen for R10 and R11 with AsCas12a and for R9, R10, and R11 with LbCas12a (**Fig. 3B(iii)** and **Fig. 3C(iii)**). Finally, we applied RAPID to *trans* cleave chimeric homopolymers [R13: (ArA)_3_, R14: (TrU)_3_, R15: (GrG)_3_, and R16: (CrC)_3_]. The results showed good to excellent *trans* cleavage effects for R13, R14, and R16 with both Cas12a orthologs, expanding the chimeric reporter toolbox, while R15 was not cleaved by either Cas12 enzyme (**Fig. 3B(iv)** and **Fig. 3C(iv)**), perhaps due to the guanine’s photoinduced electron transfer [50]. All sequences are listed in **Table S7**, and kinetic plots are available in **supplementary Fig. S6A-C**.

The results showed that selected RNA-based reporters could be leveraged for diagnostics, albeit weakly. However, chimeric probes performed comparable to some ssDNA sequences, with enhanced performance using chimeric probe R3, which significantly decreased the non-specific background signal of AsCas12a (**Fig. 3B**), indicating that AsCas12a possesses a broader substrate tolerance for these variants than LbCas12a. Additionally, it was noted that the background signal from R1, which is the traditional reporter substrate for Cas12a, was significantly higher than R3 – the novel reporter for AsCas12a, aligning with observations from traditional CRISPR-Cas12a systems [24, 51].

Furthermore, ssDNA sequences demonstrated superior *trans* cleavage activity compared to their RNA counterparts when interacting with LbCas12a (**Fig. 3C (ii & iii)**). This differential activity highlights the inherent preference of LbCas12a for ssDNA substrates within the RAPID system. In comparison to AsCas12a, LbCas12a exhibited only slightly reduced ability for cleaving the chimeric probes, R3 and R4, and excellent cleavage of chimeric reporters R14 and R16, respectively, suggesting flexible *trans* cleavage substrate specificity within the RAPID platform. All the kinetic plots are presented in **supplementary Fig. S6A-C**. For LbCas12a, testing non-canonical reporter sequences also provided a productive strategy to improve the signal-to-noise ratio.

Wanting to benchmark the performance of chimeric reporters, we then integrated CRISPR-Cas12a with an upstream recombinase polymerase amplification (RPA) reaction for a PAM-containing dsDNA target (synthetic HPV-18 – **Table S8**) and chimeric reporter (R3: (rUA)_3_). We were pleased to find both LbCas12a and AsCas12a orthologs, in combination with RPA, provided sensitivity down to 1 aM (**supplementary Fig. S7A,B, Method 8**). Control reactions without RPA were run in parallel to identify the detection limit of the Cas12a orthologs alone (10 pM target concentration). Reporter R3 exhibited comparable fluorescence output to canonical reporters while maintaining reduced background signal. Importantly, the results highlight the broad applicability of chimeric reporter R3 in both RAPID and conventional Cas12a detection systems.

### Electrospray ionization mass spectrometry (ESI-MS) evidence for AsCas12a *trans* cleavage of RAPID reporters

To directly validate that RAPID fluorescence outputs reflect bona fide Cas12a-mediated reporter hydrolysis, we analysed post-reaction mixtures by ESI–MS for three representative reporters (**Supplementary Fig. S8, Method 18, Table 9**) alongside matched negative controls processed without AsCas12a (**Supplementary Fig. S9, Method 18, Table 9**). For the canonical ssDNA reporter R1 (5′-TTATT-3′; a widely used DETECTR-type motif), AsCas12a treatment yielded smooth, high-intensity peaks corresponding to short nucleotide products: m/z 321.049 (T; RT 3.49 s; 1.35×10□), m/z 330.061 (A; RT 3.59 s; 1.40×10□), and m/z 625.095 (TT; RT 3.77 s; 3.04×10□) indicating cleavage, whereas longer candidate intermediates (TA, TTA) were not detected under these conditions. In the enzyme-free control, none of these fragment peaks were observed, supporting that detectable R1 fragmentation requires AsCas12a activity rather than spontaneous hydrolysis or buffer-dependent degradation [2].

The chimeric RNA/DNA reporter R3 (5′-rU-A-rU-A-rU-A-3′) showed an analogous, enzyme-dependent fragmentation profile (**Supplementary S8B**), with detected products limited to rU (m/z 323.029; RT 4.18 s; 5.29×10□), A (m/z 330.061; RT 3.92 s; 1.65×10□), and the dinucleotide rUA (m/z 636.086; RT 4.23 s; 1.92×10□), while longer fragments (for example rUArU, ArUA) were not detected and no fragment peaks were present in the no-enzyme control. Taken together, the restriction of detectable products to mono- and dinucleotide species for both R1 and R3, combined with the absence of longer intermediates, is consistent with rapid, multiple-turnover Cas12a trans-cleavage in which initial internal scission events generate transient oligomers that are further cleaved, collapsing the product distribution to short, chemically well-resolved fragments, providing ESI–MS support for the reporter degradation that underpins DETECTR-like fluorescence readouts [2]. In contrast, the poly(G)-rich reporter R15 (GrGGrGGrG) produced no detectable cleavage fragments even in AsCas12a-treated samples (**Supplementary Fig. S8C**), aligning with prior quantitative studies showing that Cas12a trans-cleavage is strongly reporter-sequence dependent and that poly(G) reporters are refractory (or nearly refractory) to *trans* cleavage [52]. This reporter-dependent fragmentation pattern provides a robust, orthogonal MS-level rationale for the observed fluorescence behaviour (strong signal for TTATT and rUA-containing reporters, and absent signal for poly(G) reporters) and supports the prioritisation of chimeric reporter designs that preserve strong cleavage products while minimising nonspecific background.

### Ligation-induced DNA repair with RAPID

Having introduced a strand break into the Cas12a DNA substrate, here we test whether *in situ* ligation of a DNA nick can restore *trans* cleavage activity (**supplementary Fig. S10A**). To test this concept, we introduce a duplex probe phosphorylated at the 5′-end of the target strand to enhance ligation efficiency (**Fig. 4A**). We then selected four distinct nick positions—P5, P8, P9, and P12— corresponding to activated (P5, P8), semi-muted (P9), or completely muted (P12) RAPID configurations (**Fig. 1D**). For configurations where the strand break impairs *trans* cleavage (P9, P12), the addition of ligase to one-pot RAPID reactions completely restores activity (**Fig. 4B** – green bars, **supplementary Fig. S10B**). As expected, both unimpaired or active RAPID configurations (P5, P8) do not benefit from the addition of ligase. The activity of these reactions was benchmarked against RAPID reactions with a fully intact PAM-containing dsDNA substrate and a no-template control (NTC).

**Fig. 4:**
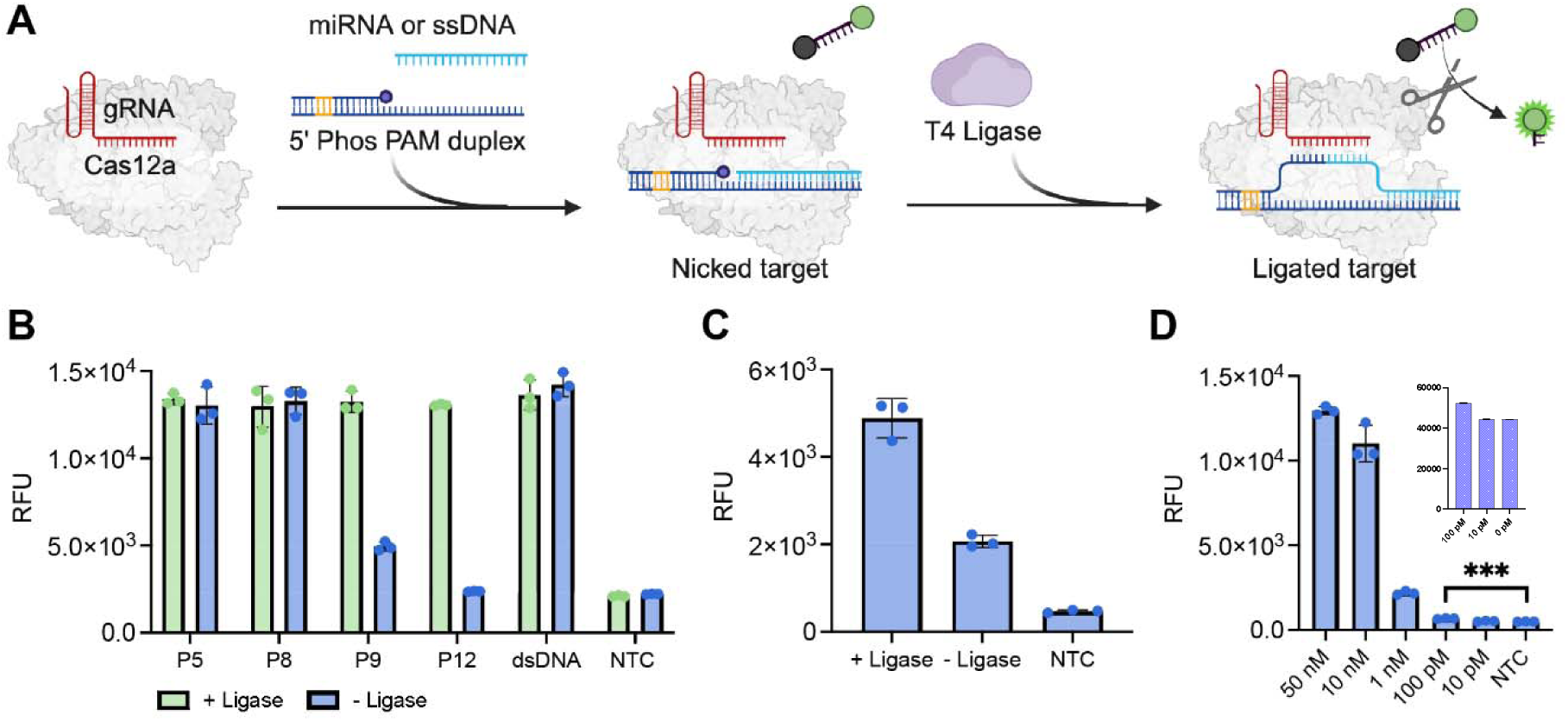
Ligation-Induced *Trans* Cleavage. (A) Schematic representation of nicked DNA repair using T4 DNA ligase. The duplex probe contains a 5’-end that is phosphorylated at the nicked point, enabling repair in the presence of ligase, which restores the *trans* cleavage signal in one-pot reaction format. (B) Results depicting ligation activity of nicked DNA at various positions. The green bar is post-ligation, while the blue bar is before ligation. Positions P5 and P8 demonstrate complete activation regardless of T4 ligase presence, corresponding to the primary discovery from P1 to P20 on the heat map in Fig. 1. Position P9 exhibits partial activation without ligase, which is fully restored upon ligase addition. At position P12, no activation occurs without ligase; however, the signal is recovered with ligase. Nickless dsDNA with a PAM serves as a control template, and NTC represents the no-target control. (**Method 16**). (C) Demonstration of miRNA ligation to a DNA substrate, forming a chimeric DNA-RNA configuration in a single reaction. As position P8 showed promising results for detecting RNA (Fig. 2 driven by the addition of MgCl_2_,), here, without additional MgCl_2_ and with the presence of the miRNA target and T4 ligase, a strong signal is generated, whereas low signal is observed without T4 ligase, and no signal in the negative control lacking miRNA. (**Method 17**). (D) Testing the limit of detection for miRNA-21 using ligation-induced *trans* cleavage, achieving significant detection at concentrations as low as 100 pM. (See inset; **Method 17**). Symbols (+) and (-) denote the presence and absence of ligase enzyme, respectively. NTC is no target control (negative control). Data represent mean ± standard deviation from n = 3 technical replicates at 60 min. All sequences are contained in **Table S9**.

Beyond demonstrating a rescue of RAPID activity, we next considered the potential of ligase to enhance the detection of miRNAs with RAPID. By joining the target miRNA to a DNA substrate, a chimeric DNA–RNA substrate is formed to activate CRISPR-Cas12-mediated *trans* cleavage. We were excited to see that the addition of ligase enzyme (1.5 U) to RAPID detection of miRNA doubles the *trans* cleavage activity compared to no ligase addition (**Fig. 4C**). The resulting system, without pre-amplification, can detect miRNA-21 down to 100 pM with chimeric reporter 3 (R3) (**Fig. 4D** inset**, Method 17, Table S11**). Compared to existing miRNA detection strategies, this ligation-based approach provides enhanced flexibility and superior sensitivity.

### miRNA detection with RAPID

miRNAs are a category of small, single-stranded, non-protein-coding RNAs with a length of approximately 19-23 nucleotides, and have been widely reported as biomarkers of several cancers, diabetes, immune dysregulation, and other disease states [53, 54]. The direct detection of these biomarkers using conventional Cas12a systems is challenging due to incompatible reaction buffers with low salt concentrations, the requirement for a PAM, and high background signal [24].

Leveraging this proof-of-concept, we next optimized the RAPID system components for the direct detection of miRNAs (**Fig. 5A**). This began with an evaluation of the MgCl_2_ concentration, which we found was optimal at 31 mM. (**supplementary Fig. S11A**). Next, titrations of duplex DNA, AsCas12a enzyme, and gRNA concentrations were screened, finding optimal performance at approximately 20 nM, 90 nM, and 90 nM, respectively (**supplementary Fig. S11B-D**; heatmap - **supplementary Fig. S11E**). We also found that the addition of ∼6% PEG-8000 as a crowding agent enhances the *trans* cleavage signal (**supplementary Fig. S12**).

**Fig. 5:**
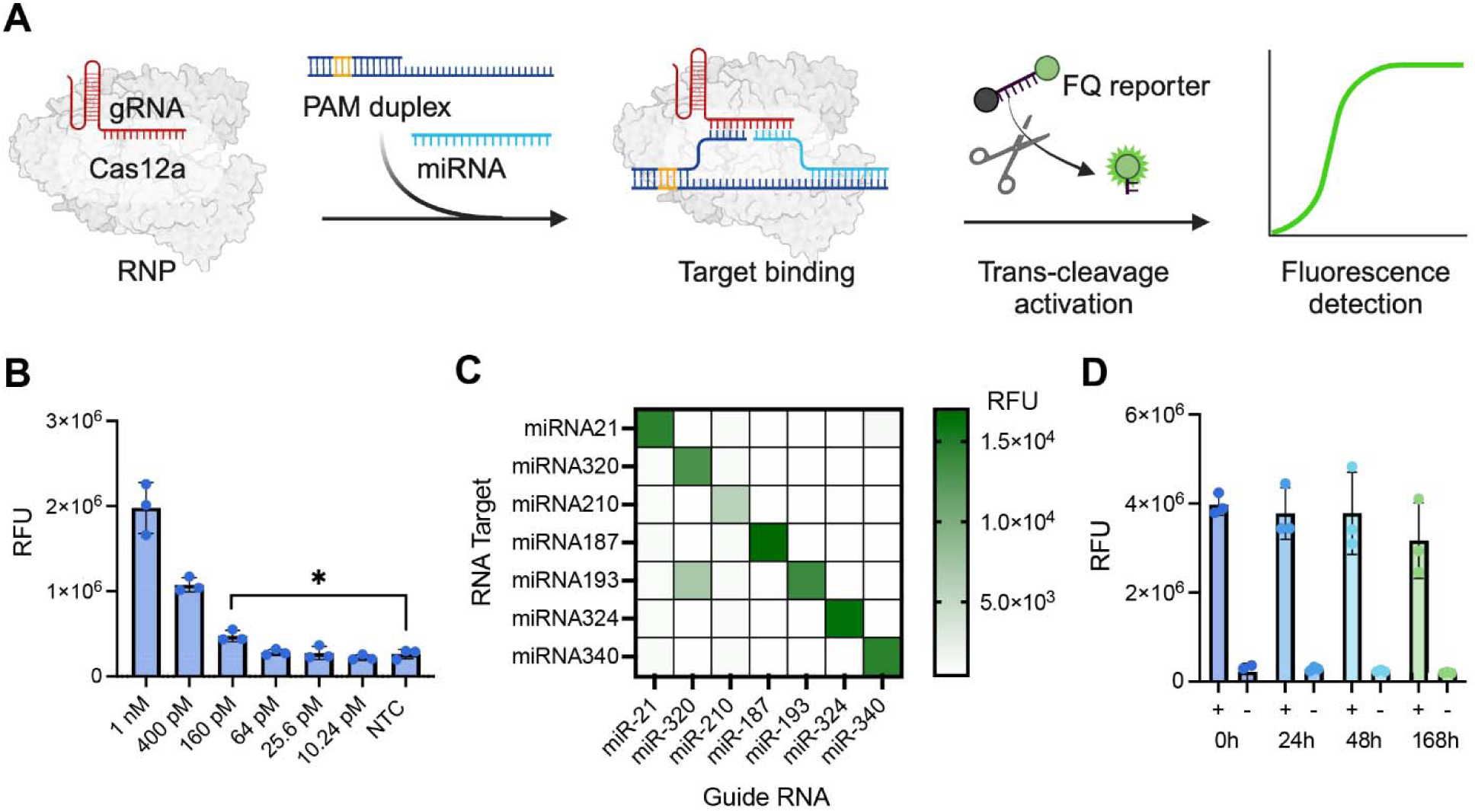
Utilizing RAPID for miRNA Diagnostics. **(A)** Schematic of the RNA detection setup using RAPID. **(B)** Limit of detection for miRNA-21 using RAPID, achieving sensitivity down to 97 pM (**supplementary Fig. S13**). **(C)** Heatmap illustrating orthogonality testing of RAPID for seven miRNAs: miRNA-21, miRNA-320, miRNA-210, miRNA-180, miRNA-193, miRNA-324, and miRNA-340. **(D)** Stability testing of freeze-dried RAPID components over 7 days, demonstrating good stability and field deployability. (**Method 9-10).** All incubation time is 1 h. Sequences in **Table S13**. n=3 technical replicates; bars represent the arithmetic mean ± SD. NTC refers to no template control.

With the optimized conditions in place, we proceeded with the development of a RAPID assay for miRNA-21 (**Supplementary Fig. S13A**), chosen for its significance in breast cancer prognosis [53]. (**Method 9**). Beginning with the use of RAPID alone, we found a detection limit of 97 pM, calculated using linear regression (**Method 9, Fig. 5B** and **supplementary Fig. S13B**), which provides comparable performance to Cas13 RNA-based miRNA detection [55, 56]. This enhancement is particularly notable given that RAPID covers a 16 nt miRNA sequence, potentially translating to greater specificity, compared to earlier miRNA targeting Cas12 systems that cover ≤12 nt and report a detection limit within the nM range [23, 24]. Additionally, we tested RAPID’s specificity for seven miRNAs: miRNAs- 320, 210, 187, 193, 324, and 340, which are biomarkers for other conditions, including cancer prognosis [53] —and confirmed orthogonality in detection (**Fig. 5C** and **supplementary Fig. S14A, Table S12-13**). Importantly, *in situ* ligation was employed to demonstrate feasibility and signal enhancement, however, subsequent miRNA experiments focused on baseline RAPID platform performance.

The isothermal nature of the RAPID platform positions it well as a tool for decentralized diagnostics, so we evaluated the performance of freeze-dried reagents for the potential distribution of test kits without a cold chain (**Method 10**). As can be seen from endpoint reads (2 h), we found that the system demonstrated robust room temperature stability for up to one week (168 h) from the time of freeze-drying (**Fig. 5D** and **supplementary Fig. S14B**). Reactions containing all the RAPID components, including AsCas12a and R3 reporter, were flash frozen in liquid nitrogen and freeze-dried overnight in a Labconco FreeZone 6 Liter −84 °C Console Freeze Dryer and vacuum packed with desiccant for storage.

### Discrimination of single and two-point mutations with RAPID

The capacity to identify point mutations in target sequences can provide an important advantage to diagnostics, enabling the detection of viral variants, the presence of drug resistance, or the detection of oncogenic mutations, among many other applications [57]. Yet, detecting such subtle differences is currently a challenge [33]. To investigate the potential of the RAPID platform to provide single nucleotide polymorphism (SNP) detection, we screened ssDNA sequences containing point mutations and paired two-base mismatches across the target binding regions (1-16, **Fig. 6**). The sequences, including the target regions (blue) and the mismatches (red), are detailed in **Tables S14-15**, respectively, facilitating a direct comparison between the mutated sequences and their wildtype (WT) counterparts.

**Fig. 6:**
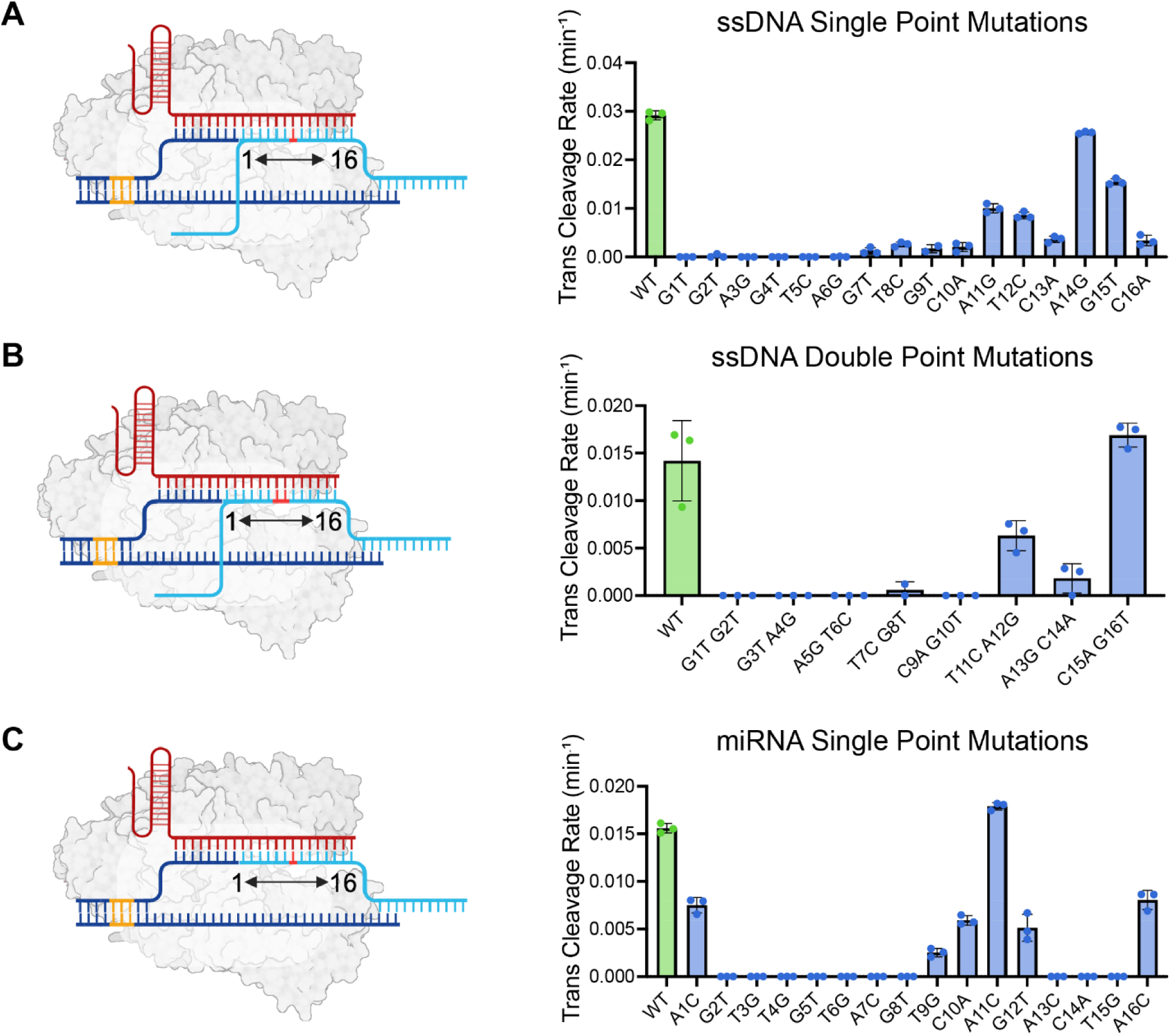
Evaluating the Ability of RAPID to Detect Mismatches in DNA and RNA. **(A) Detection of Single-Point Mutations in ssDNA:** The left panel shows schematics of the point mutation, and the right panel displays the *trans* cleavage rate. **(B) Detection of Two-Base Mismatches in ssDNA:** Schematics of the mismatches are on the left, with corresponding detection results on the right. **(C) Detection of Single-Point Mutation in miRNA-21:** RAPID targets a 16-nt region within the protospacer of miRNA-21, with the remainder binding to duplex DNA. (**Method 11**) Green bars represent the wild type; blue bars represent mutants and non-target controls (NTCs). Results are expressed as *trans* cleavage rate per min (for analysis, see **Method 11**). Bar numbering corresponds to mutation sites (e.g., G1T indicates guanine mutated to thymine at position 1, nearest to the *trans* PAM). n=3 technical replicates; bars represent the arithmetic mean ± SD. Final DNA concentration is 2 μM, while miRNA is 50 nM. *Trans* cleavage rate is analysed within 2 h. All data was collected as a time series. The *trans* cleavage rate data was analysed within 2 h (**Method 11**), while the 60-minute end point data is presented in **Supplementary Fig. S15A**.

Tracking the rate of *trans* cleavage within 2 h, RAPID demonstrated a marked ability to distinguish point mutations located proximal to the *trans* PAM region for ssDNA (positions #1 to 10) termed “RAPID seed region” (**Fig. 6A, Supplementary Fig. S15A** for endpoint measurement in RFU, **Method 11**). However, the detection efficiency waned for mutations situated more distal from the *trans* PAM (positions #11 to 16). Next, we introduced paired two-base DNA mismatches within the target sequences. As above, we saw strong selectivity in the RAPID seed region within a 120 min reaction time (**Fig. 6B, Supplementary Fig. S15B**). The capacity for SNP detection in combination with PAM-independent detection introduces an opportunity to rationally design detection schemes to place target mutations within the region of high selectivity for both SNP and paired mismatches. Importantly, these extended regions of specificity for SNPs and two-paired mismatches are not possible using the conventional CRISPR-Cas12a system containing ssDNA [2, 33].

The small size of miRNAs, and their high sequence similarities, makes the discrimination of closely related targets especially challenging for CRISPR diagnostics [58]. These small, single-stranded, non-protein-coding RNAs are proving to be biomarkers of several cancers, diabetes, immune dysregulation, and other disease states [53, 54]. Here, with the goal of demonstrating the potential of RAPID to address this need, we tested point mutations across synthetic miRNA-21 variants (**Fig 6C, Supplementary Fig. S15C**) using AsCas12a and the R3 chimeric reporter. Interestingly, RAPID provided SNP selectivity in proximal (2-8) and distal (13-15) regions of the miRNA, highlighting differences in RAPID’s interactions with miRNA and ssDNA and suggesting Cas12a activation requirements are altered when accommodating non-canonical RNA substrates.

RAPID’s sensitivity to nucleotide alterations (SNPs) parallels findings from SAHARA employing Cas12a systems for RNA detection [23]. Importantly, RAPID covers 16-nt of the target sequence and is specific over a longer region compared to the recent studies from SAHARA, which was limited to detection of 12-nt of a target sequence with limited specificity [23].

### RAPID coupled with RT-LAMP for SARS-CoV-2 RNA detection in clinical samples

To test the potential for RAPID to serve as an isothermal diagnostic alternative to gold standard RT-qPCR, we developed a RAPID assay for the SARS-CoV-2 nucleocapsid gene. Here, demonstrating the flexibility of RAPID, we replaced RPA with reverse transcriptase loop-mediated isothermal amplification RT-LAMP; (**Fig. 7A, Method 12**). To establish the protocol, we first tested RAPID separately with synthetic DNA strands that mimic the two loops of the anticipated ssDNA LAMP dumbbell amplicon structure (**Fig. 7A** and **supplementary Fig. S16A**), which forms a full dsDNA target when bound to the RAPID duplex probe for enhanced *trans* cleavage activity. The results (**supplementary Fig. S16A**) revealed that designing the system to target both ssDNA LAMP dumbbells could significantly enhance detection. With this result in hand, both loops were targeted, taking advantage for the first time, of RAPID’s split protospacer enabling converting ssDNA detection to a dsDNA format for enhanced *trans* cleavage performance.

**Fig. 7:**
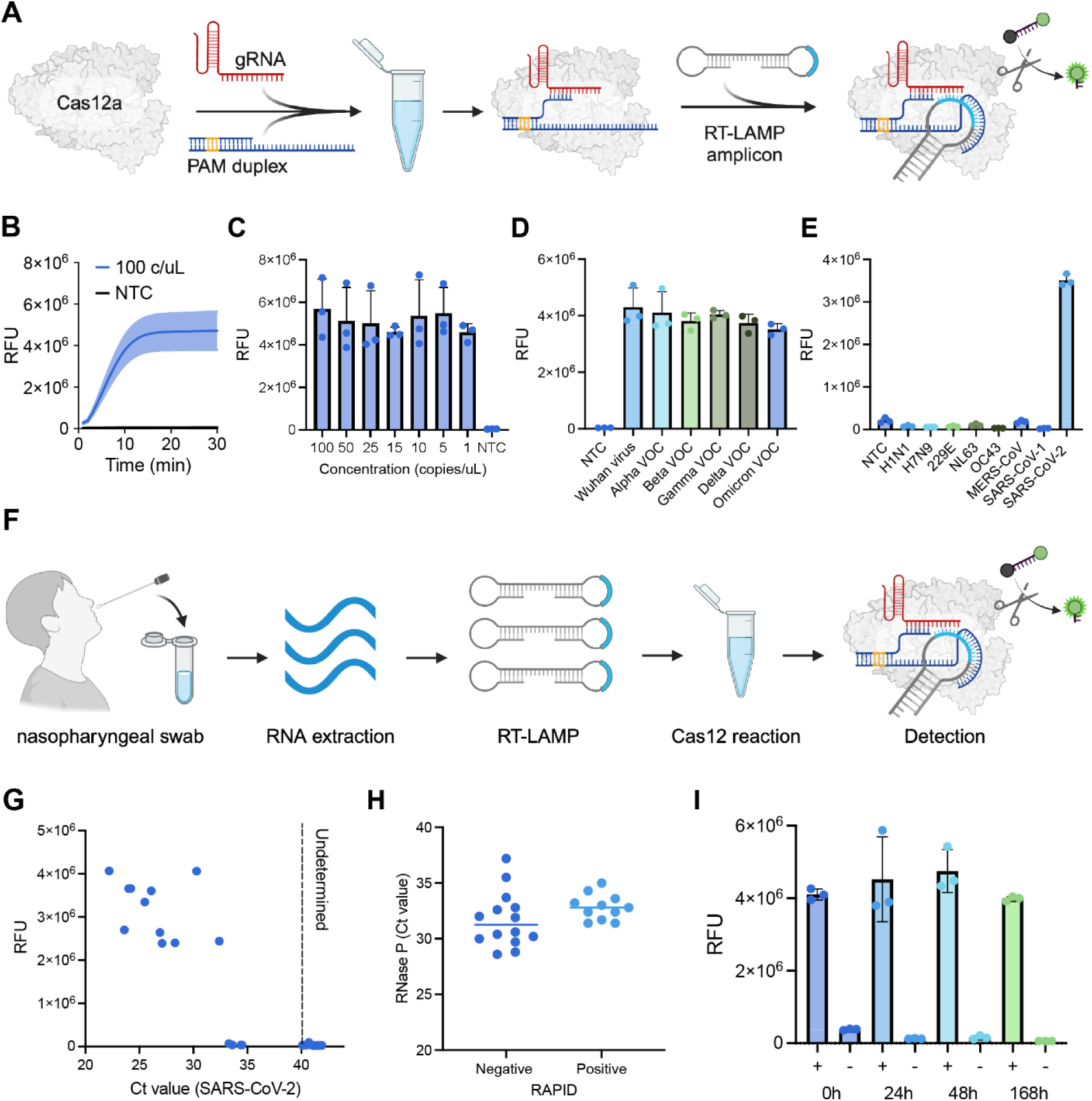
RAPID Coupled with RT-LAMP for SARS-CoV-2 RNA Detection. **(A)** Diagram illustrating the interaction of RAPID with the loop region of the LAMP-generated dumbbell structure. **(B)** Proof of concept demonstrating detection in samples with 100 copies/µL (positive) and control (negative) samples, showcasing that RNA detection with RAPID and LAMP can be completed within 15 minutes. **(C)** Limit of detection data illustrating the ultra-sensitivity of RAPID with LAMP, detecting as low as 0.1 copies/µL within 30 minutes. **(D)** Bar graphs depicting the detection of different SARS-CoV-2 variants using RAPID coupled with LAMP, all within 30 minutes. **(E)** Data demonstrating RAPID’s selectivity against RNA from other viral targets. **(F)** Schematic illustrating the workflow from collection to detection of patient samples and detection using RAPID system coupled with LAMP. **(G)** Screening results for 25 patient samples with 14 negative and 11 positive, respectively. **(H)** Data presenting RNase P values for the 25 patient samples, distinguishing between positive and negative samples. **(I)** Plot showing fluorescence intensity of the freeze-dried RAPID-LAMP assay up to 7 days. Results after 2 hours of incubation at 37 °C (**Methods 13-14**). n=3 technical replicates; bars represent the arithmetic mean ± SD. NTC refers to no template control.

Using the combined RT-LAMP and RAPID assay design, we tested full-length synthetic RNA representative of the SARS-CoV-2 (nucleocapsid gene) variants: Wuhan (Twist _102024), Alpha (Twist _103907), Beta (Twist _104043), Gamma (Twist_104044), Delta (Twist_104533), and Omicron (Twist_105204), finding detection of SARS-CoV-2 positive samples in about 5 minutes following a 30 min RT-LAMP pre-amplification reaction (**Fig. 7B** and **supplementary Fig. S16D, Tables S16-17**). We next screened to optimize the volume of RT-LAMP reaction, finding that a 25 μL volume allowed for maximizing the sample input ratio, while not inhibiting the RAPID reaction (**supplementary Fig. S16B**). Accordingly, this volume was chosen for further experiments.

Having established the conditions for viral RNA detection, we next evaluated the analytical sensitivity for SARS-CoV-2 (Wuhan) of the coupled RT-LAMP/RAPID reaction, finding detection of as low as 1 copy/μL (**Fig. 7C** and **supplementary Fig. S16C**), comparable to standard RT-qPCR methods [59, 60]. Further testing confirmed RAPID’s ability to detect synthetic RNA corresponding to SARS-CoV-2 variants of concern (VOCs), including synthetic RNA encoding nucleocapsid alpha, beta, gamma, delta, and omicron, as well as the original Wuhan strain (**Fig. 7D, supplementary Fig. S16D, Table S17**). Next, a selectivity study was performed to test RAPID’s capacity to discriminate synthetic SARS-CoV-2 target RNA sequences from other viral respiratory pathogens, including influenza virus (H1N1 and H7N9) and other coronaviruses (229E, NL63, OC43, MERS-CoV, and SARS-CoV-1) (**Fig. 7E** and **supplementary Fig. S16E**).

We next set out to evaluate the diagnostic performance of RT-LAMP/RAPID using clinical samples from suspected cases of SARS-CoV-2 infection collected in Toronto, Canada. (**Fig. 7F, Methods 13-14**). A total of 25 nasopharyngeal samples were analysed using the RAPID diagnostic platform in parallel with gold standard RT–qPCR (U.S. CDC protocol [37]), which found full concordance with RT-qPCR for samples with Ct values <33 (95% confidence interval [CI], 83.89% to 100.00%; **Fig. 7G,H**; **Table S19**). Of the 25 patient samples tested for SARS-CoV-2, four samples with Ct values >33 demonstrated false negative results (**Tables S20-21**), reducing accuracy to 84.00% (95% CI, 63.92% to 95.46%) (**Table S20**). These results support a role for RAPID in providing isothermal and low-burden screening of viral RNA targets. To ensure the RNA integrity of the patient samples and as a criterion for sample inclusion in this phase of the project, patient RNaseP measurement was performed using RT-qPCR (**Fig. 7H, Table S18**), with Ct values for RNaseP ranging from 28.8 to 37.2 (**Table S19**).Extending our earlier work with lyophilization of RAPID for miRNA detection, here, individual freeze-dried RAPID and RT-LAMP reactions were stored at room temperature to evaluate shelf stability (**Fig. 7I** and **supplementary Fig. S16F,G**). We tested the system stability up to one week, with the 2-hour endpoint reported (37 °C). The results were comparable to previously reported CRISPR-based freeze-drying [35, 61]. While these results validate the potential of RAPID for field use, they also suggest that further improvements could enhance its long-term stability beyond one week. We anticipate that by using commercial lyophilization processes to produce freeze-dried pellets and provide more effective packaging methods, the combined system has the potential for deployment as a decentralized medical diagnostic. Altogether, with capacity for PAM-independent detection and capacity for multimodal formats of nucleic acid, RAPID represents a significant leap in CRISPR-based diagnostics.

## DISCUSSION

In summary, we have discovered that the presence of a nick on the target strand of dsDNA can modulate Cas12a activity, with the effect—activation or deactivation—dependent on the nick location (**Fig. 1**). It is possible that this phenomenon mirrors the natural immune defense process, wherein the gRNA-guided Cas12a system not only identifies the protospacer sequence but also verifies its integrity. By introducing breaks or nicks, disruptions in the sequence of the target strand can lead to *trans* cleavage-controlled activation when bound to Cas12a ribonucleoproteins. We have also demonstrated, for the first time, that *in situ* ligation of pre-existing nicks can fully restore Cas enzyme activation signals (**Fig. 4**). Building upon this insight, we have developed a PAM–independent system that enables targeting of nucleic acids that do not contain a PAM sequence by modifying both the spacer length and altering its nucleic acid structure (**Fig. 2**).

With the RAPID system, we achieved for the first time a universal approach to target dsDNA compatible with both Cas12a orthologs: AsCas12a and LbCas12a, without traditional PAM constraints. We found that dsDNA could be recognized at blunt ends in a PAM-independent manner (**Fig. 2A(III), 2D**). Moreover, our system also proved capable of detecting PAM-independent dsDNA within a sequence (**Fig. 2A(IV), 2D**), adding an important new tool for directly targeting PAM-independent dsDNA. Our results indicate that AsCas12a exhibits greater versatility than LbCas12a in targeting various nucleic acid types, facilitated by RAPID. Thus, AsCas12a can effectively target both RNA and DNA, while LbCas12a shows a preference for DNA (**Fig. 2B-D**). This contrasts with previous work [29, 30], which demonstrated PAM-free detection via toehold activation and temperature modulation with Cas12a. With this distinction, RAPID is positioned as a “target PAM-independent” system.

While Cas12a traditionally shows weaker RNase activity compared to DNase activity, we enhanced the signal-to-noise ratio for AsCas12a by engineering RNA reporter sequences to incorporate interspaced DNA base modifications, creating chimeric sequence reporters. This modification significantly improved the signal intensity and reduced background noise, which varies between AsCas12a and LbCas12a, with the former generally exhibiting higher background levels (**Fig. 3**). The use of chimeric probes has thus proven, for the first time, to be an enabling method for boosting the analytical sensitivity of CRISPR-based diagnostics. Leveraging RNA-guided nucleic acid targeting, RAPID demonstrates effective *trans* cleavage of chimeric reporters, broadening our understanding of the mechanisms by which the Cas12a system recognizes and acts upon nucleic acids in *trans* cleavage contexts.

Capitalizing on these advancements, we introduced RAPID—a sensitive and selective diagnostic method that can detect ssDNA, dsDNA, miRNA, and long RNA without PAM restrictions. Enhanced by chimeric sequence reporters, RAPID achieves picomolar level sensitivity for direct detection of miRNAs (**Fig. 6**), rivaling the RNA-targeting capabilities of the Cas13 system [55, 56]. Importantly, conventional Cas12a and Cas13 systems do not effectively detect point mutations in ssDNA and RNA, respectively [31, 33]. RAPID, however, exhibits improved and robust selectivity for single- and double-point mutations in ssDNA and can discriminate point mutations in miRNA at several sites, making it suitable for SNP genotyping (**Fig. 5**). Combined with isothermal pre-amplification via RT-LAMP, RAPID reaches PCR-level performance within 40 minutes (10 min post 30 min RT-LAMP reaction) (**Fig. 7B**). Clinical validation of RAPID using SARS-CoV-2 patient samples showed complete concordance with RT-qPCR results for nasopharyngeal swab samples with Ct ≤33 in our cohort (**Fig. 7G**). Acknowledging the limitations of the system, RAPID was not able to detect samples with Ct >33, RT-LAMP/RAPID leading to false-negative calls. However, this performance is consistent with prior reports indicating reduced sensitivity of RT-LAMP and CRISPR-based detection systems at low template concentrations (high Ct values), as well as the assay-dependent nature of Ct thresholds and their relationship to viral load [6, 62, 63].

To set our work with RAPID into context, our efforts are part of a larger effort in the community to relax Cas12a’s canonical PAM constraint by engineering PAM-independent DNA activators and guide/activator splitting architectures [23, 29, 64, 65]. Methods including toehold-mediated strand displacement and pre-unwound (“bubble”) seed regions have been used to convert diverse sequences into duplex activators that engage the gRNA seed without requiring a flanking PAM, enabling PAM-independent SNP detection and modular DNA-circuit readouts [29, 64, 65]. In parallel, split-activator (such as PHD [27]) and split-gRNA strategies (such as the SCas12a system [58]) decouple target binding from nuclease activation to enable amplification-free RNA detection and multiplexing, with reported limits ranging from picomolar to ∼100 fM fluorescence/∼10 fM lateral-flow (split crRNA), and attomolar performance when coupled to RPA [23, 58]. These approaches can remain sequence- and design-dependent and may require careful mitigation of background activation and/or pre-amplification at very low template inputs; RAPID differs by using nick-position–dependent tuning of a protospacer and a trans-supplied PAM duplex (plus chimeric reporters) to reach PAM-independent activation through a distinct mechanistic route – **Fig. 2**.

In closing, RAPID establishes a fundamentally new framework for CRISPR/Cas12a diagnostics by integrating nick-position–dependent activation, a trans-supplied PAM duplex for true PAM-independent targeting, and engineered chimeric reporters that enhance signal fidelity while minimizing background. Together, these innovations enable versatile detection across ssDNA, dsDNA, and RNA substrates, including miRNAs, while supporting amplification-free sensitivity, robust SNP discrimination, and compatibility with isothermal amplification workflows for clinical deployment. Beyond expanding the functional landscape of Cas12a, RAPID provides a tunable and modular architecture that bridges mechanistic insight with practical diagnostic design. We anticipate that these principles will catalyze further advances in CRISPR engineering, including improved enzyme variants, orthogonal reporter systems, and integrated point-of-care platforms. As the community builds upon these concepts, we envision RAPID contributing to the development of scalable, decentralized, and precision-driven diagnostic technologies that extend the reach of molecular testing in both research and global healthcare settings.

## Acknowledgement

I.A.I. was supported by the Precision Medicine Initiative (PRiME) and Canadian Institute of Health Research (CIHR) at the University of Toronto with internal fellowship numbers PRMUHT2024-001 and 202410MFE-531769-419793, respectively. J.R.J.V. was supported by the Ontario Graduate Scholarship (OGS). This work was also supported by funds to K.P from the CIHR Foundation Grant Program (201610FDN-375469); the Canada Research Chairs Program (Files 950-231075 and 950-233107); the NSERC Discovery Grants Program (RGPIN-2016-06352). Furthermore, this work was also developed with funding from the Defense Advanced Research Projects Agency (DARPA), Contract No. N66001-23-2-4042 to AAG and KP. R.C and B.P.H. were supported by CIHR project grant PJT-189974.The views, opinions and/or findings expressed are those of the authors and should not be interpreted as representing the official views or policies of the Department of Defense or the U.S. Government. Special thanks to Profs. Zhigang Li and Shuhuai YAO at the Hong Kong University of Science and Technology for their supervision and discussions at the initial stage of the project. I.A.I. thanks Abasi-ifreke Karena Idorenyin, Zachary Idorenyin Iwe, and Ariel Idorenyin Iwe for their understanding and support during this work.

## Approval for Public Release

This work was cleared by DARPA and approved for public release, distribution unlimited. Further questions should be channeled to DARPA via their Public Release Center located at 675 N. Randolph Street, Room 03-028, Arlington, VA 22203-1714.

## Author contributions

I.A.I. and F.L. contributed equally and conceived the ideas. K.P., I.A.I., F.L., A.C., J.D., J.N., P.K., R.C., B.P.H., and G.L. designed, performed, and analysed the experiments. X.L. contributed in shaping initial ideas. I.A.I. guided all experiments. I.A.I, F.L., S.S., J.D., A.C., S.F.J.D.S., and G.L. wrote the manuscript. S.F.J.D.S. handled all clinical trials. T.M. provided all the patient samples. A.A.G. provided useful insights to miRNA sensing. Y.Z. handled Cas structural analysis. I.A.I., F.L., A.C., S.J.D.S., J.D., S.S., Q.M., J.R.J.V., R.Z., R.C., P.K., P.B., M.S., K.B., M.C., B.P.H., S.P., X.L., J.P.T., Y.Z., K.P., and A.A.G. discussed the results, revised or commented on the manuscript. S.Y. and Z.L. supervised the phase I of the work. K.P. edited the manuscript and supervised the rest of the work.

## Ethics declarations

### Data availability

Materials are available from Keith Pardee upon request and subject to completion of a materials transfer agreement with the Leslie Dan Faculty of Pharmacy, University of Toronto. Any additional information required to reanalyze the data reported in this article is available from the lead contact upon request.

The data underlying this article are available in the article and in its online supplementary material.

Sequences for synthetic nucleic acid oligonucleotides used in this work are detailed in the relevant sections of the supplementary material.

### Supplementary information

**Supplementary Figs. S1-16** and **Supplementary Tables S1-21**

## References

1. Mohammad N, Talton L, Hetzler Z et al. Unidirectional trans-cleaving behavior of CRISPR-Cas12a unlocks for an ultrasensitive assay using hybrid DNA reporters containing a 3′ toehold. Nucleic Acids Res 2023;51:9894–9904. 10.1093/nar/gkad715

2. Chen JS, Ma E, Harrington LB et al. CRISPR-Cas12a target binding unleashes indiscriminate single-stranded DNase activity. Science 2018;360:436–439. 10.1126/science.aar6245

3. Makarova KS, Wolf YI, Iranzo J et al. Evolutionary classification of CRISPR–Cas systems: a burst of class 2 and derived variants. Nat Rev Microbiol 2020;18:67–83. 10.1038/s41579-019-0299-x

4. Liu FX, Cui JQ, Wu Z et al. Recent progress in nucleic acid detection with CRISPR. Lab on a Chip 2023;23:1467–1492. 10.1039/d2lc00928e

5. Kaminski MM, Abudayyeh OO, Gootenberg JS et al. CRISPR-based diagnostics. Nat Biomed Engin 2021;5:643–656. 10.1038/s41551-021-00760-7

6. Broughton JP, Deng X, Yu G et al. CRISPR–Cas12-based detection of SARS-CoV-2. Nat Biotechnol 2020;38:870–874. 10.1038/s41587-020-0513-4

7. De Puig H, Lee RA, Najjar D et al. Minimally instrumented SHERLOCK (miSHERLOCK) for CRISPR-based point-of-care diagnosis of SARS-CoV-2 and emerging variants. Science Advances 2021;7:eabh2944. 10.1126/sciadv.abh2944

8. Huyke DA, Ramachandran A, Bashkirov VI et al. Enzyme kinetics and detector sensitivity determine limits of detection of amplification-free CRISPR-Cas12 and CRISPR-Cas13 diagnostics. Anal Chem 2022;94:9826–9834. 10.1021/acs.analchem.2c01670

9. Liu FX, Cui JQ, Park H, et al. Isothermal background-free nucleic acid quantification by a one-pot Cas13a assay using droplet microfluidics. Anal Chem 2022;94:5883–5892. 10.1021/acs.analchem.2c00067

10. Barnes KG, Lachenauer AE, Nitido A et al. Deployable CRISPR-Cas13a diagnostic tools to detect and report Ebola and Lassa virus cases in real-time. Nat Commun 2020;11:4131. 10.1038/s41467-020-17994-9

11. Myhrvold C, Freije CA, Gootenberg JS et al. Field-deployable viral diagnostics using CRISPR-Cas13. Science 2018;360:444–448. 10.1126/science.aas8836

12. Pardee K, Green AA, Takahashi MK et al. Rapid, low-cost detection of Zika virus using programmable biomolecular components. Cell 2016;165:1255–1266. 10.1016/j.cell.2016.04.059

13. Karlikow M, Amalfitano E, Yang X et al. CRISPR-induced DNA reorganization for multiplexed nucleic acid detection. Nat Commun 2023;14:1505. 10.1038/s41467-023-36874-6

14. Chertow DS. Next-generation diagnostics with CRISPR. Science 2018;360:381–382. 10.1126/science.aat4982

15. Bravo JPK, Hallmark T, Naegle B et al. RNA targeting unleashes indiscriminate nuclease activity of CRISPR–Cas12a2. Nature 2023;613:582–587. 10.1038/s41586-022-05560-w

16. Li Z, Zhang H, Xiao R et al. Cryo-EM structure of the RNA-guided ribonuclease Cas12g. Nat Chem Biol 2021;17:387–393. 10.1038/s41589-020-00721-2

17. Jiao C, Sharma S, Dugar et al. Noncanonical crRNAs derived from host transcripts enable multiplexable RNA detection by Cas9. Science 2021;372:941–948. 10.1126/science.abe7106

18. Nguyen LT, Smith BM, Jain PK. Enhancement of trans-cleavage activity of Cas12a with engineered crRNA enables amplified nucleic acid detection. Nat Commun 2020;11:4906. 10.1038/s41467-020-18615-1

19. Fozouni P, Son S, Derby MDdL et al. Amplification-free detection of SARS-CoV-2 with CRISPR-Cas13a and mobile phone microscopy. Cell 2021;184:323–333. 10.1016/j.cell.2020.12.001

20. Ding X, Yin K, Li Z et al. Ultrasensitive and visual detection of SARS-CoV-2 using all-in-one dual CRISPR-Cas12a assay. Nat Commun 2020;11:4711. 10.1038/s41467-020-18575-6

21. Ooi KH, Liu MM, Tay JWD et al. An engineered CRISPR-Cas12a variant and DNA-RNA hybrid guides enable robust and rapid COVID-19 testing. Nat Commun 2021;12:1739. 10.1038/s41467-021-21996-6

22. Yang J, Song Y, Deng X et al. Engineered LwaCas13a with enhanced collateral activity for nucleic acid detection. Nat Chem Biol 2023;19:45–54. 10.1038/s41589-022-01135-y

23. Rananaware SR, Vesco EK, Shoemaker GM et al. Programmable RNA detection with CRISPR-Cas12a. Nat Commun 2023;14:5409. 10.1038/s41467-023-41006-1

24. Li S, Cheng Q, Wang J et al. CRISPR-Cas12a-assisted nucleic acid detection. Cell Discovery 2018;4:20. 10.1038/s41421-018-0028-z

25. Idorenyin IA. Hairpin DNA-Mediated Isothermal Amplification Techniques for Nucleic Acid Testing. Hairpin DNA-Mediated Isothermal Amplification Techniques for Nucleic Acid Testing. 2021. Available on ProQuest http://bit.ly/3KApG6r (2 August 2025, date last accessed).

26. Xia X, Chen Q, Zuo T et al. DNA robots for CRISPR/Cas12a activity management and universal platforms for biosensing. Anal Chem 2024;96:2620–2627. 10.1021/acs.analchem.3c05210

27. Qiao J, Zhang J, Jiang Q et al. Boosting CRISPR/Cas12a intrinsic RNA detection capability through pseudo hybrid DNA–RNA substrate design. Nucleic Acids Res 2025;53:gkaf510. 10.1093/nar/gkaf510

28. O’Connell MR, Oakes BL, Sternberg SH et al. Programmable RNA recognition and cleavage by CRISPR/Cas9. Nature 2014;516:263–266. 10.1038/nature13769

29. Wu Y, Luo W, Weng Z et al. A PAM-free CRISPR/Cas12a ultra-specific activation mode based on toehold-mediated strand displacement and branch migration. Nucleic Acids Res 2022;50:11727–11737. 10.1093/nar/gkac886

30. Rananaware SR, Meister KS, Shoemaker GM et al. PAM-free diagnostics with diverse type V CRISPR-Cas systems. 2017. medRxiv, 10.1101/2024.05.02.24306194, 03 May 2024, preprint: not peer reviewed.

31. Mahas A, Marsic T, Masson ML et al. Characterization of a thermostable Cas13 enzyme for one-pot detection of SARS-CoV-2. Proc Natl Acad Sc 2022;119:e2118260119. 10.1073/pnas.2118260119

32. Thorne LG, Bouhaddou M, Reuschl A et al. Evolution of enhanced innate immune evasion by SARS-CoV-2. Nature 2022;602:487–495. 10.1038/s41586-021-04352-y

33. Zhang J, Li Z, Guo C et al. Intrinsic RNA Targeting Triggers Indiscriminate DNase Activity of CRISPRLJCas12a. Angew Chem Intl Ed 2024;63:e202403123. 10.1002/anie.202403123

34. Cheng Z, Luo X, Yu S et al. Tunable control of Cas12 activity promotes universal and fast one-pot nucleic acid detection. Nat Commun 2025;16:1166. 10.1038/s41467-025-56516-3

35. Rybnicky GA, Dixon RA, Kuhn RM et al. Development of a freeze-dried CRISPR-Cas12 sensor for detecting wolbachia in the secondary science classroom. ACS Synthetic biol 2022;11:835–842. 10.1021/acssynbio.1c00503

36. Yan Z, Eshed A, Tang AA et al. Rapid, multiplexed, and enzyme-free nucleic acid detection using programmable aptamer-based RNA switches. Chem 10, 2220–2244 (2024). 10.1016/j.chempr.2024.03.015

37. Centers for Disease Control. Research use only 2019-novel coronavirus (2019-nCoV) real-time RT-PCR primers and probes. 2020. https://stacks.cdc.gov/view/cdc/88834. (10 December 2025, date last accessed).

38. Caldecott KW. Single-strand break repair and genetic disease. Nat Rev Gen 2008;9:619–631. 10.1038/nrg2380

39. Higo T, Naito AT, Sumida T et al. DNA single-strand break-induced DNA damage response causes heart failure. Nat Commun 2017;8:15104. 10.1038/ncomms15104

40. Yamano T, Zetsche B, Ishitani R et al. Structural basis for the canonical and non-canonical PAM recognition by CRISPR-Cpf1. Mol. Cell 2017;67:633–645. 10.1016/j.molcel.2017.06.035

41. Yamano T, Nishimasu H, Zetsche B et al. Crystal structure of Cpf1 in complex with guide RNA and target DNA. Cell 2016;165:949–962.

42. Naqvi MM, Lee L, Montaguth OET et al. CRISPR–Cas12a-mediated DNA clamping triggers target-strand cleavage. Nat Chem Biol 2022;18:1014–1022. 10.1038/s41589-022-01082-8

43. Ramachandran A, Santiago JG. CRISPR enzyme kinetics for molecular diagnostics. Anal Chem 2021;93:7456–7464. 10.1021/acs.analchem.1c00525

44. Lamothe G, Veillette F, Iwe IA et al. Split Cas12a protospacer engineering enables ultra-specific, PAM-free detection. 2025. bioRxiv, 10.1101/2025.07.30.667643, 30 July 2025, preprint: not peer reviewed.

45. Roberts RW, Crothers DM. Stability and properties of double and triple helices: dramatic effects of RNA or DNA backbone composition. Science 1992;258:1463–1466. 10.1126/science.1279808

46. Hong F, Šulc P. An emergent understanding of strand displacement in RNA biology. J. Struct. Biol. 2019;207:241–249. 10.1016/j.jsb.2019.06.005

47. Fuchs RT, Curcuru J, Mabuchi M et al. Cas12a trans-cleavage can be modulated in vitro and is active on ssDNA, dsDNA, and RNA. 2019. bioRxiv, 10.1101/600890, 8 April 2019, preprint: not peer reviewed.

48. Lietard J, Ameur D, Somoza MM. Sequence-dependent quenching of fluorescein fluorescence on single-stranded and double-stranded DNA. RSC Advances 2022;12:5629–5637. 10.1039/D2RA00534D

49. Noble JE, Wang L, Cole KD et al. The effect of overhanging nucleotides on fluorescence properties of hybridising oligonucleotides labelled with Alexa-488 and FAM fluorophores. Biophys. Chem. 2005;113:255–263. 10.1016/j.bpc.2004.09.012

50. Torimura M, Kurata S, Yamada K et al. Fluorescence-quenching phenomenon by photoinduced electron transfer between a fluorescent dye and a nucleotide base. Analyt Sc 2001;17:155–160. 10.2116/analsci.17.155nh

51. Liang Y, Lin H, Zou L et al. CRISPR-Cas12a-based detection for the major SARS-CoV-2 variants of concern. Microbiol Spec 2021;9:1017. 10.1128/Spectrum.01017-21

52. Lv H, Wang J, Zhang et al. Definition of CRISPR Cas12a Trans-cleavage units to facilitate CRISPR diagnostics. Front Microbiol 2021;12:766464. 10.3389/fmicb.2021.766464

53. Dong H, Lei J, Ding L et al. MicroRNA: function, detection, and bioanalysis. Chem Rev 2013;113:6207–6233. 10.1021/cr300362f

54. Pritchard CC, Cheng HH, Tewari M. MicroRNA profiling: approaches and considerations. Nat Rev Gen 2012;13:358–369. 10.1038/nrg3198

55. Shan Y, Zhou X, Huang R. High-fidelity and rapid quantification of miRNA combining crRNA programmability and CRISPR/Cas13a trans-cleavage activity. Anal Chem 2019;91:5278–5285. 10.1021/acs.analchem.9b00073

56. Bruch R, Baaske J, Chatelle et al. CRISPR/Cas13aLJpowered electrochemical microfluidic biosensor for nucleic acid amplificationLJfree miRNA diagnostics. Adv Mater 2019;31:1905311. 10.1002/adma.201905311

57. Somers VA, Moerkerk PT, Murtagh JJ et al. A rapid, reliable method for detection of known point mutations: point-EXACCT. Nucleic Acids Res 1994;22:4840. 10.1093/nar/22.22.4840

58. Chen Y, Wang X, Zhang J et al. Split crRNA with CRISPR-Cas12a enabling highly sensitive and multiplexed detection of RNA and DNA. Nat Commun 2024;15:8342. 10.1038/s41467-024-52691-x

59. Vogels CBF, Brito AF, Wyllie AL et al. Analytical sensitivity and efficiency comparisons of SARS-CoV-2 RT–qPCR primer–probe sets. Nat Microbiol 2020;5:1299–1305. 10.1038/s41564-020-0761-6

60. Karlikow M, da Silva SJR, Guo Y et al. Field validation of the performance of paper-based tests for the detection of the Zika and chikungunya viruses in serum samples. Nat Biomed Engin 2022;6:246–256. 10.1038/s41551-022-00850-0

61. Arizti-Sanz J, Bradley A, Zhang YB et al. Simplified Cas13-based assays for the fast identification of SARS-CoV-2 and its variants. Nat Biomed Engin 2022;6:932–943. 10.1038/s41551-022-00889-z

62. Thi VLD, Herbst K, Boerner K et al. A colorimetric RT-LAMP assay and LAMP-sequencing for detecting SARS-CoV-2 RNA in clinical samples. Sc Trans Med 2020;12:eabc7075. 10.1126/scitranslmed.abc7075

63. Ali Z, Aman R, Mahas A et al. iSCAN: An RT-LAMP-coupled CRISPR-Cas12 module for rapid, sensitive detection of SARS-CoV-2. Virus Res 2020;288:198129. 10.1016/j.virusres.2020.198129

64. Shi K, Yi K, Han Y et al. PAM-free cascaded strand displacement coupled with CRISPR-Cas12a for amplified electrochemical detection of SARS-CoV-2 RNA. Anal Biochem. 2023;664:115046. 10.1016/j.ab.2023.115046

65. Chen S, Wang R, Peng S et al. PAM-less conditional DNA substrates leverage trans-cleavage of CRISPR-Cas12a for versatile live-cell biosensing. Chem Sc 2022;13:2011–2020. 10.1039/D1SC05558E

