## Supplementary Information for "RAPID: Evaluation of Cas12a Protospacer Nicking and Chimeric Reporters for PAM-independent RNA and DNA diagnostics"

### Table of contents

Supplementary Figures 1-7

**Table S1:** gRNA sequence and DNA oligos used for proof of concept.

**Table S2**: Nucleic acid oligos for nick testing on the non-target strand.

**Table S3:** gRNA sequence and DNA oligos used for proof of concept 2.

**Table S4**: gRNA sequence and DNA oligos used for proof of concept 3.

**Table S5**: Structure of AsCas12 in complex with gRNA and target DNA

**Table S6**: All nucleic acid used for testing RAPID against ssDNA, RNA, and dsDNA.

**Table S7:** Oligos and modified probes for reporter screening.

**Table S8**: HPV 18-related sequences to validate chimeric probes.

**Table S9**: Mass spectrometry–resolved cleavage profiles of RAPID reporters.

**Table S10**: Sequences for RAPID proof-of-concept diagnostic for monkeypox dsDNA virus

**Table S11**: All sequences used for the nicked DNA repair and miRNA-DNA Ligation

**Table S12**: Sequences for miRNA orthogonality testing

**Table S13**: Nucleic acid sequence for miRNA detection.

**Table S14**: Nucleic acid sequences for DNA mismatched Screening.

**Table S15**: Oligos for screening point mutations in miRNA-21.

**Table S6:** Nucleic acid sequences used for RAPID-LAMP assay for SARS_CoV-2 detection.

**Table S7**: Synthetic RNA controls for molecular reactions.

**Table S8**: RT-qPCR oligos used in this study for SARS-CoV-2.

**Table S99**: Comparison of SARS-Cov2 patient sample data with RAPID-LAMP and RT-qPCR.

**Table S20**: Patient samples (Ct value <40).

**Table S21**: Patient samples (Ct value ≤33).

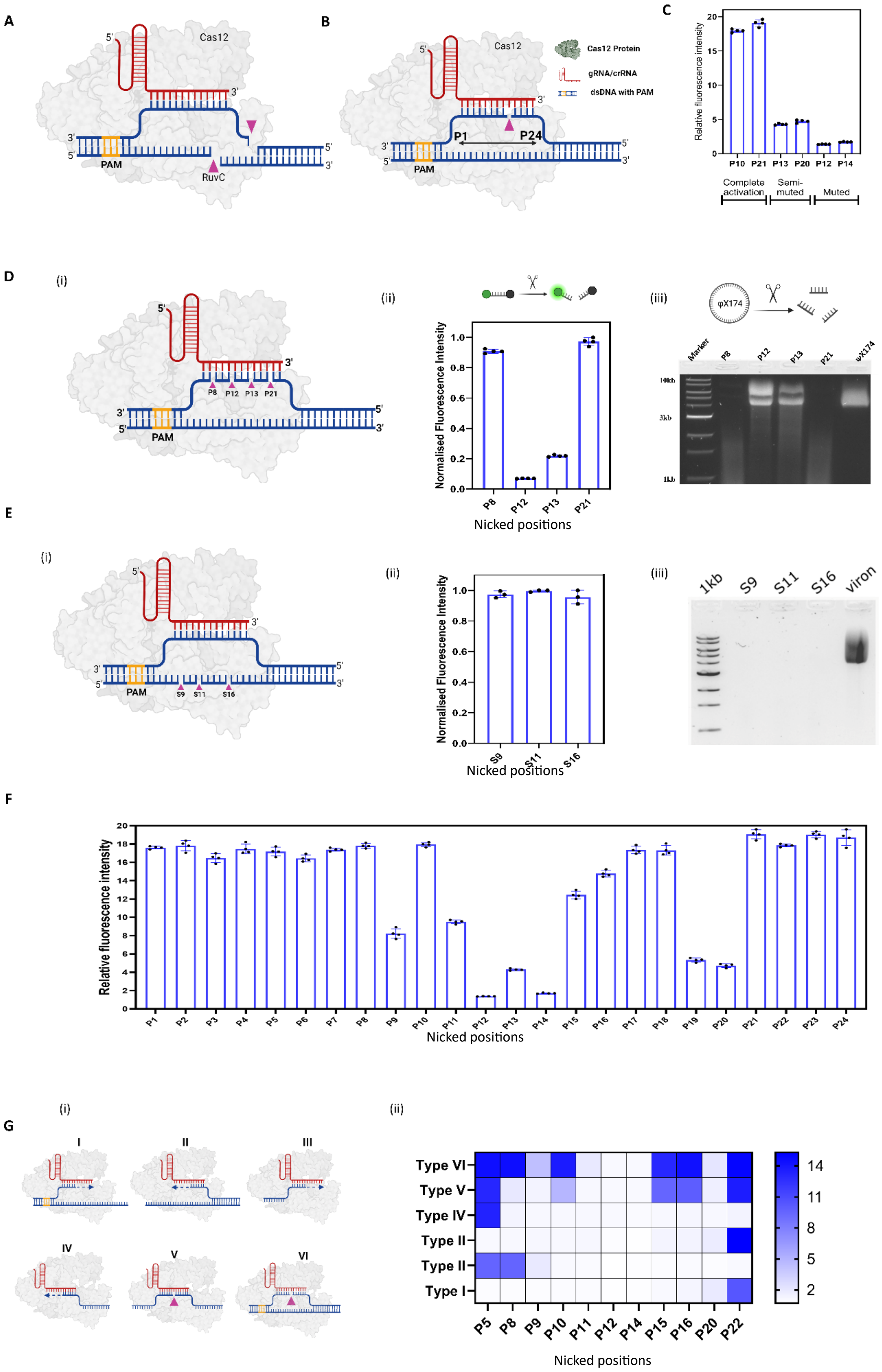

**Supplementary Fig. S1**: **Overview of the developed platform. (A)** Cas12a-induced double-stranded breaks controlled by the provision of the PAM motif, which is fundamental for gene editing. **(B)** Representation of dsDNA featuring a single-stranded break (nick) in association with the Cas12a ribonucleoprotein complex. **(C)** Graph showing Cas12a activation at various nick positions on dsDNA. Positions P10 and P21 show full activation, positions P13 and P20 show partial activation, and positions P12 and P14 show no activation. Corresponding nucleic acid sequences are listed in Table S1. **(D)** Selection and analysis of nick positions on the target strand of PAM-containing dsDNA: (i) Schematic illustration of selected nick positions. (ii) Normalized *trans*-cleavage fluorescence intensity at these nick positions. (iii) Agarose gel electrophoresis displaying the *trans*-cleaved bands of φX174 virion circular ssDNA corresponding to the nicked positions, compared to the φX174 virion DNA template. **(E)** Analysis of nicks on the non-target strand of PAM-containing dsDNA: (i) Schematic showing randomly selected nick positions on the non-target strand at positions S9, S11, and S16. (ii) Normalized fluorescence intensity at these positions, indicating no significant Cas12a activation differences. (iii) Agarose gel demonstrating the electrophoretic mobility of *trans*-cleaved φX174 virion DNA template, which was completely cleaved at these positions, suggesting that Cas12a's tunability is not evident when the non-target strand is nicked. **(F)** Graph depicting *trans*-cleavage fluorescence intensity across all considered nick positions on the target strand of dsDNA (P1 to P24). The Cas12a activation pattern is visible across these positions, with P12 and P14 showing no activation, while positions P9, P11, P13, P19, and P20 exhibit partial activation. All corresponding nucleic acid sequences are available in Table S1. **(G)** Illustration of Cas12a tunability testing using a different gRNA to demonstrate the universality of the developed platform: (i) Diagram of six RAPID system configurations similar to those described in the main text. (ii) Heatmap showing the activation pattern of Cas12a at selected nick positions across the six system types, in complete agreement with the initial findings. The fluorescence intensities at selected positions correspond to the heatmap in Fig. 1 of the main text. All nucleic acid sequences are detailed in Tables S1-3. Experiments were conducted using the LbCas12a enzyme.

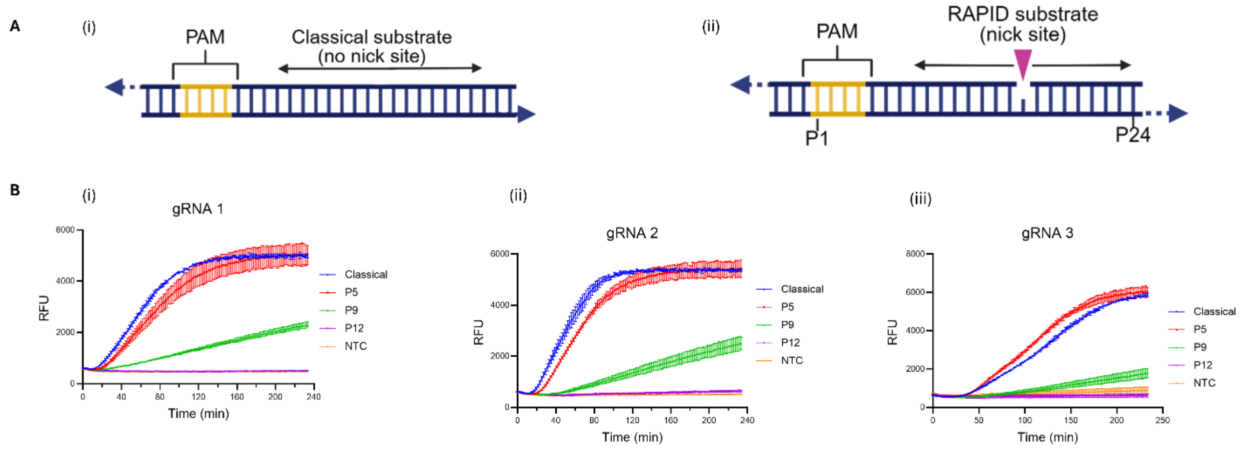

**Supplementary Fig. S2.** (A) Classical substrate without nicks (i) and RAPID substrate containing target strand nicks (ii). (B) Kinetic plots comparison of the classical reporter system lacking a nick with nicked substrates corresponding to complete activation (P5), partial activation (P9), and completely muted activation (silent, P12). These conditions were evaluated across three distinct substrates with entirely different sequences (**i–iii**). See Tables S1, S3, and S4 for the corresponding sequences. Experiments were conducted using the LbCas12a enzyme.

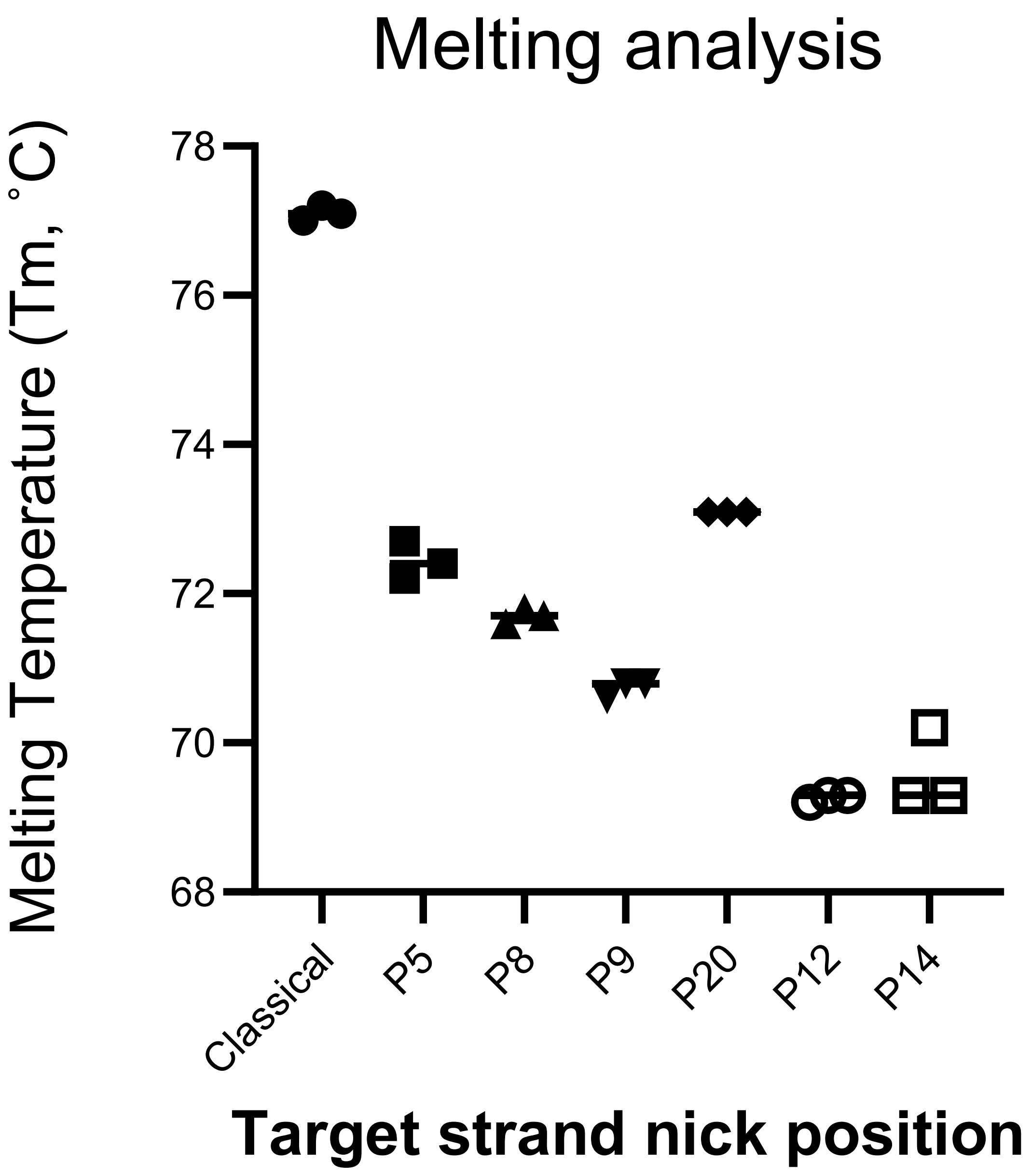

**Supplementary Fig. S3.** Melting temperature (Tm, ˚C) of Cas12 dsDNA target, with location specific nicks on the target strand. Tm was calculated from three technical replicates (n=3), mean +/- S.D. is shown.

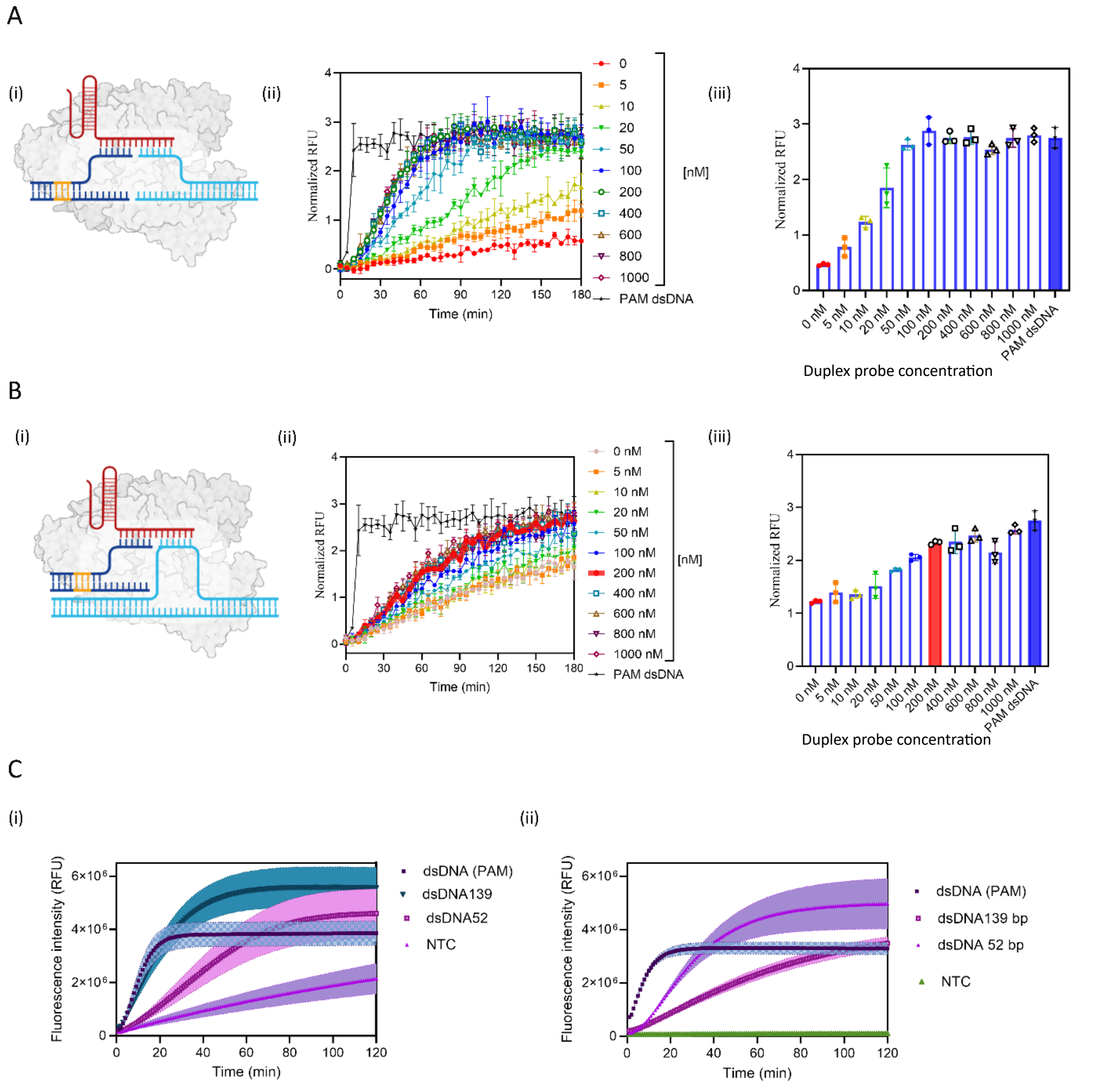

**Supplementary Fig. S4: Optimization of RAPID for PAM-free detection of dsDNA. (A) Universal *trans* PAM for blunt-ended dsDNA: (i)** Schematic illustrating the universal PAM (in deep blue) paired with blunt-ended dsDNA (in light blue). **(ii)** Kinetic curve showing the concentration of universal PAM ranging from 0 to 1000 nM with an optimal concentration around 200 nM, benchmarked against PAM-containing dsDNA (black curve). The results demonstrate that the fluorescence signal for blunt-ended PAM-free dsDNA plateaus around 60 minutes with an intensity comparable to that of PAM-containing dsDNA, indicating effective direct detection with the aid of the universal PAM. **(iii)** Bar plots representing fold changes in fluorescence intensity based on universal PAM concentrations (from 0 to 1000 nM) at 120 minutes. The plain blue bar represents the benchmark using PAM-containing dsDNA. Experiments conducted with LbCas12a; blunt-ended dsDNA at a final concentration of 10 nM. **(B) Interaction of universal PAM with PAM-free dsDNA at arbitrary locations other than the blunt ends: (i)** Assembly diagram of the ribonucleoprotein with universal PAM and PAM-free dsDNA. **(ii)** Time-dependent plot showing concentration variation of the universal PAM from 0 to 1000 nM, with an optimum concentration at 200 nM. Notably, the background signal at 0 nM is higher compared to that with blunt-ended dsDNA, possibly due to suboptimal PAM mimic formations by DNA motifs.[2] Signal enhancement was observed with the addition of the universal PAM. **(iii)** Data representation at 120 minutes showing universal PAM concentrations from 0 to 1000 nM compared to the signal from PAM-containing dsDNA (plain blue bar). Experiments conducted with LbCas12a; dsDNA at a final concentration of 10 nM. **(C) Detection of PAM-free dsDNA using AsCas12a and LbCas12a: (i)** Kinetic plot comparing fluorescence intensities from blunt-ended PAM-free dsDNA (52-bp) and PAM-free dsDNA (139-bp) to PAM-containing dsDNA, indicating similar fluorescence intensities. NTC (negative control) shows the intrinsic high background signal of AsCas12a as previously mentioned in the main text. All dsDNAs were at a final concentration of 50 nM. **(ii)** Performance comparison plots using LbCas12a for PAM-free blunt-ended dsDNA (52-bp) and PAM-free dsDNA (139-bp) relative to PAM-containing dsDNA. While all templates showed comparable performance, the response with PAM-containing dsDNA was faster than with PAM-free templates. All dsDNAs were tested at a concentration of 50 nM.

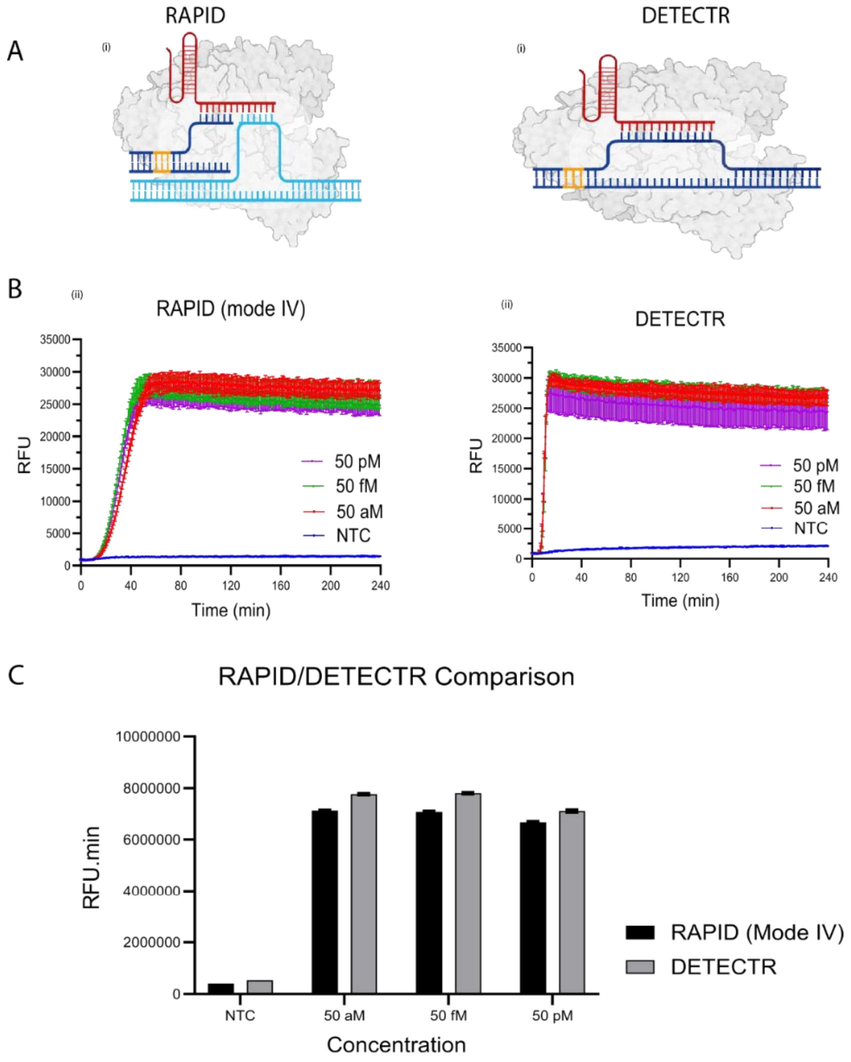

**Supplementary Fig. S5. Comparison of RAPID Mode IV and classical DETECTR for RPA-amplified MPXV dsDNA detection.** (A) RAPID (i) and DETECTR (ii) configurations. (A-i) RAPID Mode IV detection of an RPA-amplified MPXV dsDNA target across a dynamic concentration range (pM, fM, and aM), benchmarked against a non-template control (NTC). Fluorescence kinetics demonstrate sensitive detection over multiple orders of magnitude. (A-ii) Detection of the same RPA-amplified MPXV target using the classical DETECTR system under comparable conditions. While both platforms achieve similar sensitivity, DETECTR exhibits faster initial reaction kinetics, consistent with canonical Cas12a activation on intact dsDNA substrates. (C) Quantitative comparison of trans-cleavage activity between RAPID and DETECTR platforms. The area under the fluorescence curve (AUC; RFU·min) was used as an integrated measure of trans-cleavage output. Comparable overall activity is observed between the two systems despite slight differences in kinetic profiles. All reactions were performed using Reporter 7, with n = 3 technical replicates; bars represent the arithmetic mean ± standard deviation (SD). Experimental conditions are in the Method, while sequences and target designs are provided in Table S9.

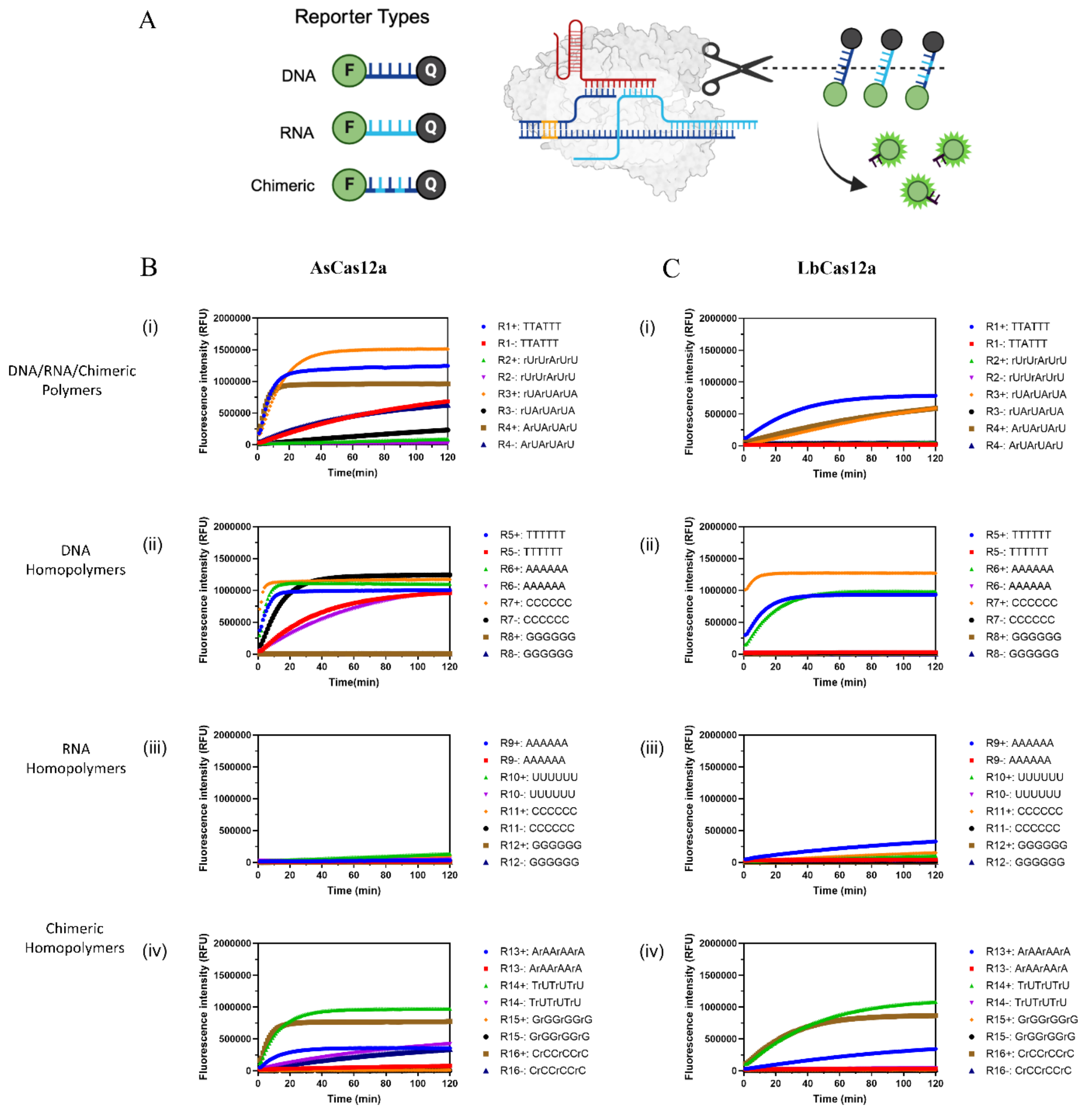

**Supplementary Fig. S6: Cas12a collateral cleavage of natural and non-natural nucleic acid reporter sequences. (A)** Schematic illustration of RAPID system *trans*-cleaving DNA/RNA/chimeric sequences. This diagram provides a visual representation of the experimental setup used to assess the specificity and efficiency of Cas12a cleavage across various nucleic acid types. **(B) & (C)** contain Kinetic plots showing the activity of AsCas12a and LbCas12a on 16 different nucleic acid reporter sequences. These sequences are categorized into four groups: **(i)** DNA/RNA/chimeric polymers (Reporters R1 to R4), where R1 serves as the traditional reporter for Cas12a but shows a high background signal particularly with AsCas12a. R3, however, significantly reduces this background while maintaining high cleavage efficiency. **(ii)** DNA homopolymers (Reporters R5 to R8), with certain polymers like R5, R6, and R7 exhibiting extremely high background signals with AsCas12a but very low signals with LbCas12a. **(iii)** RNA homopolymers (Reporters R9 to R12), which generally do not undergo cleavage by either AsCas12a or LbCas12a. **(iv)** Chimeric homopolymers (Reporters R13 to R16), where R13, R14, and R16 sequences showed high *trans* cleavage signal for both AsCas12a and LbCas12a, respectively. Additionally, R13 exhibited lowest background noise for AsCas12a. Poly G sequences (R8, R12, and R15) were resistant to cleavage for DNA, RNA, and chimeric homopolymers. The '+' and '-' signs indicate experiments conducted with and without a DNA trigger, respectively.

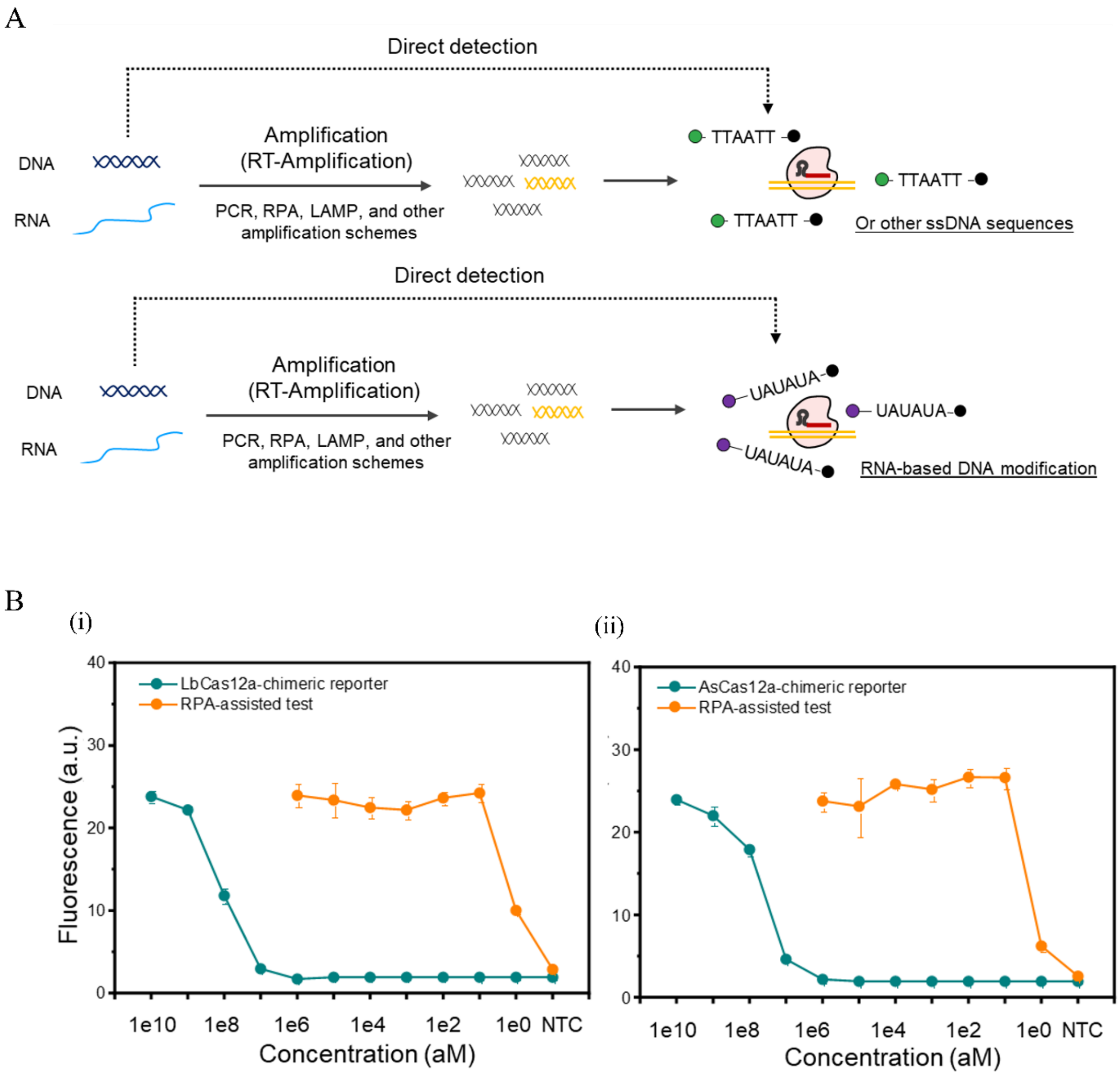

**Supplementary Fig. S7**: **Performance testing with chimeric reporter. (A)** Scheme illustrating the direct and amplification-assisted detection of PAM-containing dsDNA or RNA using standard and chimeric reporter sequences. **(B) Amplification-free and amplification-assisted (RPA) for PAM-containing nucleic acid detection for non-canonical (R3: $(rUA)_3$) reporter sequences to detect synthetic HPV-18. (i -ii)** Performance comparison of LbCas12a and AsCas12a nucleic acid detection between amplification-free and amplification-assisted strategies. Orange curve depicts amplification-assisted detection while the green curve represents direct detection. All experiments were conducted in triplicate. For the amplification-free test, the incubation time was 60 minutes. For the RPA-assisted reaction, the RPA reaction time was 20 minutes, followed by a 40-minute CRISPR reaction.

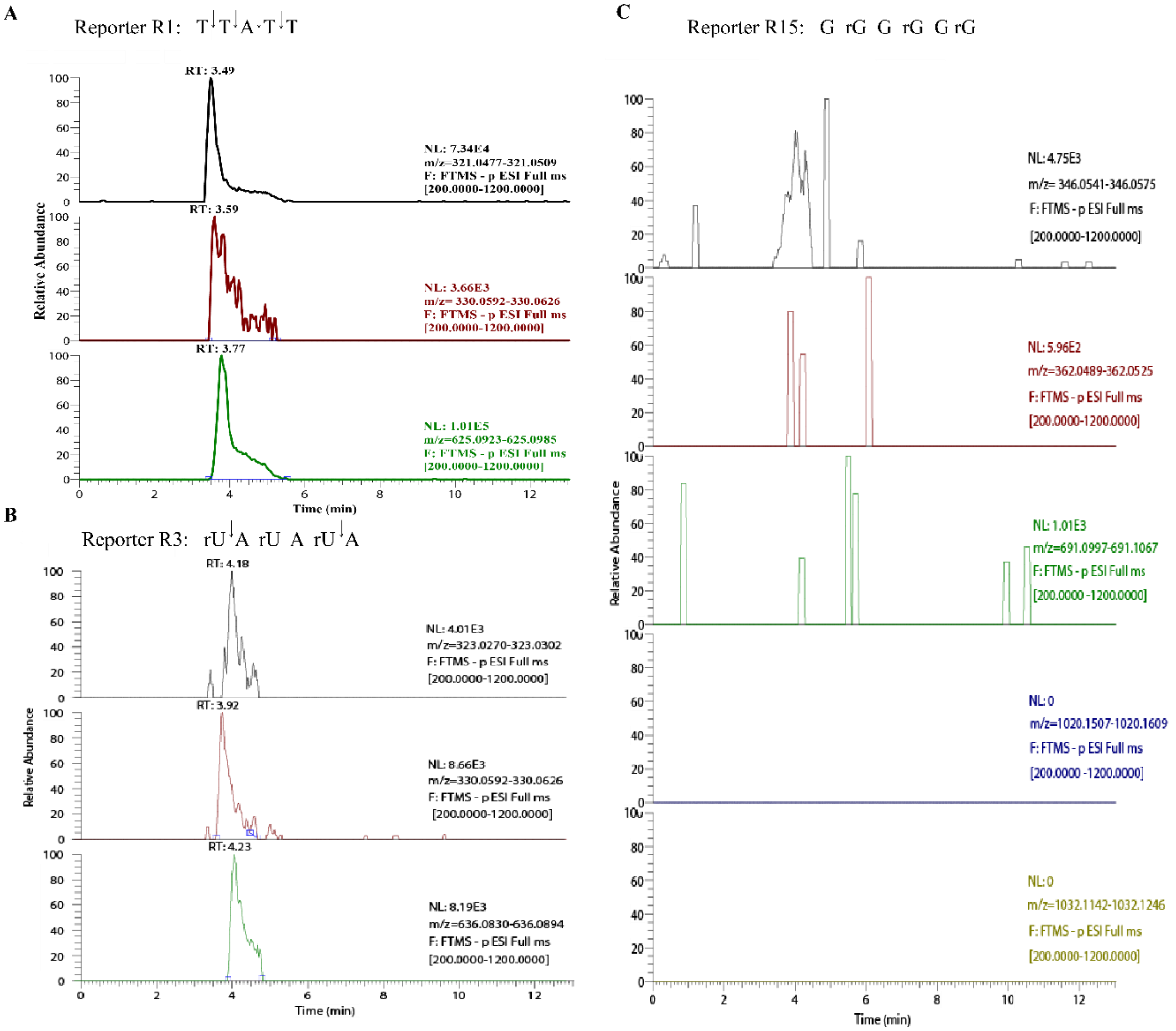

**Supplementary Fig. S8: Electrospray ionization mass spectrometry (ESI-MS) analysis of RAPID *trans cleavage* products from canonical and chimeric reporters for <u>AsCas12a treated samples.</u>** **(A)** *Trans cleavage* products of the canonical ssDNA reporter R1 (TTATT) using AsCas12a. Distinct peaks were observed at *m/z* 321, 330, and 625, corresponding to mononucleotides T ($C_{10}H_{15}N_2O_8P$) and A ($C_{10}H_{14}N_5O_6P$); and dinucleotide TT ($C_{20}H_{28}N_4O_{15}P_2$), respectively. These peaks were detected at retention times (RT) of 3.49 s (T), 3.59 s (A), and 3.77 s (TT). Other expected fragments (e.g., TTA, TA) were not detected (data not shown), indicating preferential cleavage patterns. **(B)** *Trans cleavage* products of the chimeric RNA/DNA reporter R3 (rUArUArUA). Peaks were observed at *m/z* 323 (rU; $C_9H_{13}N_2O_9P$), 330 (A; $C_{10}H_{14}N_5O_6P$), and 636 (rUA; $C_{19}H_{25}N_7O_{14}P_2$), corresponding to RT values of 4.18 s, 3.92 s, and 4.23 s, respectively. Similar to R1, certain intermediate fragments (e.g., rUArU, ArUA) were not detected, suggesting defined cleavage preferences and reduced fragmentation complexity. **(C)** Analysis of the poly(G)-rich reporter R15 (GrGGrGGrG) showed no detectable cleavage products. Expected fragments, including G (*m/z* 346), rG (*m/z* 362), GrG (*m/z* 691), GrGG (*m/z* 1020), and rGGrG (*m/z* 1032), were absent, consistent with the lack of fluorescence signal observed for poly(G)-based reporters. Across reporters, the detected cleavage products are consistent with Cas12a-mediated *trans cleavage* activity and reveal reporter-dependent fragmentation profiles. Notably, compared to canonical ssDNA reporters commonly used in DETECTR-like systems, the chimeric reporter exhibits reduced background fragmentation while maintaining defined cleavage products, supporting its improved signal-to-noise characteristics. Graph annotations: NL, maximum intensity; RT, retention time (s); *m/z*, mass-to-charge ratio; FTMS, Fourier transform mass spectrometry. Downward arrows indicate inferred cleavage positions. Peak tailing is attributed to phosphate groups and buffer effects.

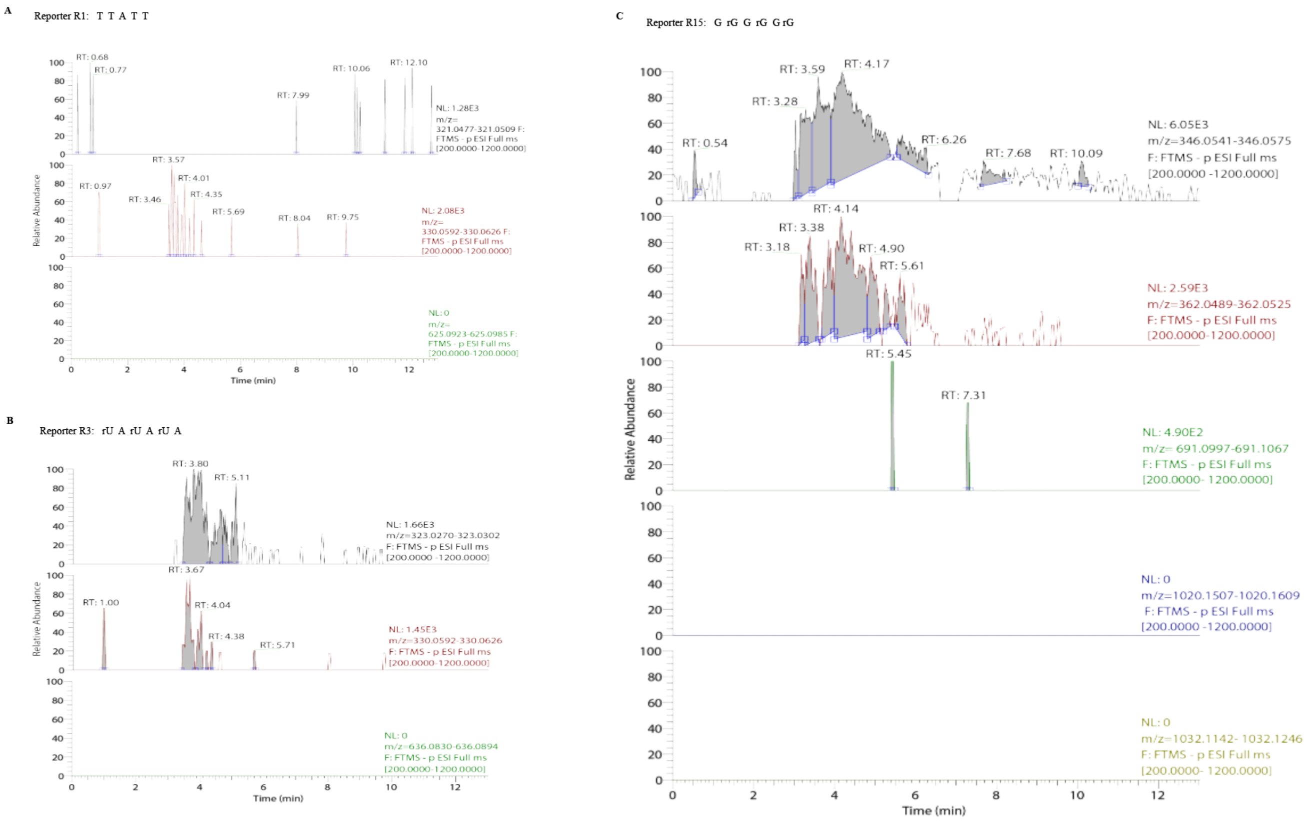

**Supplementary Fig. S9 | ESI-MS analysis of RAPID reporters in the absence of AsCas12a (negative control). (A)** Mass spectrometry analysis of the canonical ssDNA reporter R1 (TTATT) in the absence of AsCas12a. No detectable cleavage products were observed, in contrast to the AsCas12-treated condition (Supplementary Fig. 7). Specifically, no peaks corresponding to mononucleotides T (*m/z* 321; $C_{10}H_{15}N_2O_8P$) and A (*m/z* 330; $C_{10}H_{14}N_5O_6P$), or dinucleotide TT (*m/z* 625; $C_{20}H_{28}N_4O_{15}P_2$), were detected. The absence of these fragments confirms that reporter degradation is dependent on Cas12a-mediated trans-cleavage rather than spontaneous hydrolysis or buffer-induced effects. **(B)** Analysis of the chimeric RNA/DNA reporter R3 (rUArUArUA) without AsCas12a treatment. No fragment peaks were detected at *m/z* 323 (rU; $C_9H_{13}N_2O_9P$), 330 (A; $C_{10}H_{14}N_5O_6P$), or 636 (rUA; $C_{19}H_{25}N_7O_{14}P_2$), all of which are observed upon Cas12 treatment. These results further confirm that cleavage of the chimeric reporter is enzyme-dependent and not due to intrinsic instability of RNA/DNA hybrid sequences. **(C)** Mass spectrometry analysis of the poly(G)-rich reporter R15 (GrGGrGGrG) in the absence of AsCas12a. No detectable fragment peaks were observed, including those corresponding to G (*m/z* 346), rG (*m/z* 362), GrG (*m/z* 691), GrGG (*m/z* 1020), and rGGrG (*m/z* 1032). This is consistent with the lack of fluorescence signal observed for poly(G)-based reporters and confirms the absence of non-specific degradation.

Collectively, these negative control experiments demonstrate that reporter fragmentation is strictly dependent on Cas12a activity, thereby validating that the cleavage products observed in enzyme-treated samples arise from bona fide trans-cleavage events rather than background chemical degradation or instrument artifacts. Graph annotations: NL, maximum intensity; RT, retention time (s); *m/z*, mass-to-charge ratio; FTMS, Fourier transform mass spectrometry.

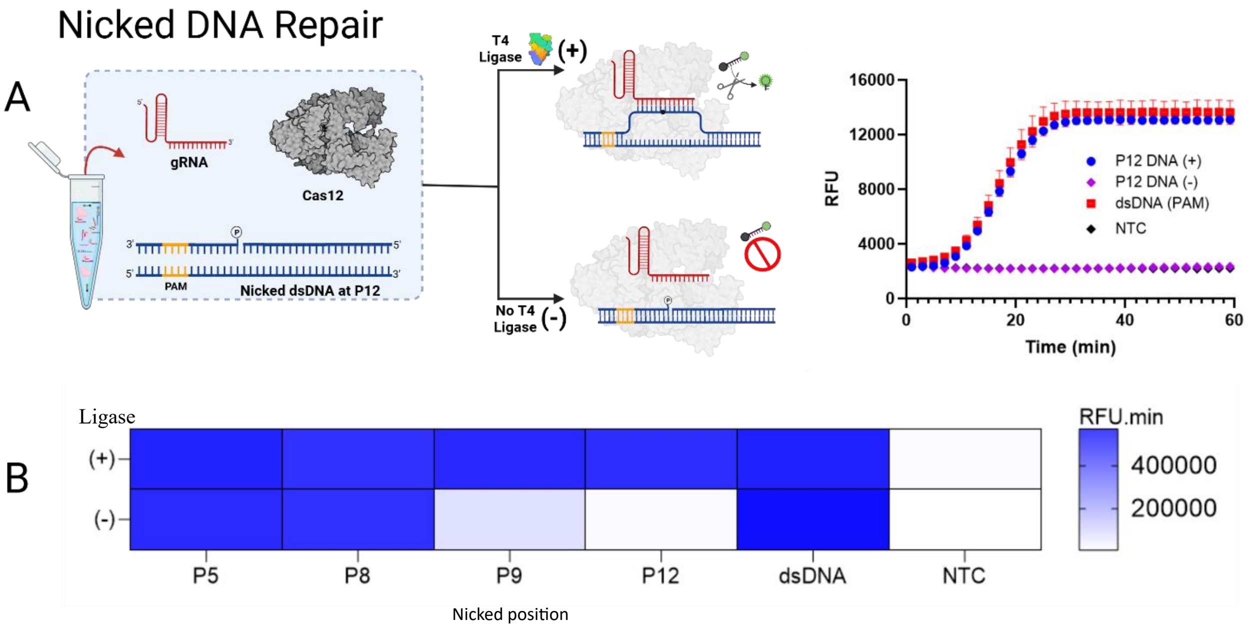

**Supplementary Fig. S10**: **DNA Ligation-Induced *Trans* Cleavage.** (A) Schematic representation of nicked DNA repair using T4 DNA ligase. Here, the three DNA strands are assembled to form a nick on the target strand. The 5'-end of the nicked DNA is phosphorylated, enabling repair in the presence of ligase, which restores the *trans* cleavage signal upon activation. Without the ligase enzyme, no *trans* cleavage is observed. (+) indicates the addition of ligase enzyme, while (-) denotes its absence. NTC (no target control) behaves similarly to (-) at P12. PAM-containing dsDNA is used as a positive control in comparison to the ligated P12 substrate. (B) Heat map depicting ligation activity of nicked DNA at various positions. Positions P5 and P8 demonstrate complete activation regardless of ligase presence, corresponding to the primary discovery from P1 to P20. Position P9 exhibits partial activation without ligase, which is fully restored upon ligase addition. At position P12, no activation occurs without ligase; however, the signal is recovered with ligase. dsDNA serves as a control template without a nick, and NTC represents the no-target control. RFU.min represents the area under the curve calculated from kinetic data within 60 minutes. Data represent mean ± standard deviation from n = 3 technical replicates.

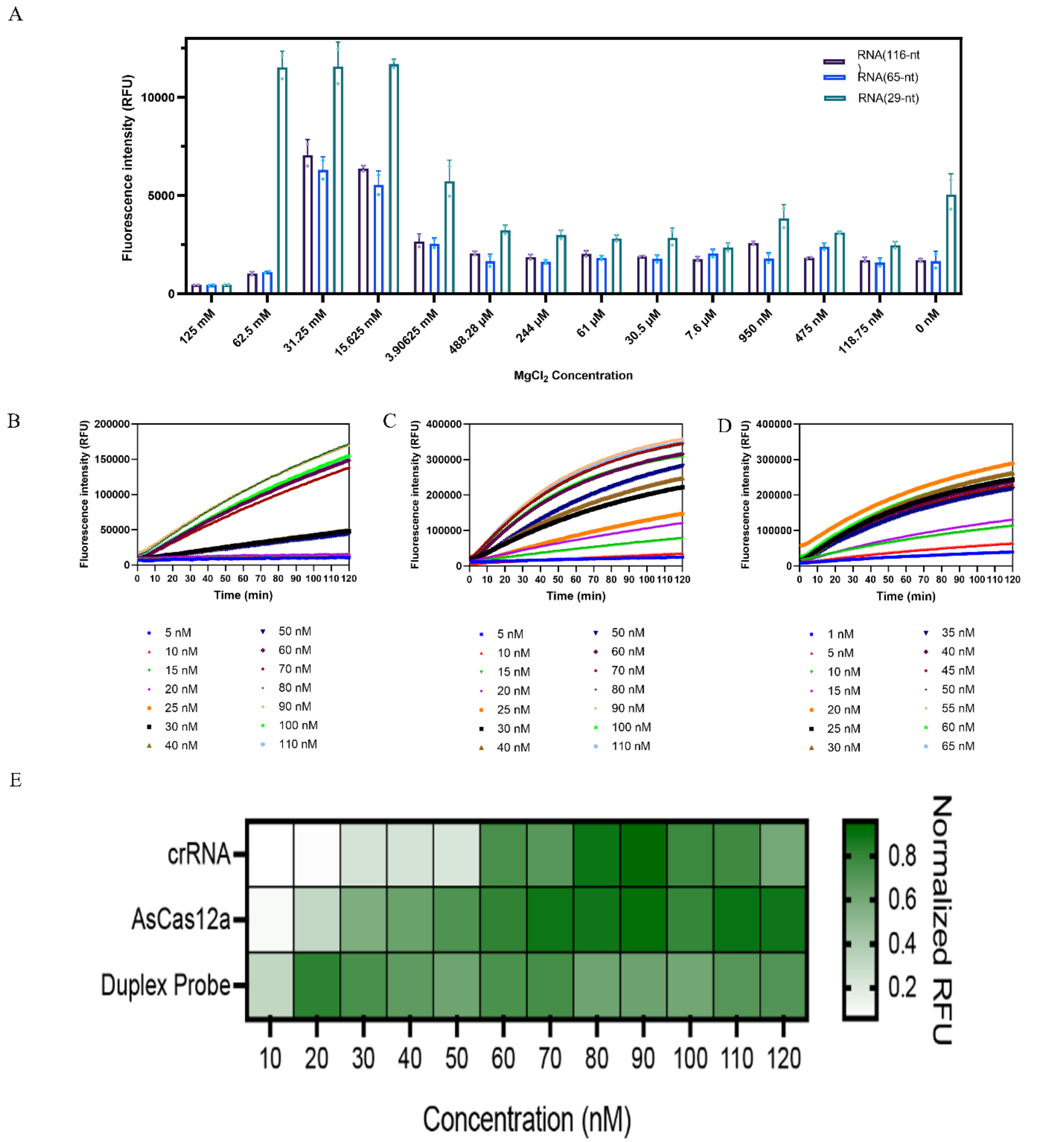

**Supplementary Fig. S11: RAPID optimized for miRNA detection using AsCas12a. (A)** Optimization of $MgCl_2$ concentrations for different lengths of RNA (29-nt, 65-nt, and 116-nt). The concentration of $MgCl_2$ ranged from 0 to 125 mM, with the optimal concentration identified at 31.25 mM. **(B), (C), and (D)** are kinetic plots showing the optimization of gRNA, AsCas12a, and duplex DNA probe, respectively. The optimal concentrations were determined to be 90 nM for gRNA, 90 nM for AsCas12a, and 20 nM for the duplex DNA probe. **(E)** Heatmap displaying the optimization of gRNA, AsCas12a, and PAM duplex concentrations, with optimal concentrations identified as 90 nM, 90 nM, and 20 nM, respectively.

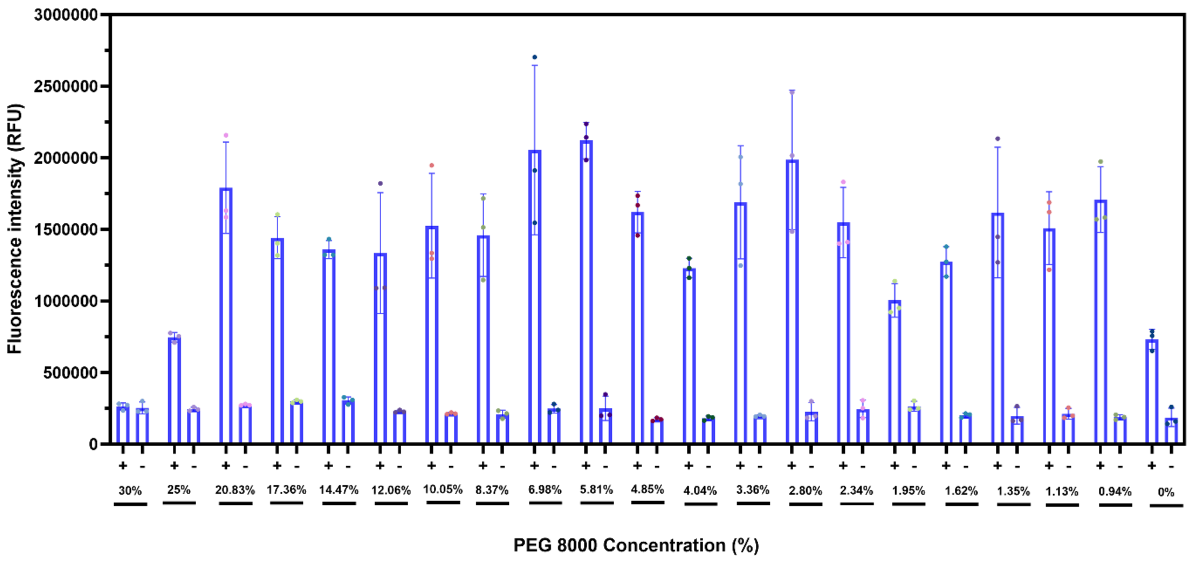

**Supplementary Fig. S12.** Bar graphs illustrating the optimization of PEG8000 concentrations, ranging from 0 to 30%, with the optimal concentration at 5.81%. The (+) and (-) signs represent reactions with and without the miRNA trigger.

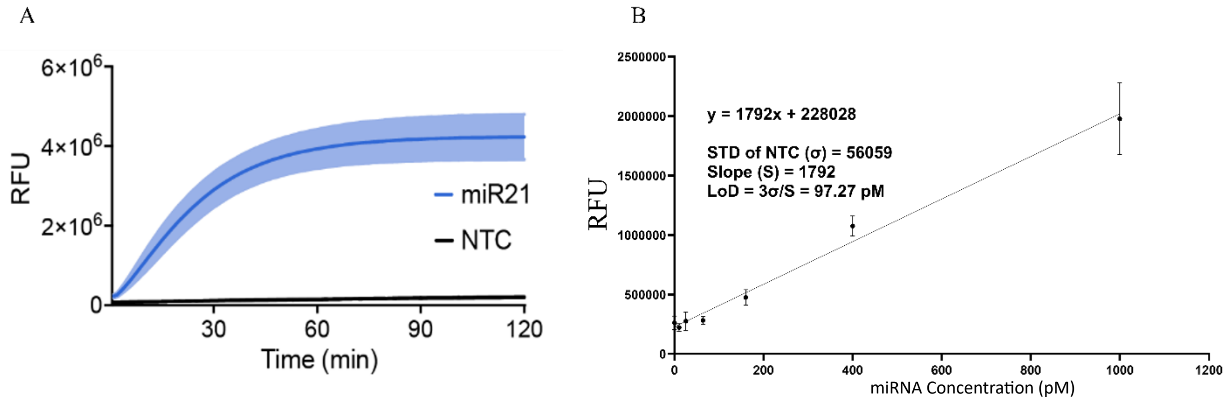

**Supplementary Fig. S13**: **(A)** Kinetics of miRNA-21 detection using RAPID, analyzed over a 120-minute period. (**B)** Limit of detection of miRNA-21 calculated from 3σ/S, where σ is the standard deviation of the background signal and S is the slope of the fluorescence of target concentration line.

A

B

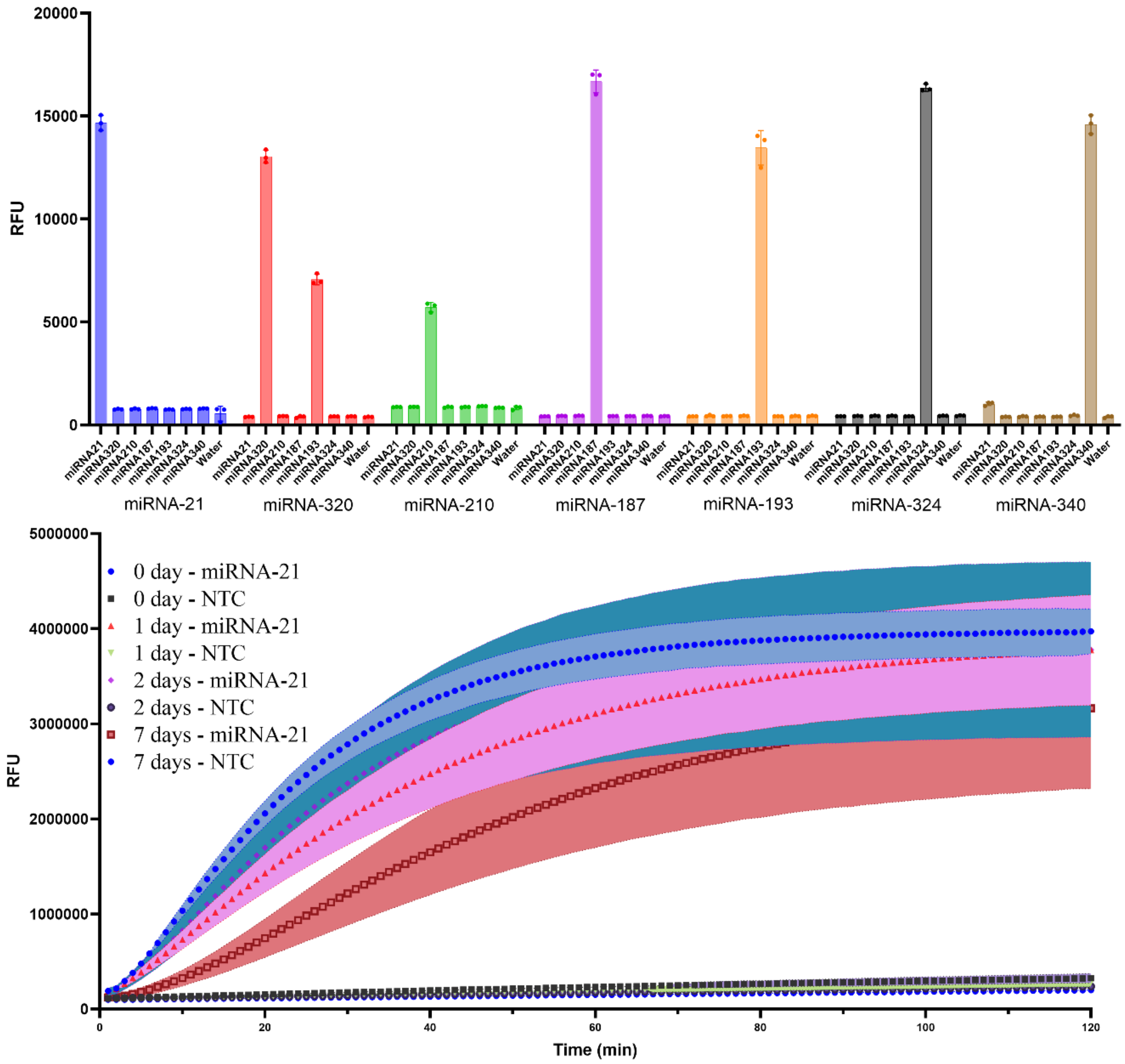

**Supplementary Fig. S14**: **(A)** Orthogonality testing of RAPID for miRNAs- 21, 320, 210, 187, 193, 324, and 340. The data, plotted after 60 mins, shows 100% orthogonality except for miRNAs-320 and 193 with very close similarity, confirming the system's specificity. **(B)** Kinetic plot for freeze-drying stability of miRNA-21 over 7 days. The results indicate that RAPID for miRNA detection is very stable, although stability can be further enhanced for longer storage to facilitate field deployability.

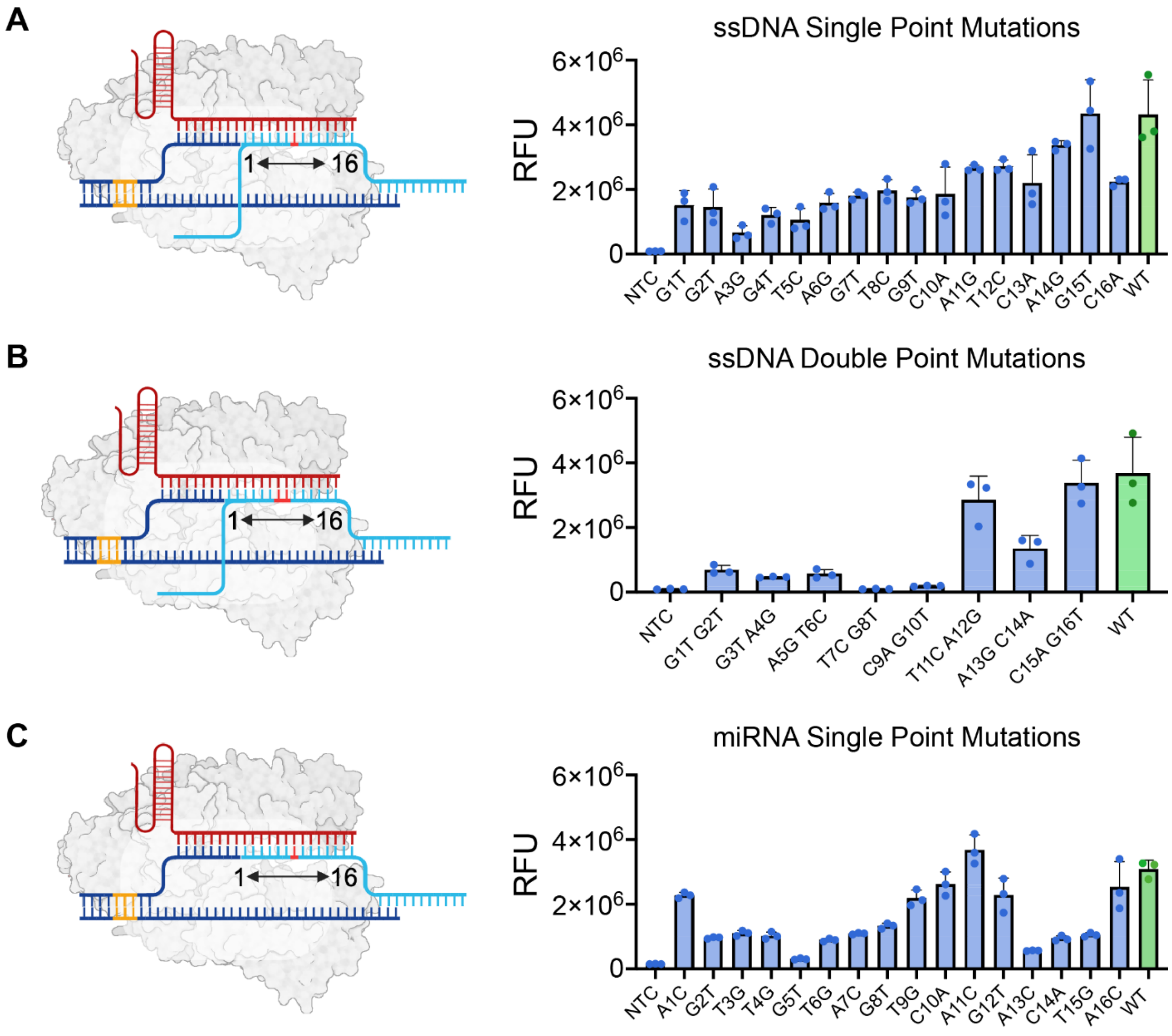

**Supplementary Fig. S15: Evaluating the Ability of RAPID to Detect Mismatches in DNA and RNA after 60 min reaction time. (A) Detection of Single-Point Mutations in ssDNA:** The left panel shows schematics of the point mutation, and the right panel displays results after 60 minutes of reaction. **(B) Detection of Two-Base Mismatches in ssDNA:** Schematics of the mismatches are on the left, with corresponding detection results on the right after 60 min. **(C) Detection of Single-Point Mutation in miRNA-21:** RAPID targets a 16-nt region within the protospacer of miRNA-21, with the remainder binding to duplex DNA. (**Method 11**) Green bars represent the wild type, blue bars represent mutants and non-target controls (NTCs). Results are expressed as mean ± SD (n = 3), measured in relative fluorescence units (RFU). Bar numbering corresponds to mutation sites (e.g., G1T indicates guanine mutated to thymine at position 1, nearest to the *trans* PAM, involving a switch from purines to pyrimidines and vice versa). NTC refers to no template control. **Note:** In the sequence positioning, nucleotide 1 is proximal to the trans PAM while nucleotide 16 is distal. All experiments were conducted relative to the wild-type sequence and included negative controls for comparison. The sequences used in these experiments are detailed in **Table S7**. Final DNA concentration is 2 uM, while miRNA is 50 nM.

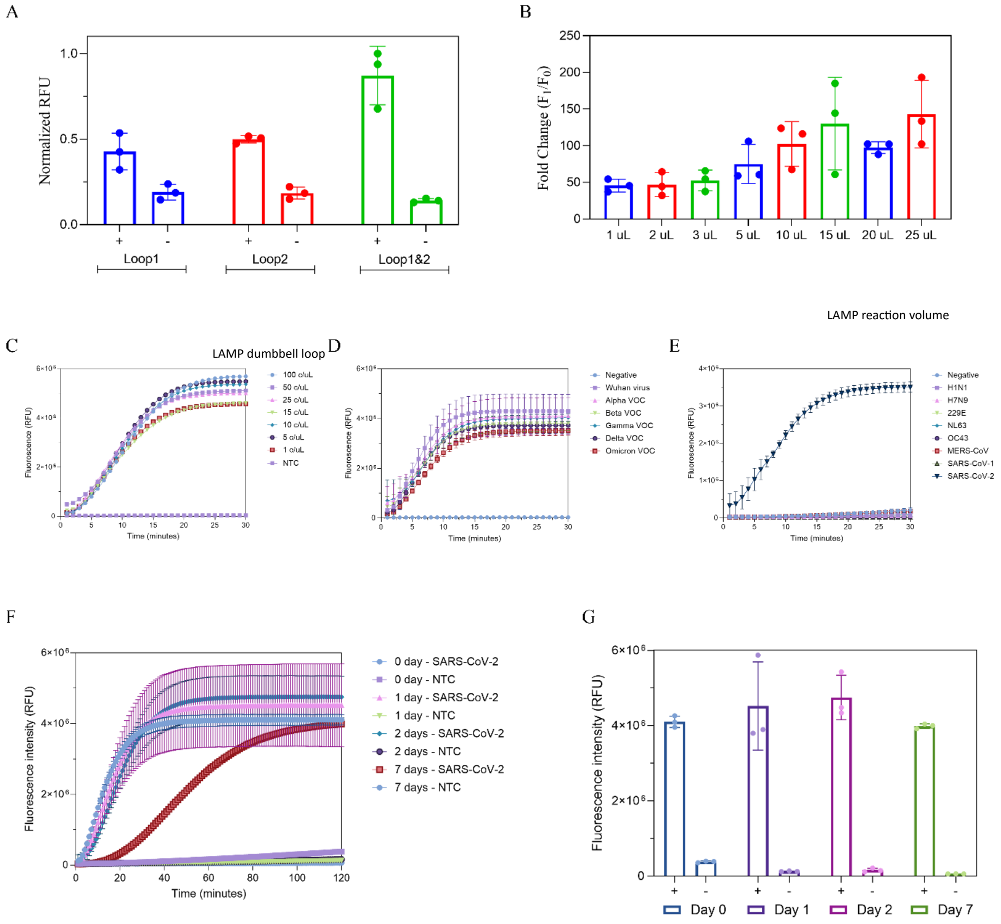

**Supplementary Fig. S16**: **Coupling of RAPID with LAMP for viral RNA detection. (A)** RAPID with ssDNA strands at a 1 nM concentration that mimic the loops of SARS-CoV-2 LAMP dumbbells. The results show that targeting the two dumbbells can significantly enhance detection. **(B)** Bar plots showing the volume of LAMP reactions spiked into the RAPID reaction mix. The results indicate no inhibition of the *trans*-cleavage signal up to 25 µL for the SARS-CoV-2 target, which was chosen for further RAPID-LAMP experiments. **(C)** Original time-course fluorescence signal for varying concentrations of the SARS-CoV-2 synthetic target. RAPID-LAMP demonstrates robustness, detecting down to 0.1 copies/µL within 5 minutes. **(D)** Time-dependent graph of RAPID-LAMP detecting various SARS-CoV-2 variants, including Wuhan virus, Alpha VOC, Beta VOC, Gamma VOC, Delta VOC, and Omicron VOC. **(E)** Kinetic plot of RAPID-LAMP's selectivity against various viral RNAs, including H1N1, H7N9, 229E, NL63, OC43, MERS-CoV, SARS-CoV-1, and SARS-CoV-2. The system showed 100% selectivity. **(F)** Time-course fluorescence signal of the stability test of the RAPID-LAMP assay. The components were freeze-dried and rehydrated with water, showing good stability up to 7 days, demonstrating the potential for field application of the RAPID-LAMP technology. Further testing is required to determine stability for at least a 1-year shelf life. **(G)** Plot showing fluorescence intensity of the freeze-dried RAPID-LAMP assay after 2 hours, demonstrating robust stability of the developed tool.

**Table S1:** gRNA sequence and DNA oligos used for proof of concept (see Fig.1)

| Name | Fragment | Sequences (5′-3′) | Purification |
|---|---|---|---|
| gRNA_1 | N/A | UAAUUUCUACUAAGUGUAGAUUGAAGUAGAUAU GGCAGCAC | HPLC |
| DNA Reporter (R1) | N/A | /56-FAM/TTATT/31ABkFQ | HPLC |
| NTS_01 | N/A | GAAGTTCAT GTTTCTGAA GTAGATATGGCAGCAC TAATCTA ATATG | Standard Desalting |
| TS_01_P1 | Fragment A | ACATGAACTTC | Standard Desalting |
| | Fragment B | CATATTAGATTAGTGCTGCCATATCTACTTCAGAA | |
| TS_01_P2 | Fragment A | AACATGAACTTC | Standard Desalting |
| | Fragment B | CATATTAGATTAGTGCTGCCATATCTACTTCAGA | |
| TS_01_P3 | Fragment A | AAACATGAACTTC | Standard Desalting |
| | Fragment B | CATATTAGATTAGTGCTGCCATATCTACTTCAG | |
| TS_01_P4 | Fragment A | GAAACATGAACTTC | Standard Desalting |
| | Fragment B | CATATTAGATTAGTGCTGCCATATCTACTTCA | |
| TS_01_P5 | Fragment A | AGAAACATGAACTTC | Standard Desalting |
| | Fragment B | CATATTAGATTAGTGCTGCCATATCTA CTTC | |
| TS_01_P6 | Fragment A | CAGAAACATGAACTTC | Standard Desalting |
| | Fragment B | CATATTAGATTAGTGCTGCCATATCTA CTT | |
| TS_01_P7 | Fragment A | TCAGAAACATGAACTTC | Standard Desalting |
| | Fragment B | CATATTAGATTAGTGCTGCCATATCTA CT | |
| TS_01_P8 | Fragment A | TTCAGAAACATGAACTTC | Standard Desalting |
| | Fragment B | CATATTAGATTAGTGCTGCCATATCTA C | |
| TS_01_P9 | Fragment A | CTTCAGAAACATGAACTTC | Standard Desalting |
| | Fragment B | CATATTAGATTAGTGCTGCCATATCTA | |
| TS_01_P10 | Fragment A | A CTTCAGAAACATGAACTTC | Standard Desalting |
| | Fragment B | CATATTAGATTAGTGCTGCCAT ATCT | |
| TS_01_P11 | Fragment A | TA CTTCAGAAACATGAACTTC | Standard Desalting |
| | Fragment B | CATATTAGATTAGTGCTGCCAT ATC | |
| TS_01_P12 | Fragment A | CTA CTTCAGAAACATGAACTTC | Standard Desalting |
| | Fragment B | CATATTAGATTAGTGCTGCCAT AT | |
| TS_01_P13 | Fragment A | TCTA CTTCAGAAACATGAACTTC | Standard Desalting |
| | Fragment B | CATATTAGATTAGTGCTGCCAT A | |
| TS_01_P14 | Fragment A | ATCTA CTTCAGAAACATGAACTTC | Standard Desalting |
| | Fragment B | CATATTAGATTAGTGCTGCCAT | |
| TS_01_P15 | Fragment A | T ATCTACTTCAGAAACATGAACTTC | Standard Desalting |

| | | | |
|---|---|---|---|
| | Fragment B | CATATTAGATTAGTGCT GCCA | |
| TS_01_P16 | Fragment A | AT ATCTACTTCAGAAACATGAACTTC | Standard Desalting |
| | Fragment B | CATATTAGATTAGTGCT GCC | |
| TS_01_P17 | Fragment A | CAT ATCTACTTCAGAAACATGAACTTC | Standard Desalting |
| | Fragment B | CATATTAGATTAGTGCT GC | |
| TS_01_P18 | Fragment A | CCAT ATCTACTTCAGAAACATGAACTTC | Standard Desalting |
| | Fragment B | CATATTAGATTAGTGCT G | |
| TS_01_P19 | Fragment A | GCCAT ATCTACTTCAGAAACATGAACTTC | Standard Desalting |
| | Fragment B | CATATTAGATTAGTGCT | |
| TS_01_P20 | Fragment A | T GCCATATCTACTTCAGAAACATGAACTTC | Standard Desalting |
| | Fragment B | CATATTAGATTAGTGC | |
| TS_01_P21 | Fragment A | CT GCCATATCTACTTCAGAAACATGAACTTC | Standard Desalting |
| | Fragment B | CATATTAGATTAGTG | |
| TS_01_P22 | Fragment A | GCT GCCATATCTACTTCAGAAACATGAACTTC | Standard Desalting |
| | Fragment B | CATATTAGATTAGT | |
| TS_01_P23 | Fragment A | TGCT GCCATATCTACTTCAGAAACATGAACTTC | Standard Desalting |
| | Fragment B | CATATTAGATTAG | |
| TS_01_P24 | Fragment A | GTGCT GCCATATCTACTTCAGAAACATGAACTTC | Standard Desalting |
| | Fragment B | CATATTAGATTA | |

**Table S2**: Nucleic acid oligos for nick testing on the non-target strand

| Name | Fragment | Sequences (5′-3′) | Purification |
|---|---|---|---|
| gRNA_1 | N/A | UAAUUUCUACUAAGUGUAGAUUGAAGUAGAU AUGGCAGCAC | HPLC |
| DNA Reporter (R1) | N/A | /56-FAM/TTATTT/31ABkFQ | HPLC |
| HPV TS | N/A | CATATTAGATTA GTGCTGCCATATCTACTTCAGAAA CATGAACTTC | Standard Desalting |
| NTS_01_S9 | Fragment A | GAAGTTCATGTTTCTGAA | Standard Desalting |
| | Fragment B | GTAGATATGGCAGCACTAATCTAATATG | |
| NTS_01_S11 | Fragment A | GAAGTTCATGTTTCTGAAGT | Standard Desalting |

| | | | |
|---|---|---|---|
| | Fragment B | AGATATGGCAGCACTAATCTAATATG | |
| NTS_01_S16 | Fragment A | GAAGTTCATGTTTCTGAAGTAGATA | Standard Desalting |
| | Fragment B | TGGCAGCACTAATCTAATATG | |

**Table S3:** gRNA sequence and DNA oligos used for proof of concept 2 (see Supplementary Fig.S1G and S4B)

| Name | Fragment | Sequences (5′-3′) | Purification |
|---|---|---|---|
| gRNA_2 | N/A | UAAUUUCUACUAAGUGUAGAU ACUUGCAAUGGACCAUAUCC | HPLC |
| DNA Reporter | N/A | /56-FAM/TTATTT/31ABkFQ | HPLC |
| NTS_02 | N/A | GAAGTTCAT GTTTCACTTGCAATGGACCATATCCTAATCTAATATG | Standard Desalting |
| TS-02 | NA | CATATTAGATTAGGATATGGTCCATTGCAAGTGAAACATGAACTTC | Standard Desalting |
| TS_02_P5 | Fragment A | TGAAACATGAACTTC | Standard Desalting |
| | Fragment B | CATATTAGATTAGGATATGGTCCATTGCAAG | |
| TS_02_P8 | Fragment A | AAGTGAAACATGAACTTC | Standard Desalting |
| | Fragment B | CATATTAGATTAGGATATGGTCCATTGC | |
| TS_02_P9 | Fragment A | CAAGTGAAACATGAACTTC | Standard Desalting |
| | Fragment B | CATATTAGATTAGGATATGGTCCATTG | |
| TS_02_P10 | Fragment A | GCAAGTGAAACATGAACTTC | Standard Desalting |
| | Fragment B | CATATTAGATTAGGATATGGTCCATT | |
| TS_02_P11 | Fragment A | TGCAAGTGAAACATGAACTTC | Standard Desalting |
| | Fragment B | CATATTAGATTAGGATATGGTCCAT | |
| TS_02_P12 | Fragment A | TTGCAAGTGAAACATGAACTTC | Standard Desalting |
| | Fragment B | CATATTAGATTAGGATATGGTCCA | |
| TS_02_P14 | Fragment A | CATTGCAAGTGAAACATGAACTTC | Standard Desalting |
| | Fragment B | CATATTAGATTAGGATATGGTC | |
| TS_02_P15 | Fragment A | CCATTGCAAGTGAAACATGAACTTC | Standard Desalting |
| | Fragment B | CATATTAGATTAGGATATGGT | |
| TS_02_P16 | Fragment A | TCCATTGCAAGTGAAACATGAACTTC | Standard Desalting |
| | Fragment B | CATATTAGATTAGGATATGG | |
| TS_02_P20 | Fragment A | ATGGTCCATTGCAAGTGAAACATGAACTTC | Standard Desalting |
| | Fragment B | CATATTAGATTAGGAT | |
| TS_02_P22 | Fragment A | ATATGGTCCATTGCAAGTGAAACATGAACTTC | Standard Desalting |
| | Fragment B | GCATATTAGATTAGG | |

**Table S4:** gRNA sequence and DNA oligos used for proof of concept 3 (see Fig.S4C)

| Name | Fragment | Sequences (5′-3′) | Purification |
|---|---|---|---|
| gRNA_3 | N/A | UAAUUUCUACUAAGUGUAGAUCGUCGCCGUCCAGCUCGACC | IVT |
| DNA Reporter | N/A | /56-FAM/TTATTT/31ABkFQ | HPLC |

| | | | |
|---|---|---|---|
| NTS_03 | N/A | CTTGTGGCCGTTTA CGTCGCCGTCCAGCTCGACC AGGATGGGCACC | Standard Desalting |
| TS-03 | NA | GGTGCCCATCCTGGTCGAGCTGGACGGCGACGTAAACGGCCACAAG | Standard Desalting |
| TS_03_P5 | Fragment A | GTAAA CGGCCACAAG | Standard Desalting |
| | Fragment B | GGTGCCCATCCTGGTCGAGCTGGACGGCGAC | |
| TS_03_P9 | Fragment A | CGA CGTAAACGGCCACAAG | Standard Desalting |
| | Fragment B | GGTGCCCATCCTGGTCGAGCTGGACGG | |
| TS_03_P12 | Fragment A | C GGCGACGTAAACGGCCACAAG | Standard Desalting |
| | Fragment B | GGTGCCCATCCTGGTCGAGCTGGA | |

**Table S5**: Structure of AsCas12 in complex with gRNA and target DNA.[1] PDB ID: 5B43

| Nick Position | Interacting amino acid (domain) |
|---|---|
| P9 | S186 (REC1), F1052 (RuvC-II) |
| P11 | Q1014 (RuvC-II) |
| P12 | I964 (RuvC-II) |
| P13 | R951 (BH), R955 (BH) |
| P14 | K524 (REC2), R955 (BH) |
| P19 | N278 (REC1), R301 (REC1) |
| P20 | N282 (REC1) |

**Table S6**: All nucleic acid used for testing RAPID against ssDNA, RNA, and dsDNA (Fig. 2)

| Name | Sub-Template | Sequences (5′-3′) | Type | Production | Purification |
|---|---|---|---|---|---|
| AsCas12a gRNA | T7_Template | GCGCTAATACGACTCACTAT AGGGTAATTTCTACTCTTG TAGATTGAAGTAGATATGGCAGCAC | ssDNA | PCR and IVT | Standard desalting |
| | FW_Prime | GCGCTAATACGACTCAC | ssDNA | | |
| | Rev_Primer | GTGCTGCCATATCTACTTC | ssDNA | | |
| DNA Reporter (R1) | Single | /56-FAM/TTATTT/31ABkFQ | | IDT | HPLC |
| LbCas12a gRNA | N/A | UAAUUUCUACUAAGUGUAGAU UGAAGUAGAUAUGGCAGCAC | RNA | IDT | HPLC |
| miRNA-28 nt | N/A | CAUAUUAGAUUAGUGCUGCCAUAUC UA C | RNA | IDT | HPLC |
| RNA-65 nt | T7_Template | GCGCTAATACGACTCACTATAG GGGCTGTCATTGATGCATATTA GATTAGTGCTGCCATATCTACA GGTCGACTTTCAAGAATTCATAT | ssDNA | PCR and IVT | Standard desalting |
| | FW_Prime | GCGCTAATACGACTCAC | ssDNA | | |
| | Rev_Primer | ATATGAATTCTTGAAAGTCGACC | ssDNA | | |

| | | | | | |
|---|---|---|---|---|---|
| RNA-116 nt | T7_Template | GCGCTAATACGACTCACTAT<br>AGGGTGCAAACACCATCAAGA<br>ATATCAAAGATAAAGCTGTCAT<br>TGATGAACATATTAGATT<br>AGTGCTGCCATATCTACAGG<br>TCGACTTTCAAGAATTCAT<br>ATCCCTGGTAGCCATTGCG | gBlock | PCR and IVT | Standard desalting |
| | FW_Prime | GCGCTAATACGACTCAC | ssDNA | | |
| | Rev_Primer | CGCAATGGCTACCAG | ssDNA | | |
| dsDNA-52 bp | DNA Template | GCGCTAATACGACTCACTATAGGGCA<br>TATTAGATTAGTGCTGCCATATCTAC | ssDNA | PCR | Standard desalting |
| | FW_Prime | GCGCTAATACGACTCAC | ssDNA | | |
| | Rev_Primer | GTAGATATGGCAGCACTAATC | ssDNA | | |
| dsDNA- 139 bp | DNA Template | GCGCTAATACGACTCACTATAG<br>GGTGCAAACACCATCAAGAAT<br>ATCAAAGATAAAGCTGTCATT<br>GATGAACATATTAGATTAGT<br>GCTGCCATATCTACAGGTCGA<br>CTTTCAAGAATTCATATCCCTGGTA<br>GCCATTGCG | gBlock | PCR | Standard desalting |
| | FW_Prime | GCGCTAATACGACTCAC | ssDNA | | |
| | Rev_Primer | CGCAATGGCTACCAG | ssDNA | | |
| dsDNA with PAM | DNA template | GCGCTAATACGACTCACTATAGGG<br>TGCAAACACCATCAAGAATATC<br>AAAGATAAAGCTGTCATTGATGA<br>ACATATTAGATTAGTGCTGC<br>CATATCTACTTCAGAAACATG<br>AACTTCAGGTCGACTTTC<br>AAGAATTCATATCCCTGGTAGCCATTG<br>CG | gBlock | PCR | Standard desalting |
| | FW_Prime | GCGCTAATACGACTCAC | ssDNA | | |
| | Rev_Primer | CGCAATGGCTACCAG | ssDNA | | |
| ssDNA 28 nt | N/A | CATATTAGATTAGTGCTGCCATATCTA<br>C | ssDNA | N/A | Standard desalting |
| ssDNA 89 nt | N/A | GCGCTAATACGACTCACTATAG<br>GGGCTGTCATTGATGCATATT<br>AGATTAGTGCTGCCATATC<br>TACAGGTCGACTTTCAAGAATTCATAT | ssDNA | N/A | Standard desalting |
| ssDNA 139 nt | N/A | GCGCTAATACGACTCACTATAGG<br>GTGCAAACACCATCAAGAATAT<br>CAAAGATAAAGCTGTCATTGATG<br>AACATATTAGATTAGTGCTGCCATATC<br>TACAGGTCGACTTTCAAGA<br>ATTCATATCCCTGGTAGCCATTGCG | Ultramer | N/A | Standard desalting |
| Duplex Probe | Sense | GAAGTTCAT GTTTCTGAA GTAGA<br>TATGGCAGCAC TAATCTA ATATG | ssDNA | IDT Duplexing | Standard desalting |
| | Anti-sense | TTCAGAAACATGAACTTC | ssDNA | | |
| Universal PAM | sense | TGTTTCTGAA | ssDNA | IDT Duplexing | Standard desalting |
| | Anti-sense | TTCAGAAACA | ssDNA | | |

**Table S7:** Oligos and modified probes for reporter screening. See Fig. 3

| Name | Short form | Sub-Template | Sequences (5′-3′) | Purification |
|---|---|---|---|---|
| Reporter 1 | R1 | N/A | /56-FAM/TTATT/31ABkFQ | HPLC |
| Reporter 2 | R2 | N/A | /56-FAM/rUrUrArUrU/31ABkFQ | HPLC |
| Reporter 3 | R3 | N/A | /56-FAM/rUArUArUA/31ABkFQ | HPLC |
| Reporter 4 | R4 | N/A | /56-FAM/ArUArUArU/31ABkFQ | HPLC |
| Reporter5 | R5 | N/A | /56-FAM/TTTTTT/31ABkFQ | HPLC |
| Reporter 6 | R6 | N/A | /56-FAM/AAAAAA/31ABkFQ | HPLC |
| Reporter 7 | R7 | N/A | /56-FAM/CCCCCC/31ABkFQ | HPLC |
| Reporter 8 | R8 | N/A | /56-FAM/GGGGGG/31ABkFQ | HPLC |
| Reporter 9 | R9 | N/A | /56-FAM/AAAAAA/31ABkFQ | HPLC |
| Reporter 10 | R10 | N/A | /56-FAM/UUUUUU/31ABkFQ | HPLC |
| Reporter 11 | R11 | N/A | /56-FAM/CCCCCC/31ABkFQ | HPLC |
| Reporter 12 | R12 | N/A | /56-FAM/GGGGGG/31ABkFQ | HPLC |
| Reporter 13 | R13 | N/A | /56-FAM/ArAArAArA/31ABkFQ | HPLC |
| Reporter 14 | R14 | N/A | /56-FAM/TrUTrUTrU/31ABkFQ | HPLC |
| Reporter 15 | R15 | N/A | /56-FAM/GrGGrGGrG/31ABkFQ | HPLC |
| Reporter 16 | R16 | N/A | /56-FAM/CrCCrCCrC/31ABkFQ | HPLC |
| LbCas12a gRNA | N/A | N/A | UAAUUUCUACUAAGUGUAGAUUGAA GUAGAUAUGGCAGCAC | HPLC |
| AsCas12a gRNA | N/A | T7_Template | GCGCTAATACGACTCACTATAG GGTAATTTCTACTCTTGTAGATT GAAGTAGATATGGCAGCAC | Standard Desalting |
| | | FW_Prime | GCGCTAATACGACTCAC | |
| | | Rev_Primer | GTGCTGCCATATCTACTTC | |
| ssDNA 89 nt | Trigger | N/A | GCGCTAATACGACTCACTATAGGGGCT GTCATTGATGCATATTAGATTAGTGCTG CCATATCTACAGGTCGACTTTCAAGAAT TCATAT | Standard Desalting |
| Duplex Probe | N/A | Sense | GAAGTTCAT GTTTCTGAA GTAGATATGGCAGCAC TAATCTA ATATG | Standard Desalting |
| | | Anti-sense | TTCAGAAACATGAACTTC | |

**Table S8**: HPV 18-related sequences to validate chimeric probes.

| Name | Sequence (5’-3’) |
|---|---|
| HPV 18 | TTGTTACCTCTGACTCCCAGTTGTTTAATAAACCATATTGGTT ACATAAGGCACAGGGTCATAACAATGGTGTTTGCTGGCATA ATCAATTATTTGTTACTGTGGTAGATACCACTCCCAGTACCA ATTTAACAATATGTGCTTCTACACAGTCTCCTGTACCTGGGC AATATGATGCTACCAAATTTAAGCAGTATAGCAGACATGTT GAGGAATATGATTTGCAGTTTATTTTTCAGTTGTGTACTATT ACTTTAACTGCAGATGTTATGTCCTATATTCATAGTATGAAT AGCAGTA |
| R1 Reporter | /56-FAM/TTATT/3IABkFQ/ |
| R3 Reporter | /56-FAM/rUArUArUA/3IABkFQ/ |
| HPV 18-LbCas12 and AsCas12-spacer sequence | ACAAUAUGUGCUUCUACACA |

Table S9: Mass spectrometry–resolved cleavage profiles of RAPID reporters.

| Reporter RI: 5' – T-T-A-T-T-3' | | | | | |
|---|---|---|---|---|---|
| Fragments | Formula | m/z | RT | Positive control (AsCas12a treated) - Intensity | Negative control (No AsCas12a) |
| T | C10H15N2O8P | 321.0493 | 3.49 | 1.35E+06 | ND |
| A | C10H14N5O6P | 330.0609 | 3.59 | 1.40E+05 | ND |
| TT | C20H28N4O15P2 | 625.0954 | 3.77 | 3.04E+06 | ND |
| TTA | C30H40N9O20P3 | 938.153 | | ND | ND |
| TA | C20H27N7O13P2 | 634.1069 | | ND | ND |
| Reporter R3: 5' – rU-A-rU-A-rU-A -3' | | | | | |
| Fragments | Formula | m/z | RT | Positive control (AsCas12a treated) - Intensity | Negative control (No AsCas12a) |
| rU | C9H13N2O9P | 323.0286 | 4.18 | 5.29E+04 | ND |
| A | C10H14N5O6P | 330.0609 | 3.92 | 1.65E+05 | ND |
| rUA | C19H25N7O14P2 | 636.0862 | 4.23 | 1.92E+05 | ND |
| rUArU | C28H36N9O22P3 | 942.1115 | | ND | ND |
| ArUA | C20H37N14O19P3 | 869.1499 | | ND | ND |
| Reporter R15: 5' – G-rG-G-rG-G-rG -3' | | | | | |
| Fragments | Formula | m/z | RT | Positive control (AsCas12a treated) - Intensity | Negative control (No AsCas12a) |
| G | C10H14N5O7P | 346.0558 | | ND | ND |
| rG | C10H14N5O8P | 362.0507 | | ND | ND |
| GrG | C20H26N10O14P2 | 691.1032 | | ND | ND |
| GrGG | C30H38N15O20P3 | 1020.1558 | | ND | ND |
| rGGrG | C30H34N15O21P3 | 1032.1194 | | ND | ND |

**Table S10**: Sequences for RAPID proof-of-concept diagnostic for monkeypox dsDNA virus

| | Name | Sequence (5′–3′) | Reference | Source |
|---|---|---|---|---|
| **Primers** | b6r-RPA-fwd | GGAATGATACTGTCACGTGTCCTAATGCGG | Cheng *et al.* “b6r-RPA-1F” | IDT ssDNA oligo |
| | b6r-RPA-rev | CCGTACAACTTATATACGAAACACCAATAACC | Cheng *et al.* “b6r-RPA-1R” | |
| **crRNA** | CRISPR | UAAUUUCUACUAAGUGUAGAU**ACUGGUUGACACGAUCCGUG** | Cheng *et al.* “b6r-TTTA” | IDT Alt-R™ A.s. Cas12a crRNA |
| | RAPID | UAAUUUCUACUAAGUGUAGAU**ugaaAUGACUAUCAACUGUG** | this study | |
| **Target** | pUC57-b6r | 165115..166068* | GenBank: ON563414.3 | GenScript FLASH gene-to-plasmid |

** Genome region provide for sequences too long to display in table.*

**Table S11**: All sequences used for the nicked DNA repair and miRNA-DNA Ligation

| Name | Sub-Template | Sequences (5′-3′) | Production |
|---|---|---|---|
| **DNA Ligation** | | | |
| P5 DNA | Fragment A | 5'Phos AGAAACATGAACTTC | Standard (IDT) |
| | Fragment B | CATATTAGATTAGTGCTGCCATATCTA CTTC | |
| | NTS | GAAGTTCAT GTTTCTGAA GTAGATATGGCAGCAC TAATCTA ATATG | |
| P8 DNA | Fragment A | 5'Phos TTCAGAAACATGAACTTC | Standard (IDT) |
| | Fragment B | CATATTAGATTAGTGCTGCCATATCTA C | |
| | NTS | GAAGTTCAT GTTTCTGAA GTAGATATGGCAGCAC TAATCTA ATATG | |
| P9 DNA | Fragment A | 5'Phos CTTCAGAAACATGAACTTC | Standard (IDT) |
| | Fragment B | CATATTAGATTAGTGCTGCCATATCTA | |
| | NTS | GAAGTTCAT GTTTCTGAA GTAGATATGGCAGCAC TAATCTA ATATG | |
| P12 DNA | Fragment A | 5'Phos CTA CTTCAGAAACATGAACTTC | Standard (IDT) |
| | Fragment B | CATATTAGATTAGTGCTGCCAT AT | |
| | NTS | GAAGTTCAT GTTTCTGAA GTAGATATGGCAGCAC TAATCTA ATATG | |
| **miRNA Ligation** | | | |
| P5 RNA | Fragment A | 5'Phos AGAAACATGAACTTC | DNA - Standard (IDT) miRNA - IVT |
| | miRNA | CAUAUUAGAUUAGUGCUGCCAUAUCUA CUUC | |
| | NTS | GAAGTTCAT GTTTCTGAA GTAGATATGGCAGCAC TAATCTA ATATG | |
| P8 RNA | Fragment A | 5'Phos TTCAGAAACATGAACTTC | DNA - Standard (IDT) miRNA - Standard (IDT) |
| | miRNA | CAUAUUAGAUUAGUGCUGCCAUAUCUA C | |
| | NTS | GAAGTTCAT GTTTCTGAA GTAGATATGGCAGCAC TAATCTA ATATG | |
| P9 RNA | Fragment A | 5'Phos CTTCAGAAACATGAACTTC | DNA - Standard (IDT) miRNA - IVT |
| | miRNA | CAUAUUAGAUUAGUGCUGCCAUAUCUA | |
| | NTS | GAAGTTCAT GTTTCTGAA GTAGATATGGCAGCAC TAATCTA ATATG | |
| P12 RNA | Fragment A | 5'Phos CTA CTTCAGAAACATGAACTTC | DNA - Standard (IDT) miRNA - IVT |
| | miRNA | CAUAUUAGAUUAGUGCUGCCAU AU | |
| | NTS | GAAGTTCAT GTTTCTGAA GTAGATATGGCAGCAC TAATCTA ATATG | |

**Table S12**: Sequences for miRNA orthogonality testing

| Name | Fragment | Sequence |
|---|---|---|
| miR-193b-5p | | CGGGGUUUUGAGGGCGAGAUGA |

| | | |
|---|---|---|
| miR-187a-3p | | UCGUGUCUUGUGUUGCAGCCGG |
| hsa-miR-324-5p | | CGCAUCCCCUAGGGCAUUGGUG |
| hsa-miR-340-5p | | UUAUAAAGCAAUGAGACUGAUU |
| hsa-miR-21 | | UAGCUUAUCAGACUGAUGUUGA |
| hsa-miR-320a | | AAAAGCUGGGUUGAGAGGGCGA |
| has-miR-210-3p | | CUGUGCGUGUGACAGCGGCUGA |
| miRNA-193 | Duplex _193(sense) | GAAGTTCAT GTTTC TGAA TCATCTCGCCCTCAAA ACCCCG A ATATG |
| | Duplex_193 (antisense) | TTCAGAAACATGAACTTC |
| miRNA-187 | Duplex _187(sense) | GAAGTTCAT GTTTC TGAA CCGGCTGCAACACAAG ACACGA A ATATG |
| | Duplex_187 (antisense) | TTCAGAAACATGAACTTC |
| miRNA-324 | Duplex_324 (sense) | GAAGTTCAT GTTTC TGAA CACCAATGCCCTAGGG GATGCG A ATATG |
| | Duplex_324 (antisense) | TTCAGAAACATGAACTTC |
| miRNA-340 | Duplex_340 (sense) | GAAGTTCAT GTTTC TGAA AATCAGTCTCATTGCT TTATAA A ATATG |
| | Duplex_340 (antisense) | TTCAGAAACATGAACTTC |
| miRNA-21 | Duplex_21 (sense) | GAAGTTCAT GTTTCTGAA TCAACATCAGTCTGAT AAGCTA TTCAAG |
| | Duplex_21 (antisense) | TTCAGAAACATGAACTTC |
| miRNA-320 | Duplex_320 (sense) | GAAGTTCAT GTTTCTGAA TCGCCCTCTCAACCCA GCTTTT TTCAAG |
| | Duplex_320 (antisense) | TTCAGAAACATGAACTTC |
| miRNA-210 | Duplex_210 (sense) | GAAGTTCAT GTTTCTGAA TCAGCCGCTGTCACAC GCACAG TTCAAG |
| | Duplex_210 (antisense) | TTCAGAAACATGAACTTC |
| crRNA_193 | | /AltR1/rUrA rArUrU rUrCrU rArCrU rCrUrU rGrUrA rGrArU rUrGrA rArUrC rArUrC rUrCrG rCrCrC rUrCrA rArA/AltR2/ |
| crRNA_187 | | /AltR1/rUrA rArUrU rUrCrU rArCrU rCrUrU rGrUrA rGrArU rUrGrA rArCrC rGrGrC rUrGrC rArArC rArCrA rArG/AltR2/ |
| crRNA_324 | | /AltR1/rUrA rArUrU rUrCrU rArCrU rCrUrU rGrUrA rGrArU rUrGrA rArCrA rCrCrA rArUrG rCrCrC rUrArG rGrG/AltR2/ |
| crRNA_340 | | /AltR1/rUrA rArUrU rUrCrU rArCrU rCrUrU rGrUrA rGrArU rUrGrA rArArA rUrCrA rGrUrC rUrCrA rUrUrG rCrU/AltR2/ |
| crRNA_21 | | UAAUUUCUACUCUUGUAGAU UGAA UCAACAUCAGUCUGAU |

| | | |
|---|---|---|
| crRNA_320 | | /AltR1/rUrA rArUrU rUrCrU rArCrU rCrUrU rGrUrA rGrArU rCrGrC rCrCrU rCrUrC rArArC rCrCrA rGrCrU rUrU/AltR2/ |
| crRNA_210 | | UAAUUUCUACUCUUGUAGAU UGAA UCAGCCGCUGUCACAC |

*r indicates RNA bases

**Table S13**:Nucleic acid sequence for miRNA detection. See Fig. 5

| Name | Sub-Template | Sequences (5′-3′) | Production |
|---|---|---|---|
| miRNA-21 | T7-DNA Template | GCGCTAATACGACTCACTATAGGGTAGCTTA TCAGACTGATGTTGA | PCR_IVT |
| | FW_Primer | GCGCTAATACGACTCACTATAGGG | |
| | Rev_Primer | TCAACATCAGTCTGATAAGC | |
| miRNA-21 AsCas12a gRNA | T7-DNA Template | GCGCTAATACGACTCACTATAGGG TAATTTCTACTCTTGTAGAT TGAA TCAACATCAGTCTGAT | PCR_IVT |
| | FW_Primer | GCGCTAATACGACTCACTATAGGG | |
| | Rev_Primer | ATCAGACTGATGTTGATTCA | |
| miRNA-21 Duplex Probe | Sense | GAAGTTCAT GTTTCTGAA TCAACATCAGTCTGAT AAGCTA TTCAAG | IDT_Duplexing |
| | Anti Sense | TTCAGAAACATGAACTTC | |
| miRNA-320a | T7-DNA Template | GCGCTAATACGACTCACTATAGGG AAAAGCTGGGTTGAGAGGGCGA | PCR_IVT |
| | FW_Primer | GCGCTAATACGACTCACTATAGGG | |
| | Rev_Primer | TCGCCCTCTCAACCC | |
| miRNA-320a AsCas12a gRNA | T7-DNA template | GCGCTAATACGACTCACTATAGGG TAATTTCTACTCTTGTAGAT TGAA TCGCCCTCTCAACCCA | PCR_IVT |
| | FW_Primer | GCGCTAATACGACTCACTATAGGG | |
| | Rev_Primer | TGGGTTGAGAGGGCGATTC | |
| miRNA-320a Duplex Probe | Sense | GAAGTTCAT GTTTCTGAA TCGCCCTCTCAACCCA GCTTTT TTCAAG | IDT_Duplexing |
| | Anti Sense | TTCAGAAACATGAACTTC | |
| miRNA-210-3p | T7-DNA Template | GCGCTAATACGACTCACTATAGGG CTGTGCGTGTGACAGCGGCTGA | PCR_IVT |
| | FW_Primer | GCGCTAATACGACTCACTATAGGG | |
| | Rev_Primer | TCAGCCGCTGTCACA | |
| miRNA-210-3p AsCas12a gRNA | T7-DNA Template | GCGCTAATACGACTCACTATAGGG TAATTTCTACTCTTGTAGAT TGAA TCAGCCGCTGTCACAC | PCR_IVT |
| | FW_Primer | GCGCTAATACGACTCACTATAGGG | |
| | Rev_Primer | GTGTGACAGCGGCTGATTC | |
| miRNA-210-3p Duplex Probe | Sense | GAAGTTCAT GTTTCTGAA TCAGCCGCTGTCACAC GCACAG TTCAAG | IDT_Duplexing |
| | Anti Sense | TTCAGAAACATGAACTTC | |

| | | | |
|---|---|---|---|
| Chimeric Reporter | R3 | /56-FAM/rUArUArUA/31ABkFQ | IDT_HPLC |

*PCR_IVT means that PCR was performed to amplify the ssDNA substrate before performing in vitro reaction
**IDT_Duplexing means that the duplexing was done by IDT and shipped dry to us

#### **Table S14**: Nucleic acid sequences for DNA mismatched Screening.

| Name | Sub-Template | Sequences (5′-3′) | Production | Purification |
|---|---|---|---|---|
| **DNA Point Mutation** | | | | |
| WT | N/A | ACTGCTGCCTGGAGTTGAATTTCTTGAACTGTTGC**GACTACGTGATGAGG**AACGA | IDT probes | Standard |
| G1T | N/A | ACTGCTGCCTGGAGTTGAATTTCTTGAACTGTTGC**GACTACGTGATGAGT**AACGA | IDT probes | Standard |
| G2T | N/A | ACTGCTGCCTGGAGTTGAATTTCTTGAACTGTTGC**GACTACGTGATGATG**AACGA | IDT probes | Standard |
| A3G | N/A | ACTGCTGCCTGGAGTTGAATTTCTTGAACTGTTGC**GACTACGTGATGGGG**AACGA | IDT probes | Standard |
| G4T | N/A | ACTGCTGCCTGGAGTTGAATTTCTTGAACTGTTGC**GACTACGTGATTAGG**AACGA | IDT probes | Standard |
| T5C | N/A | ACTGCTGCCTGGAGTTGAATTTCTTGAACTGTTGC**GACTACGTGACGAGG**AACGA | IDT probes | Standard |
| A6G | N/A | ACTGCTGCCTGGAGTTGAATTTCTTGAACTGTTGC**GACTACGTGGTGAGG**AACGA | IDT probes | Standard |
| G7T | N/A | ACTGCTGCCTGGAGTTGAATTTCTTGAACTGTTGC**GACTACGTTATGAGG**AACGA | IDT probes | Standard |
| T8C | N/A | ACTGCTGCCTGGAGTTGAATTTCTTGAACTGTTGC**GACTACGCGATGAGG**AACGA | IDT probes | Standard |
| G9T | N/A | ACTGCTGCCTGGAGTTGAATTTCTTGAACTGTTGC**GACTACTTGATGAGG**AACGA | IDT probes | Standard |
| C10A | N/A | ACTGCTGCCTGGAGTTGAATTTCTTGAACTGTTGC**GACTAAGTGATGAGG**AACGA | IDT probes | Standard |
| A11G | N/A | ACTGCTGCCTGGAGTTGAATTTCTTGAACTGTTGC**GACTGCGTGATGAGG**AACGA | IDT probes | Standard |
| T12C | N/A | ACTGCTGCCTGGAGTTGAATTTCTTGAACTGTTGC**GACCACGTGATGAGG**AACGA | IDT probes | Standard |
| C13A | N/A | ACTGCTGCCTGGAGTTGAATTTCTTGAACTGTTGC**GAATACGTGATGAGG**AACGA | IDT probes | Standard |
| A14G | N/A | ACTGCTGCCTGGAGTTGAATTTCTTGAACTGTTGC**GGCTACGTGATGAGG**AACGA | IDT probes | Standard |
| G15T | N/A | ACTGCTGCCTGGAGTTGAATTTCTTGAACTGTTGC**TACTACGTGATGAGG**AACGA | IDT probes | Standard |
| C16A | N/A | ACTGCTGCCTGGAGTTGAATTTCTTGAACTGTTGA**GACTACGTGATGAGG**AACGA | IDT probes | Standard |
| **Paired Two DNA Base Mismatches** | | | | |
| WT | N/A | ACTGCTGCCTGGAGTTGAATTTCTTGAACTGTTGC**GACTACGTGATGAGG**AACGA | IDT Probes | Standard |
| G1T G2T | N/A | ACTGCTGCCTGGAGTTGAATTTCTTGAACTGTTGC**GACTACGTGATGATT**AACGA | IDT Probes | Standard |
| G3T A4G | N/A | ACTGCTGCCTGGAGTTGAATTTCTTGAACTGTTGC**GACTACGTGATTGGG**AACGA | IDT Probes | Standard |
| A5G T6C | N/A | ACTGCTGCCTGGAGTTGAATTTCTTGAACTGTTGC**GACTACGTGGCGAGG**AACGA | IDT Probes | Standard |
| T7C G8T | N/A | ACTGCTGCCTGGAGTTGAATTTCTTGAACTGTTGC**GACTACGCTATGAGG**AACGA | IDT Probes | Standard |
| C9A G10T | N/A | ACTGCTGCCTGGAGTTGAATTTCTTGAACTGTTGC**GACTAATTGATGAGG**AACGA | IDT Probes | Standard |
| T11C A12G | N/A | ACTGCTGCCTGGAGTTGAATTTCTTGAACTGTTGC**GACCGCGTGATGAGG**AACGA | IDT Probes | Standard |
| A13G C14A | N/A | ACTGCTGCCTGGAGTTGAATTTCTTGAACTGTTGC**GGATACGTGATGAGG**AACGA | IDT Probes | Standard |
| C15A G16T | N/A | ACTGCTGCCTGGAGTTGAATTTCTTGAACTGTTGA**TACTACGTGATGAGG**AACGA | IDT Probes | Standard |
| Deplex probe | Sense strand | GAAGTTCAT GTTTCTGAA CCTCATCACGTAGTCGCAACAGTTCAAG | IDT Duplexing | Standard |
| | Anti Sense strand | TTCAGAAACATGAACTTC | | |
| LbCas12a gRNA | T7_Template | GCGCTAATACGACTCACTATAGGG TAATTTCTACTAAGTGTAGAT TGAA CCTCATCACGTAGTCG | PCR_IVT | Standard |
| | FW_Primer | GCGCTAATACGACTCAC | | |
| | Rev_Primer | CGACTACGTGATGAGGT | | |

*The region in blue is part of the protospacer region that binds to the gRNA
**The sequences highlighted in red represent the mismatches.
***IVT denotes *in vitro* transcription

**Table S15**: Oligos for screening point mutations in miRNA-21

| Name | Sub-Template | Sequences (5′-3′) | Production |
|---|---|---|---|
| WT | N/A | GCGCTAATACGACTCACTATAGGG **TAGCTT** **ATCAGACTGATGTTGA** | IVT-IDT Probes |
| A1C | N/A | GCGCTAATACGACTCACTATAGGG **TAGCTT** **ATCAGACTGATGTTGg** | IVT-IDT Probes |
| G2T | N/A | GCGCTAATACGACTCACTATAGGG **TAGCTT** **ATCAGACTGATGTTtA** | IVT-IDT Probes |
| T3G | N/A | GCGCTAATACGACTCACTATAGGG **TAGCTT** **ATCAGACTGATGTcGA** | IVT-IDT Probes |
| T4G | N/A | GCGCTAATACGACTCACTATAGGG **TAGCTT** **ATCAGACTGATGcTGA** | IVT-IDT Probes |
| G5T | N/A | GCGCTAATACGACTCACTATAGGG **TAGCTT** **ATCAGACTGATtTTGA** | IVT-IDT Probes |
| T6G | N/A | GCGCTAATACGACTCACTATAGGG **TAGCTT** **ATCAGACTGAcGTTGA** | IVT-IDT Probes |
| A7C | N/A | GCGCTAATACGACTCACTATAGGG **TAGCTT** **ATCAGACTGgTGTTGA** | IVT-IDT Probes |
| G8T | N/A | GCGCTAATACGACTCACTATAGGG **TAGCTT** **ATCAGACTtATGTTGA** | IVT-IDT Probes |
| T9G | N/A | GCGCTAATACGACTCACTATAGGG **TAGCTT** **ATCAGACcGATGTTGA** | IVT-IDT Probes |
| C10A | N/A | GCGCTAATACGACTCACTATAGGG **TAGCTT** **ATCAGAaTGATGTTGA** | IVT-IDT Probes |
| A11C | N/A | GCGCTAATACGACTCACTATAGGG **TAGCTT** **ATCAGgCTGATGTTGA** | IVT-IDT Probes |
| G12T | N/A | GCGCTAATACGACTCACTATAGGG **TAGCTT** **ATCAtACTGATGTTGA** | IVT-IDT Probes |
| A12C | N/A | GCGCTAATACGACTCACTATAGGG **TAGCTT** **ATCgGACTGATGTTGA** | IVT-IDT Probes |
| C14A | N/A | GCGCTAATACGACTCACTATAGGG **TAGCTT** **ATaAGACTGATGTTGA** | IVT-IDT Probes |
| T15G | N/A | GCGCTAATACGACTCACTATAGGG **TAGCTT** **AcCAGACTGATGTTGA** | IVT-IDT Probes |
| A16C | N/A | GCGCTAATACGACTCACTATAGGG **TAGCTT** **gTCAGACTGATGTTGA** | IVT-IDT Probes |
| Duplex DNA | Sense | GAAGTTCAT GTTTCTGAA TCAACATCAGTCTGAT AAGCTA TTCAAG | IDT duplexing |
| | Anti-Sense | TTCAGAAACATGAACTTC | |
| AsCas12a gRNA | T7-DNA Template | GCGCTAATACGACTCACTATAGGG TAATTTCTACTCTTGTAGAT TGAA TCAACATCAGTCTGAT | PCR_IVT |
| | FW_Primer | GCGCTAATACGACTCAC | |
| | Rev_Primer | ATCAGACTGATGTTGATTCA | |

*Sequence in blue is miRNA-21 when it is transcribed
**Underlined sequence is the domain that bind to gRNA
***Letters in lowercase denote point mutations
****IVT means in vitro transcription

**Table S16:** Nucleic acid sequences used for RAPID-LAMP assay for SARS-CoV-2 Detection.

| Name | Sub-Template | Sequences (5′-3′) | SEQ ID NO: |
|---|---|---|---|
| SARS_CoV-2_N_Ref2B | RNA Target | CCAAAAGAUCACAUUGGCACCCGCAAUCCUGCUAACAAUGC UGCAAUCGUGCUACAACUUCCUCAAGGAACAACAUUGCCAA AAGGCUUCUACGCAGAAGGGAGCAGAGGCGGCAGUCAAGCC UCUUCUCGUUCCUCAUCACGUAGUCGCAACAGUUCAAGAAA UUCAACUCCAGGCAGCAGUAGGGGAACUUCUCCUGCUAGAA UGGCUGGCAAUGGCGGU | IVT RNA |
| LAMP Primers | Ref2B_N_F3 | AGATCACATTGGCACCCG | Standard probes from IDT |
| | Ref2B_N_B3 | CCATTGCCAGCCATTCTAGC | |
| | Ref2B_N_FIP | TGCTCCCTTCTGCGTAGAAGCCAATGCTGCAATCGTGCTAC | |
| | Ref2B_N_BIP | GGCGGCAGTCAAGCCTCTTCCCTACTGCTGCCTGGAGTT | |
| | Ref2B_N_LF | GCAATGTTGTTCCTTGAGGAAGTT | |
| | Ref2B_N_LB | GTTCCTCATCACGTAGTCGCAACA | |

| | | | |
|---|---|---|---|
| LAMP Amplicon | N/A | GGCGGCAGTCAAGCCTCTTCCCTACTGCTGCCTGGAGTTGAAT TTCTTGAACTGTTGCGACTACGTGATGAGGAACGAGAAGAGG CTTGACTGCCGCCTCTGCTCCCTTCTGCGTAGAAGCCTTTTGGC AATGTTGTTCCTTGAGGAAGTTGTAGCACGATTGCAGCATT G GCTTCTACGCAGAAGGGAGCA | From LAMP reaction |
| SARS_CoV-2 Duplex Probe_Loop 1 | Sense | GAAGTTCAT GTTTCTGAA CCTCATCACGTAGTCGCAACAGTTCAAG | IDT duplexing |
| | Anti Sense | TTCAGAAACATGAACTTC | |
| SARS_CoV-2 Duplex Probe_Loop 2 | Sense | GAAGTTCAT GTTTCTGAA TTTGGCAATGTTGTTC CTTGAGGAAGTT | IDT duplexing |
| | Anti Sense | TTCAGAAACATGAACTTC | |
| LbCas12a gRNA_Loop 1 | T7_Template | GCGCTAATACGACTCACTATAGGG TAATTTCTACTAAGTGTAGAT TGAA CCTCATCACGTAGTCG | PCR_IVT |
| | FW_Primer | GCGCTAATACGACTCAC | |
| | Rev_Primer | CGACTACGTGATGAGGT | |
| LbCas12a gRNA_Loop 2 | T7_Template | GCGCTAATACGACTCACTATAGGG TAATTTCTACTAAGTGTAGAT TGAA TTTGGCAATGTTGTTC | PCR_IDT |
| | FW_Primer | GCGCTAATACGACTCAC | |
| | Rev_Primer | GAACAACATTGCCAAATTCA | |

*PCR_IVT means that PCR was performed to amplify the ssDNA substrate before performing in vitro reaction
**IDT_Duplexing means that the duplexing was done by IDT and shipped dry to us

**Table S17**: Synthetic RNA controls for molecular reactions. Target synthetic RNA controls were obtained from Twist Bioscience.

| Twist RNAs | Catalog Number |
|---|---|
| Wuhan virus | 102024 |
| Alpha | 103907 |
| Beta | 104043 |
| Gamma | 104044 |
| Delta | 104533 |
| Omicron | 105204 |

**Table S18**: RT-qPCR oligos used in this study for SARS-CoV-2.

| Name | Sequence (5’-3’)* |
|---|---|
| 2019-nCoV_N1 Forward Primer | GACCCCAAAATCAGCGAAAT |
| 2019-nCoV_N1 Reverse Primer | TCTGGTTACTGCCAGTTGAATCTG |
| 2019-nCoV_N1 Probe | FAM-ACCCCGCATTACGTTTGGTGGACC-BHQ1 |
| RNase P Forward Primer | AGATTTGGACCTGCGAGCG |
| RNase P Reverse Primer | GAGCGGCTGTCTCCACAAGT |
| RNase P Forward Probe | FAM-TTCTGACCTGAAGGCTCTGCGCG-BHQ1 |

*U.S. CDC recommended protocol for testing patient samples.

**Table S19**: Comparison of SARS-Cov2 patient sample data with RAPID and RT-qPCR.

| ID | Ct (N1-SARS-CoV-2) | RNA copy number | Ct (RNaseP) | Final disposition in parallel with the RT-qPCR |
|---|---|---|---|---|
| 1 | ND | 0 | 29.7 | True negative |
| 2 | ND | 0 | 30.2 | True negative |
| 3 | ND | 0 | 32.8 | True negative |
| 4 | ND | 0 | 35.5 | True negative |
| 5 | ND | 0 | 28.6 | True negative |
| 6 | 25.5 | 45449.2 | 31.4 | True positive |
| 7 | 24 | 123358.8 | 34.3 | True positive |
| 8 | 22.2 | 398830.4 | 32.8 | True positive |
| 9 | 24.2 | 109784.4 | 32.4 | True positive |
| 10 | 26.1 | 31025.9 | 33.6 | True positive |
| 11 | ND | 0 | 32.6 | True negative |
| 12 | ND | 0 | 32 | True negative |
| 13 | ND | 0 | 31.9 | True negative |
| 14 | ND | 0 | 30 | True negative |
| 15 | ND | 0 | 30.4 | True negative |
| 16 | 32.4 | 488.3 | 32.4 | True positive |
| 17 | 27.1 | 15697.1 | 33 | True positive |
| 18 | 23.6 | 161040.6 | 31.4 | True positive |
| 19 | 26.9 | 18313.4 | 31.7 | True positive |
| 20 | 28.3 | 7006.1 | 33.2 | True positive |
| 21 | 34.5 | 14.0 | 33.7 | False negative |
| 22 | 34.4 | 15.0 | 37.2 | False negative |
| 23 | 33.3 | 34.2 | 30.6 | False negative |
| 24 | 30.3 | 285.6 | 35.0 | True positive |
| 25 | 33.6 | 27.1 | 28.8 | False negative |

**Table S20**: Patient samples (Ct value <40).

| Statistic | Value (95% CI) |
|---|---|
| **Clinical sensitivity** | 73.33% (44.90% to 92.21%) |
| **Clinical specificity** | 100.00% (69.15% to 100.00%) |

| | |
|---|---|
| **Disease prevalence** | 60.00% (38.67% to 78.87%) |
| **Accuracy** | 84.00% (63.92% to 95.46%) |

* To check the calculations, visit: https://www.medcalc.org/calc/diagnostic_test.php

**Table S21**: Table of statistical analyses at a 95% confidence interval for clinical sensitivity, specificity, and disease prevalence, highlighting that RAPID achieved 100% accuracy in samples with a cycle threshold (Ct) value ≤33.

| **Statistic** | **Value (95% CI)** |
|---|---|
| **Clinical sensitivity** | 100.00% (71.51% to 100.00%) |
| **Clinical specificity** | 100.00% (69.15% to 100.00%) |
| **Disease prevalence** | 52.38% (29.78% to 74.29%) |
| **Accuracy** | 100.00% (83.89% to 100.00%) |
